# Distinctive cross-ancestry genetic architecture for age-related macular degeneration

**DOI:** 10.1101/2022.08.16.22278855

**Authors:** Bryan R. Gorman, Georgios Voloudakis, Robert P. Igo, Tyler Kinzy, Christopher W. Halladay, Tim B. Bigdeli, Biao Zeng, Sanan Venkatesh, Jessica N. Cooke Bailey, Dana C. Crawford, Kyriacos Markianos, Frederick Dong, Patrick Schreiner, Wen Zhang, VA Million Veteran Program, International AMD Genomics Consortium (IAMDGC), Tamer Hadi, Matthew D. Anger, Amy D. Stockwell, Ronald B. Melles, Jie Yin, Hélène Choquet, Rebecca Kaye, Karina Patasova, Praveen J. Patel, Brian L. Yaspan, Eric Jorgenson, Pirro G. Hysi, Andrew J. Lotery, J. Michael Gaziano, Philip S. Tsao, Steven J. Fliesler, Jack M. Sullivan, Paul B. Greenberg, Wen-Chih Wu, Themistocles L. Assimes, Saiju Pyarajan, Panos Roussos, Neal S. Peachey, Sudha K. Iyengar

## Abstract

To effectively reduce vision loss due to age-related macular generation (AMD) on a global scale, knowledge of its genetic architecture in diverse populations is necessary. A critical element, AMD risk profiles in African and Hispanic/Latino ancestries, remains largely unknown due to lower lifetime prevalence. We combined genetic and clinical data in the Million Veteran Program with five other cohorts to conduct the first multi-ancestry genome-wide association study of AMD and discovered 63 loci (30 novel). We observe marked cross-ancestry heterogeneity at major risk loci, especially in African-ancestry populations which demonstrate a primary signal in a Major Histocompatibility Complex Class II haplotype and reduced risk at the established *CFH* and *ARMS2/HTRA1* loci. Broadening efforts to include ancestrally-distinct populations helped uncover genes and pathways which boost risk in an ancestry-dependent manner, and are potential targets for corrective therapies.

**One Sentence Summary:** Probing electronic health record data with genomics unearths novel paths to age-related macular degeneration.

## Introduction

Blindness is one of the most feared health problems, reflecting the critical role that vision plays in daily living. Age-related macular degeneration (AMD) is a common disorder that impacts the central retina region or macula which is specialized for high acuity vision. Because AMD spares the peripheral retina, patients retain useful side vision, but the loss of central vision interferes with important daily activities such as reading, close work, driving, and recognizing faces.

AMD is a progressive disease that alters various histological layers of the retina, although progression rates vary by age, sex, and ancestry (*1*). Lipoproteinaceous deposits called drusen formed between the Bruchs’ membrane and the retinal pigment epithelium (RPE) are the earliest indications. The clinical presentation of dry AMD typically involves drusen, pigmentary abnormalities and geographic atrophy occupying the central fovea. Decreased choroidal perfusion and retinal tissue oxygenation may stimulate choroidal neovascularization, resulting in wet AMD.

AMD shows differential risk across diverse populations (*2, 3*) with Black subjects being 74% less likely to receive an AMD diagnosis compared to non-Hispanic white subjects after adjustment for age and sex (*3*). In the same study, Hispanic/Latino subjects showed a 44% reduction, while Asian subjects showed a 19% reduction in AMD diagnosis. The cause of the reduced risks in these populations is not well understood, although genetic or environmental exposure differences, or disparities in life expectancy are likely factors (*4*). While genome-wide association studies (GWAS) for AMD have been published for European ancestry (EA) and Asian ancestry (AS) populations, comparable analyses have not been conducted for African ancestry (AA) or Hispanic/Latino-ancestry (HA) populations in part because of lower AMD prevalence.

GWAS have been highly successful in exploring the etiology of AMD; *CFH* is widely regarded as the first disease susceptibility locus discovered through a GWAS (*5–7*). The largest genetic analysis for AMD focused primarily on EA populations (*8*), leaving the question of risk in other ancestral groups unanswered. A central feature of the current investigation is the multi-ancestry nature of the biobank-scale sample available from the Department of Veterans Affairs (VA) Million Veteran Program (MVP) (*9*). Examining 61,248 cases and 364,472 controls from multiple cohorts, we discovered 30 new genetic loci, and validated 33 known loci. In this study, we synthesize several lines of evidence to show that decreased AMD prevalence in AA is at least partly mediated by reduced risk at *CFH*, and other alternative complement pathway genes, allowing effects of other loci to be isolated. This work underscores the value of inclusive population studies to advance our mechanistic understanding of AMD and likely other diseases as well.

## Results

### Epidemiology of AMD in MVP

An overview of the present study is depicted in **Fig. 1**, with additional details in **Fig. S1. Table S1** summarizes the AMD cohorts used for our analyses. Among demographic factors **(Tables S2a, S2b)**, not surprisingly, increased age was significantly associated with AMD status. Female sex was also significantly associated with AMD status, even in MVP, where 91% of participants are male. In a multiple regression model, smoking and history of heavy drinking (score of 8 or higher on at least one AUDIT-C questionnaire) were significant risk factors in the MVP EA cohort, and were positively associated but below significance in AA or HA. Elevated HDL cholesterol was significantly associated in the EA and AA cohorts, while BMI showed a modest negative association with AMD in the EA cohort; a standard deviation increase was associated with an odds ratio (OR) of 0.96 [0.95-0.98]; p=2×10^-6^), even in the fully adjusted model, which was replicated with similar effect size (though not with statistical significance) in the AA and HA cohorts. Reduced serum albumin was significantly associated with AMD in all three cohorts, with similar effect sizes.

**Fig. 1.**
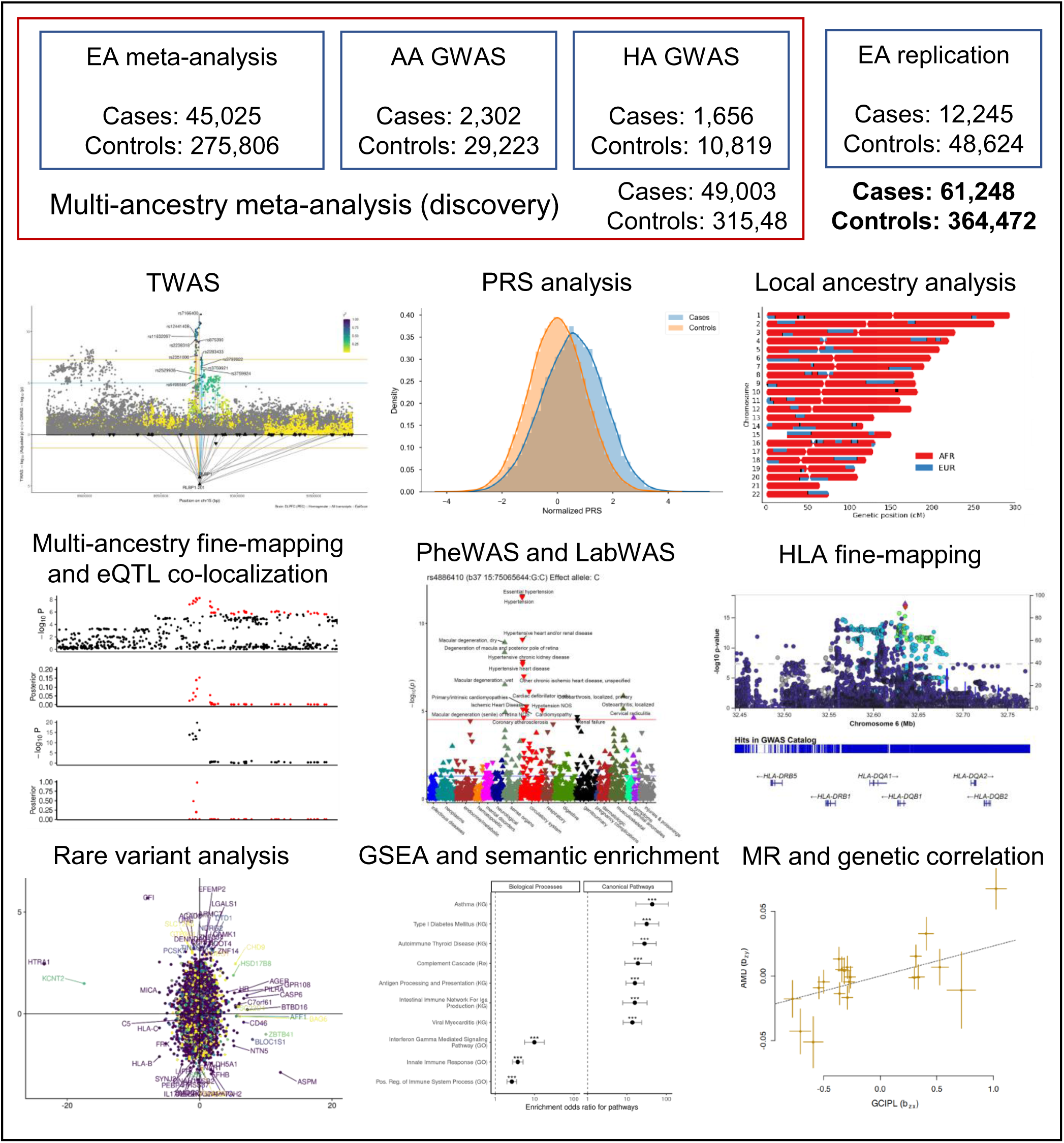
Overview of AMD GWAS meta-analysis primary and secondary analyses. Abbreviations: EA, European ancestry; AA, African ancestry; HA, Hispanic/Latino ancestry; TWAS, transcriptome-wide association study; PRS, polygenic risk score; eQTL, expression quantitative trait loci; PheWAS, phenome-wide association study; LabWAS, lab measurement-wide association study; HLA, human leukocyte antigen; GSEA, gene set enrichment analysis; MR, Mendelian randomization.

Using summary statistics from a prior GWAS conducted by the International AMD Genomics Consortium (IAMDGC) (*8*), we constructed polygenic risk scores (PRS) in MVP using PRS-CS software (*10*). We observed strong evidence of increasing penetrance of AMD with genetic risk and age in MVP males (**Fig. 2a**), ranging from a prevalence of approximately 35% in the top PRS decile within the 75-85 age range, compared to less than 10% in the bottom decile. History of heavy alcohol use and smoking are significant lifestyle risk factors **(Table S2b)**, yet they demonstrate smaller changes in penetrance with age compared to the PRS.

**Fig. 2.**
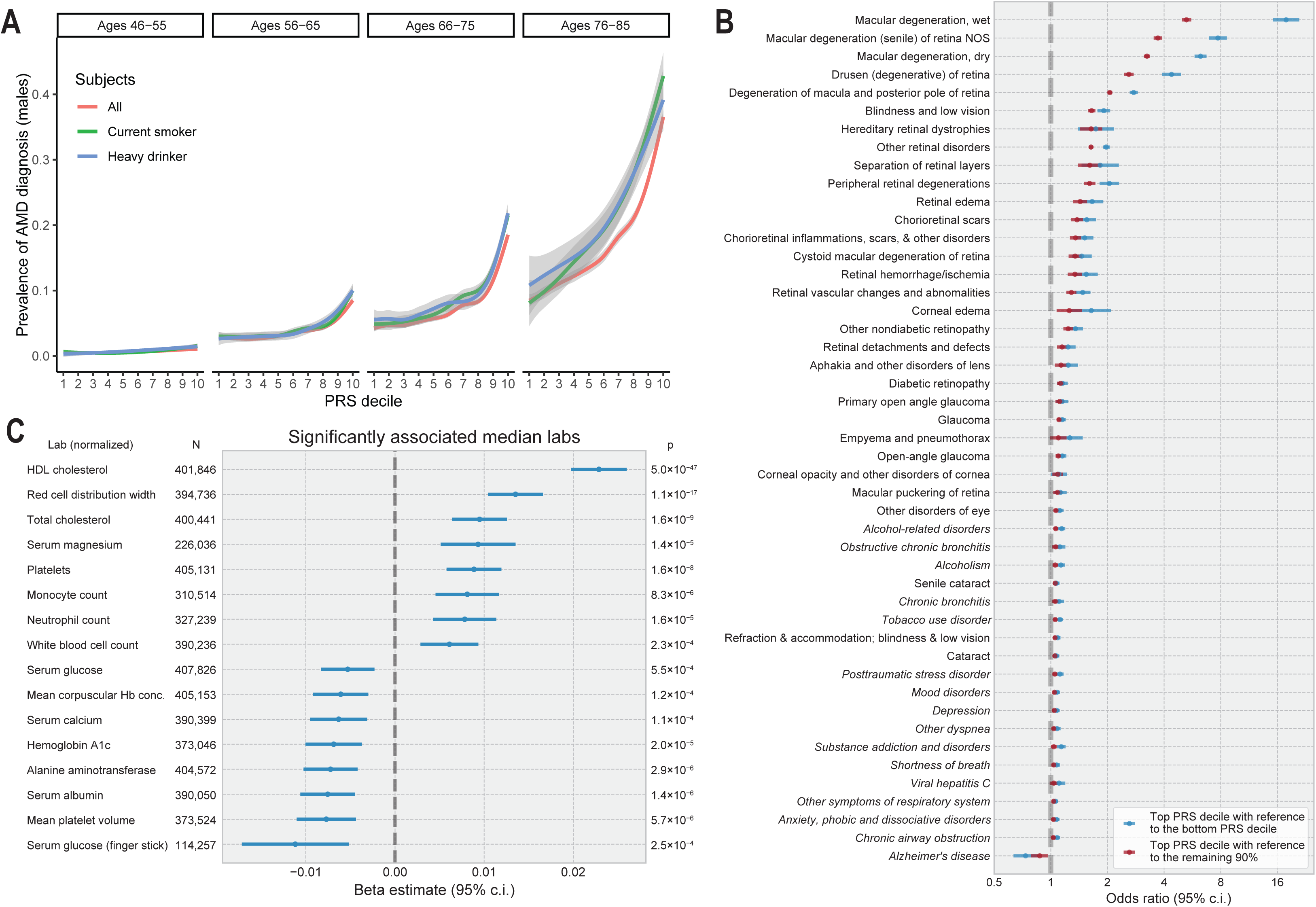
Penetrance and pleiotropy of the AMD PRS. (**A**) Prevalence of AMD in EA males in MVP as a function of PRS decile, age range, and lifestyle risk factors. (**B**) PheWAS of the AMD PRS with 1,665 ICD code-based binary phenotypes in MVP EA. Phenotypes with association p-values less than 1×10^−5^ are shown. Odds ratios contrasting the top decile of PRS vs. the bottom decile (blue) and the top decile vs. the remaining 90% (red) are presented, with the x-axis in log scale. Phenotypes not directly related to vision are italicized. (**C**) LabWAS of the AMD PRS with 69 quantitative clinical lab measurements in MVP EA. Labs significant after Bonferroni adjustment for the number of labs are shown.

To quantify if there were an overall reduction in risk for AMD in other ancestry groups across the entire MVP cohort, we performed a Cox proportional-hazards model with time to AMD diagnosis as the outcome and found that AA veterans had a 57% reduction in risk and HA veterans had a 26% reduction in risk, relative to EA veterans **(Table S2c)**. Supporting this overall result, the PRS demonstrated weak transferability to non-European ancestries in our selected case-control sample **(Fig. S2)**: in our EA sample, a standard deviation increase in the PRS was associated with an AMD OR=1.76 [95% confidence interval (CI) 1.73-1.78] in a fully adjusted model **(Table S2b)**, but only OR=1.13 [1.08-1.18; p=1.2×10^-7^] in our AA sample and OR=1.47 [1.39-1.56; p=1.6×10^-41^] in our HA sample.

We next interrogated pleiotropy of the AMD PRS using diagnostic codes (i.e., phenome-wide association study, or PheWAS) and laboratory measures (i.e., laboratory-wide association study, or LabWAS) recorded in the EHR. We evaluated PheWAS associations of 1,665 ICD code-based phenotypes with the normalized PRS and contrasted the top vs. bottom deciles and top decile vs. remaining 90% to evaluate risk stratification. Fifty-four phenotypes were significantly associated **(Fig. 2b; Table S3)**. As expected, retinal codes were highly significant -- compared to the bottom PRS decile, the top decile had OR=17.8 [15.1-20.9] for a diagnosis of wet AMD. Unsurprisingly, alcohol and tobacco exposure also showed strong associations. Other notable positive associations included cataracts, refractive error, and mood disorders. Interestingly, the AMD PRS was also associated with lowered risks of Alzheimer’s disease and other dementias, which may reflect opposing roles of HDL cholesterol. Of the 69 lab measurements, 16 were significantly associated, including positive associations with HDL cholesterol, red cell distribution width, platelets, serum magnesium, monocytes, and neutrophils, compared with negative associations with triglycerides, serum albumin and serum glucose **(Fig 2c; Table S4)**.

### European meta-analysis for AMD identifies 27 novel loci

GWAS of AMD in two MVP tranches and meta-analysis with the summary statistics from five other cohorts (IAMDGC, GERA, UK Biobank, Genentech geographic atrophy, and Genentech choroidal neovascularization) was followed by replication, i.e., meta-analysis with a third MVP tranche that later became available to us, for a total EA analysis comprising 57,290 AMD cases and 324,430 controls **(Table S1)**. This analysis (**Fig. 3a**) revealed 60 index variants: 33 previously reported and 27 novel significant loci **(Table S5; Figs. S3-S4)**. Due to oversampling for severity in the IAMDGC, effect sizes of the index variants at major association peaks are lower overall in the MVP EA cohorts, similar to other EHR-based cohorts **(Table S6)**. However, genetic correlation (*11*) between the MVP and IAMDGC GWAS was very high (r_G_=0.95; se=0.04). Of the 34 loci reported by the IAMDGC (*8*), 31 are genome-wide significant in the EA meta-analysis; the exceptions are *PRLS-SPEF2*, *TRPM3*, and *VTN*.

**Fig. 3.**
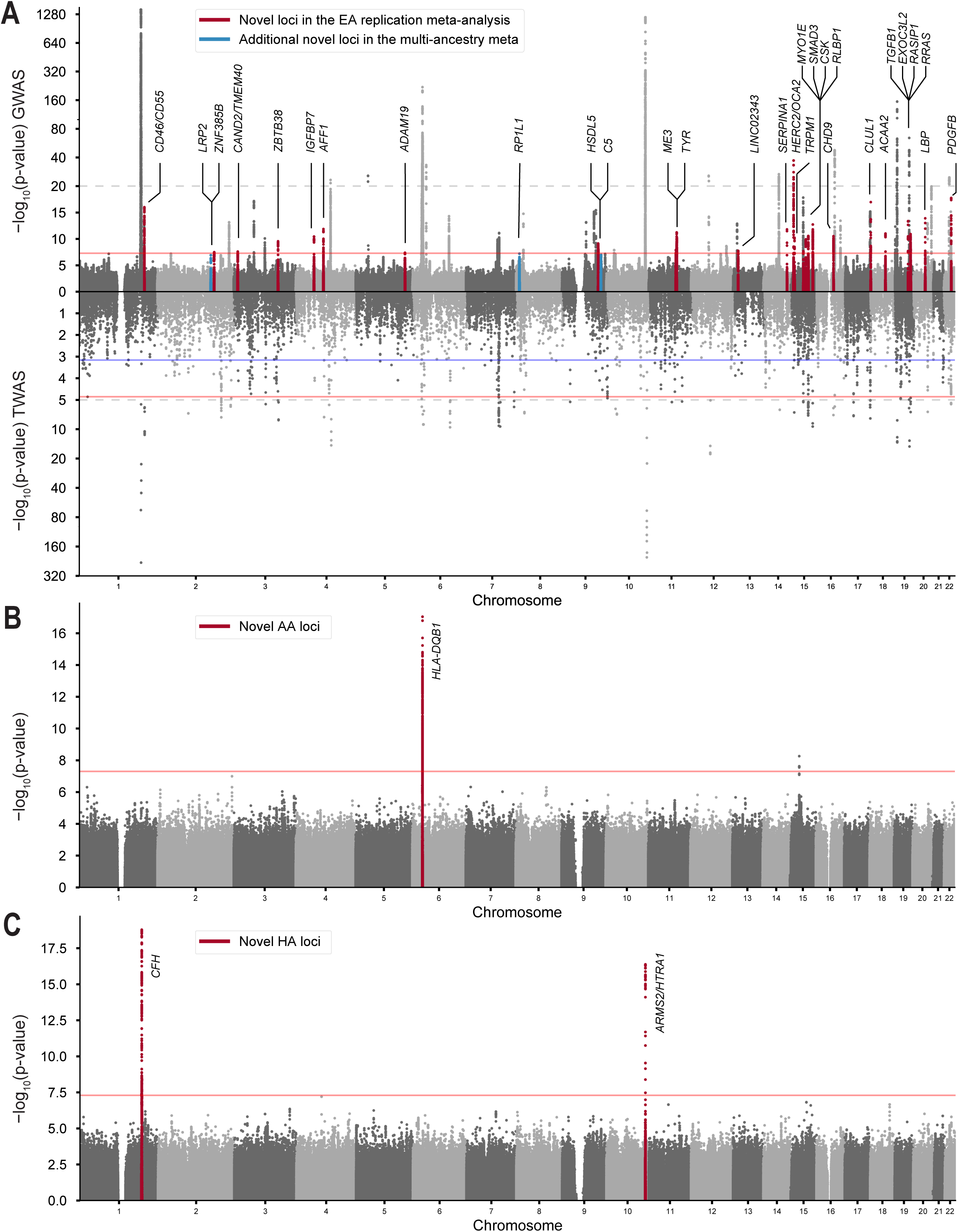
GWAS analyses identify 27 novel loci in EA and the first loci in AA and HA. (**A**) Miami plot of the EA GWAS replication meta-analysis (57,290 cases, 324,430 controls) and DLPFC TWAS (45,045 cases, 275,806 controls). Red lines: genome-wide significance in the GWAS and TWAS analyses; blue line: FDR-corrected association p-value < 0.05 in the DLPFC TWAS; dashed gray line: transition from linear to log-scale on the y-axis. (**B**) Manhattan plot of the AA GWAS (2,302 cases, 29,223 controls). (A second potential locus near *COPS2* could not be confirmed in other ancestries.) (**C**) Manhattan plot of the HA GWAS (1,656 cases, 10,819 controls).

We observed novel loci in or near genes related to complement cascade regulation (*CD46/CD55*), TGF-β signaling (*TGFB1*, *SMAD3*, *ADAM19*), pigmentation (*TYR, HERC2/OCA2, TRPM1*), angiogenesis and vascular homeostasis (*RRAS*, *RASIP1, IGFBP7*, *PDGFB*, *MYO1E*, *EXOC3L2*), inflammation (*CSK/ULK3*), proteolysis (*SERPINA1*), cell proliferation and apoptosis (*ZBTB38*, *ZNF385B*), ubiquitin-proteasome system (*CAND2/TEMEM40*), lipid metabolism and biogenesis (*CHD9*, *LBP*, *HSDL2*, *AFF1*, *ACAA2/LIPG*, *ME3*), and photoreceptor function (*RLBP1, CLUL1*). Finally, we observed a novel locus in an intergenic region near *LINC02343*, a non-coding RNA of uncharacterized function. A conditional analysis revealed 176 mutually significant variants in or near the 60 loci **(Table S7)**.

At the 20q13 locus, the previously reported *MMP9* index SNP was not replicated in our meta-analysis, and a new signal at nearby *PLTP* (phospholipid transfer protein), a *CETP* and *LBP* paralog, emerged. Although partially in LD (r^2^ = 0.2), conditioning on the *MMP9* SNP (rs1888235) in our meta-analysis, and on the *PLTP* SNP (rs17447545) in IAMDGC, weakened but did not eliminate either signal, demonstrating independence **(Fig. S5)**. This result is consistent with the reported specificity of the *MMP9* signal for wet AMD, which is enriched in IAMDGC (*8*).

Rare protective missense mutations in novel genes *CFD* and *RRAS* emerged, independent of any common variation. The *CFD* missense mutation (p.Glu69Lys, rs35186399), which was not genotyped in the IAMDGC study, has OR=0.73 [0.66-0.81; p=8.0×10^-10^], with a frequency just under 1%, and is a pQTL for decreased complement C8 levels (*12*). Although rs35186399 is not predicted by most variant effect classifiers to have a significant impact on protein function (*13, 14*), it is within a CTCF binding site, which could potentially affect the insulation of topologically associated domains and enhancer-promoter regulatory interactions. We also identified a novel protective mutation in *RRAS*, a Ras-family GTPase with reported regulatory functions in angiogenesis and vascular homeostasis (*15–17*). The putatively protein-altering mutation, rs61760904 (p.Asp133Asn), has a frequency of 0.6% in MVP EA, with OR=0.75 [0.68- 0.83; p=9.5×10^-9^], and also associates with increased blood pressure (*18*).

At *SERPINA1*, which encodes α-1 antitrypsin (A1AT), we observed associations with the pathogenic PiZ (p.Glu366Lys; rs28929474) and PiS (p.Glu288Val; rs17580) alleles. Intriguingly, PiZ is protective for AMD (OR=0.81 [0.77-0.86; p=3.2×10^-12^]), while PiS increases risk (OR=1.10 [1.06-1.14; p=1.2×10^-7^]). A1AT is a serine protease inhibitor primarily expressed in the liver which regulates proinflammatory neutrophil elastase. Notably, A1AT is cleaved by HTRA1 to form NET (neutrophil extracellular trap) inhibitory peptides (NIPs) (*19*). Z and S allele heterozygosity are associated with numerous inflammatory biomarkers, including serum albumin (*20*), C reactive protein, and glycoprotein acetyls (*21*).

Genetic correlations with a number of eye traits and related phenotypes revealed some of the strongest correlations with pigment dispersion syndrome and glaucoma (*22*), as well as eye color (*23*) and hair color (*24*) **(Table S8)**. Similarly, genetic risk to intraocular pressure was inversely correlated with that of AMD. We further scanned genetic correlations with blood-based biomarkers (Pan-UKB team, 2020) and found the strongest correlations with serum albumin (r_G_=- 0.13; p=4.2×10^-8^), albumin/globulin ratio, and kidney biomarkers phosphate, calcium, and cystatin C **(Table S9)**.

We further explored the causality of pigmentation associations using Mendelian randomization (MR) **(Fig. 4; Table S10)**. Drawing instrumental variables from some of the largest studies of pigmentation, we report for the first time a causal effect of pigmentation on AMD in EA populations. Comparing AMD data with those from a study (*23*) in which eye color was measured on a quantitative scale (range 0-6) from blue to dark brown, we find that each step towards darker eye color decreases the risk of AMD by about 5% (OR=0.95 [0.94-0.97]). Selecting only the instruments with known associations in other pigmentary traits, which likely represent the best proxies for melanin metabolism in the retina (*23*), we obtain a similar effect estimate (OR=0.95 [0.93-0.98]). To further confirm the involvement of melanin metabolic pathways in AMD, we examined other pigmentary traits. We found that each step up from blond to black hair color (range 0-4) (*24*) decreases the risk of AMD by 12% (OR=0.88 [0.85-0.92]). The more modest protective effect of eye color relative to hair color may be due to the higher phenotypic contribution of light diffraction genes (*23*). Finally, we find also that proxies of skin color, such as non-melanoma skin cancers (*25*) (OR=1.07 [1.02-1.12]) and low tanning response (*26*) (OR=1.07 [1.02-1.13]) are significantly associated in MR models with higher AMD risk. We strongly believe that these examples are simply proxies for fundamental pigmentation processes and approximate the presence of melanin in the RPE and choroid.

**Fig. 4.**
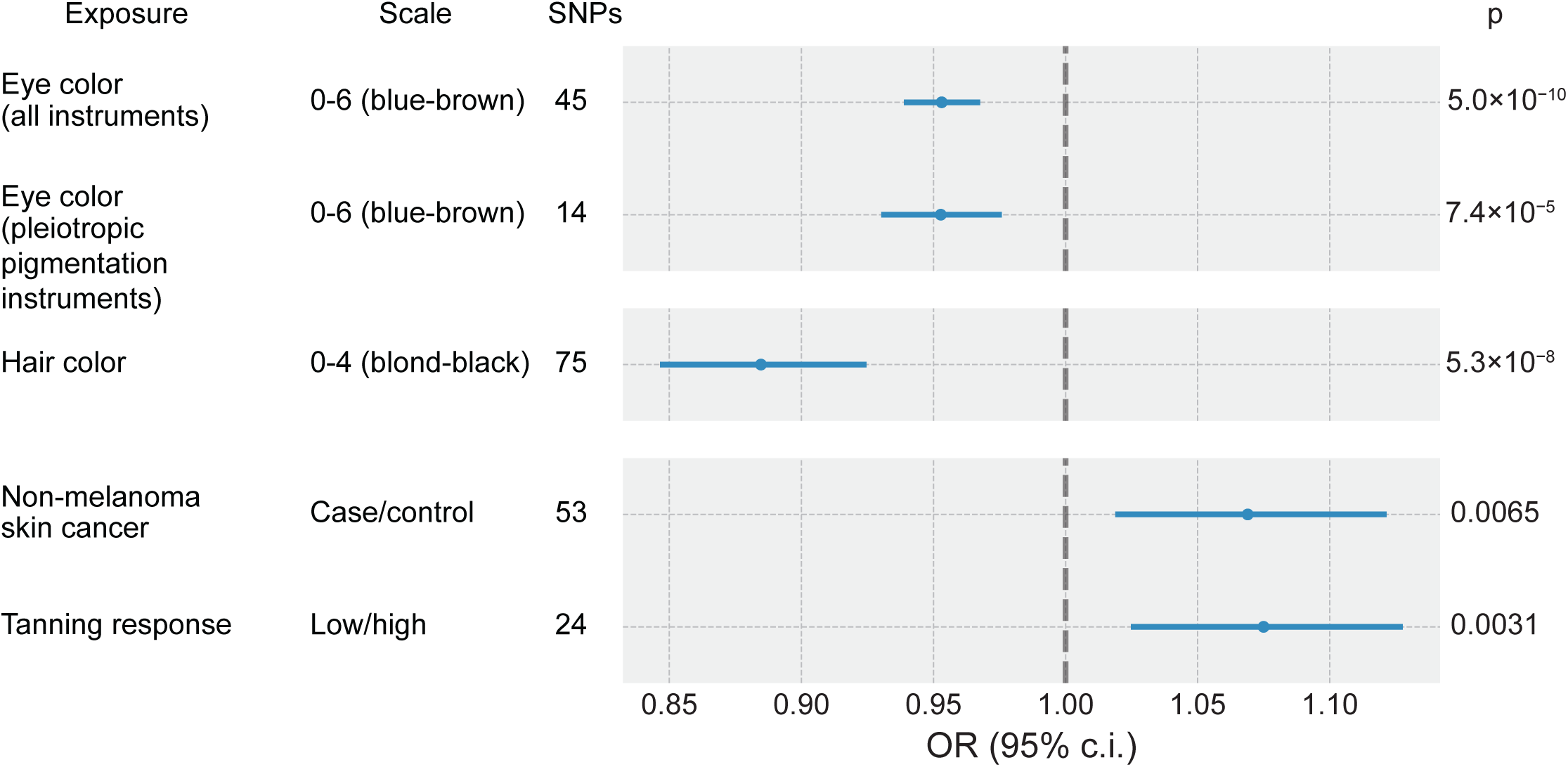
MR of pigmentation traits on AMD risk in European ancestries. Forest plot of OR and 95% confidence intervals from the inverse variance-weighted MR analyses are shown. Eye color (*23*) was analyzed using all genome-wide significant instruments and using a selection of 14 instruments with known pleiotropic associations with other pigmentation traits. Other traits representing proxies for pigmentation, such as hair color (*24*), non-melanoma skin cancer (*25*), and tanning response (*26*) were also analyzed.

### Multi-ancestry GWAS for AMD identifies the first loci in admixed Black and Hispanic-Latino populations

We conducted the first GWAS on AMD in individuals of African and Hispanic/Latino ancestries. In the AA GWAS (2,302 cases and 29,223 controls), a primary locus centered at *HLA-DQB1* (index SNP rs3844313; OR=1.41 [1.30-1.52]; p=9.1×10^-18^) reached genome-wide significance (**Figs. 3b** **and S6)**. Supporting the AA result, the *HLA-DQB1* locus was significant in the EA GWAS at a direction-consistent but lower OR **(Fig. 5a; Table S11)**. In contrast to the EA population, much smaller effect sizes were observed in the AA GWAS at major AMD risk loci *CFH*, *ARMS2*, and *CFB/C2* (**Fig. 5a**). However, EA index markers have similar allele frequencies in the MVP AA sample.

**Fig. 5.**
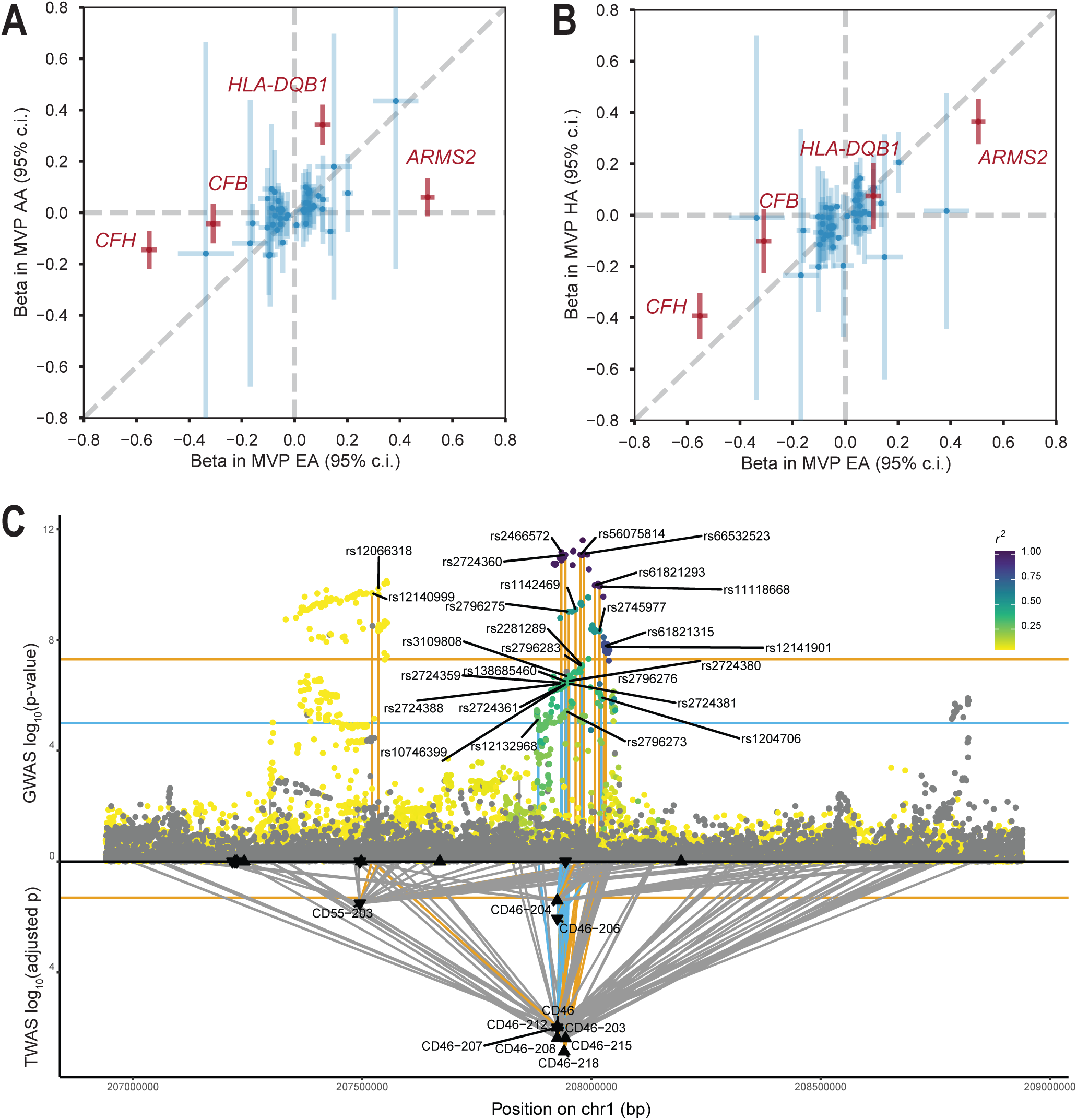
Highlighted results from the multi-ancestry GWAS and TWAS. (**A**) Comparison of effect sizes (log-OR) of EA and AA index SNPs in the MVP EA GWAS (x-axis) and the MVP AA GWAS (y-axis). (**B**) Comparison of effect sizes in the MVP EA GWAS (x-axis) and the MVP HA GWAS (y-axis). Highlighted highly heterogeneous loci are shown in red. Error bars correspond to 95% confidence intervals. (**C**) Regional Miami plot of genetically regulated gene expression at the 1q32 (*CD46/CD55*) locus. GWAS results are above the axis and TWAS results from the DFPLC brain model below the axis. GWAS points are colored according to linkage disequilibrium *r^2^* with the top SNP.

We constructed a new AA-specific PRS based on the AA GWAS and used it to score unrelated AA subjects in MVP that were not included in our GWAS cohort. We then performed PheWAS and LabWAS on the AA PRS (N up to 80,345 depending on the phenotype) to better understand the pleiotropy of genetic risk in AA populations. We observed significant increases in median neutrophil count, white blood cell count, neutrophil fraction, and HDL cholesterol, and a decrease in lymphocyte fraction **(Table S12)**. This result demonstrates that lipid metabolism and immune pathways continue to play a role in AMD pathogenesis in AA despite divergent genetic architecture.

In the HA GWAS (1,545 cases and 10,930 controls), *CFH* and *ARMS2* reached genome-wide significance (**Fig. 3c** **and S6; Table S13)**, despite smaller sample size than AA. The HA GWAS showed effect sizes more comparable to the EA estimates, though still generally smaller at the major EA loci (**Fig. 5b**). In summary, AMD risk is attenuated in AA and HA which we ascribe at least partly to genetic precursors, with AA showing the most dissimilar profiles to EA.

### Multi-ancestry meta-analysis and fine-mapping identifies three additional novel loci and candidate causal variants

To capture cross-ancestry effects and to increase discovery power, we conducted a multi-ancestry meta-analysis, combining our EA meta-analysis, EA replication GWAS, and AA and HA GWAS reaching a total of 61,248 cases and 364,472 controls. A total of 62 loci were genome-wide significant, including three novel loci beyond that reported above for the EA meta-analysis: *LRP2*, *RP1L1*, and complement *C5* **(Table S14; Fig. S7)**, and all significant EA loci except for *ADAM19*. *LRP2* encodes the transmembrane low-density lipoprotein receptor megalin, which is expressed in the RPE (*27, 28*), and *RP1L1* encodes a retina-specific protein involved in photoreceptor differentiation and function (*29*).

We leveraged differential LD structure across ancestries, along with multi-model expression data, to identify candidate causal variants. We fine-mapped genome-wide significant loci in the multi-ancestry and EA discovery meta-analyses using the program FINEMAP (*30*) **(Tables S15-S16)**. To co-localize eQTLs, multivariate multiple QTL (mmQTL) (*31*) was first applied to detect eQTL, and eCAVIAR (*32*) was then used to obtain a co-localization posterior probability (CLPP) for each GWAS credible set variant. Based on co-localization with the multi-ancestry meta-analysis, we identified seven candidate causal variants modifying the gene expression of *CD46, RDH5*, *BLOC1S1, FAM227A*, *CFI*, *TNFRSF10A*, and *ABCA1* **(Fig. S8; Table S17)**. Expression co-localization of the variant with the highest CLPP, *TNFRSF10A*, is shown in **Fig. S8a**. *TNFRSF10A* encodes for a receptor, which is activated by tumor necrosis factor and induces cell apoptosis.

### Genetic risk at *CFH* in admixed Black and Hispanic/Latino populations comes from European haplotypes

We conducted local ancestry analyses to investigate the contribution of genetic risk at *CFH* in admixed individuals. We inferred local ancestry (*33*), and extracted ancestry-specific imputed dosages and haplotype counts. On average, the MVP AA cohort had 81% African (AFR) and 19% European (EUR) admixture, and the MVP HA cohort had 31% Native American (NAT), 61% EUR, and 8% AFR admixture. We used Fisher’s Exact Test to test for significant differences in the proportion of cases and controls with EUR haplotypes at *CFH* **(Table S18)**. We conducted a 2×2 test on haplotypes in AA subjects, which was statistically significant (OR=1.12 [1.04-1.20]; p=0.0033) and a 3×2 test on haplotypes in admixed HA subjects (AFR/EUR/NAT x case/control) which was also statistically significant (p=1.7×10^-5^).

The *CFH* locus was tested using the Tractor method (*34*), a local ancestry-aware GWAS model which partitions the contributions to genetic risk according to the ancestry of origin of each haplotype to obtain ancestry-specific marginal effect size estimates (**Fig. 6**). In AA subjects, the conventional GWAS found a modest effect at the established risk polymorphism *CFH* Y402H (rs1061170) with OR=1.13 [1.06-1.20, p=1.8×10^-4^]. However, the Tractor GWAS found differential effect across haplotype ancestry, with AFR haplotypes (“AFR tract”) having OR=1.07 [1.00-1.15; p=0.045] and EUR haplotypes (“EUR tract”) having OR=1.40 [1.23-1.60; p=7.1×10^-7^] (**Fig. 6c**). Moreover, the genetic architecture of the EUR tract was similar to the analysis conducted within EA individuals (**Fig. 6a**). Within HA subjects (**Fig. 6b-c**), however, the conventional GWAS demonstrated a large effect (OR=1.49 [1.37-1.63]; p=2.4×10^-19^) similar to that in EA only. Examining the risk by local ancestry, the EUR and the native American (NAT) tracts demonstrated a large effect size, whereas the AFR tract did not (OR=1.04 [0.78-1.39; p = 0.79]), consistent with the AFR tract in the AA population. However, the NAT tract had much smaller allele frequency (3% in NAT vs 38% in EUR haplotypes). Thus, most of the risk at *CFH* in the AA and HA GWAS arises from EUR haplotypes.

**Fig. 6.**
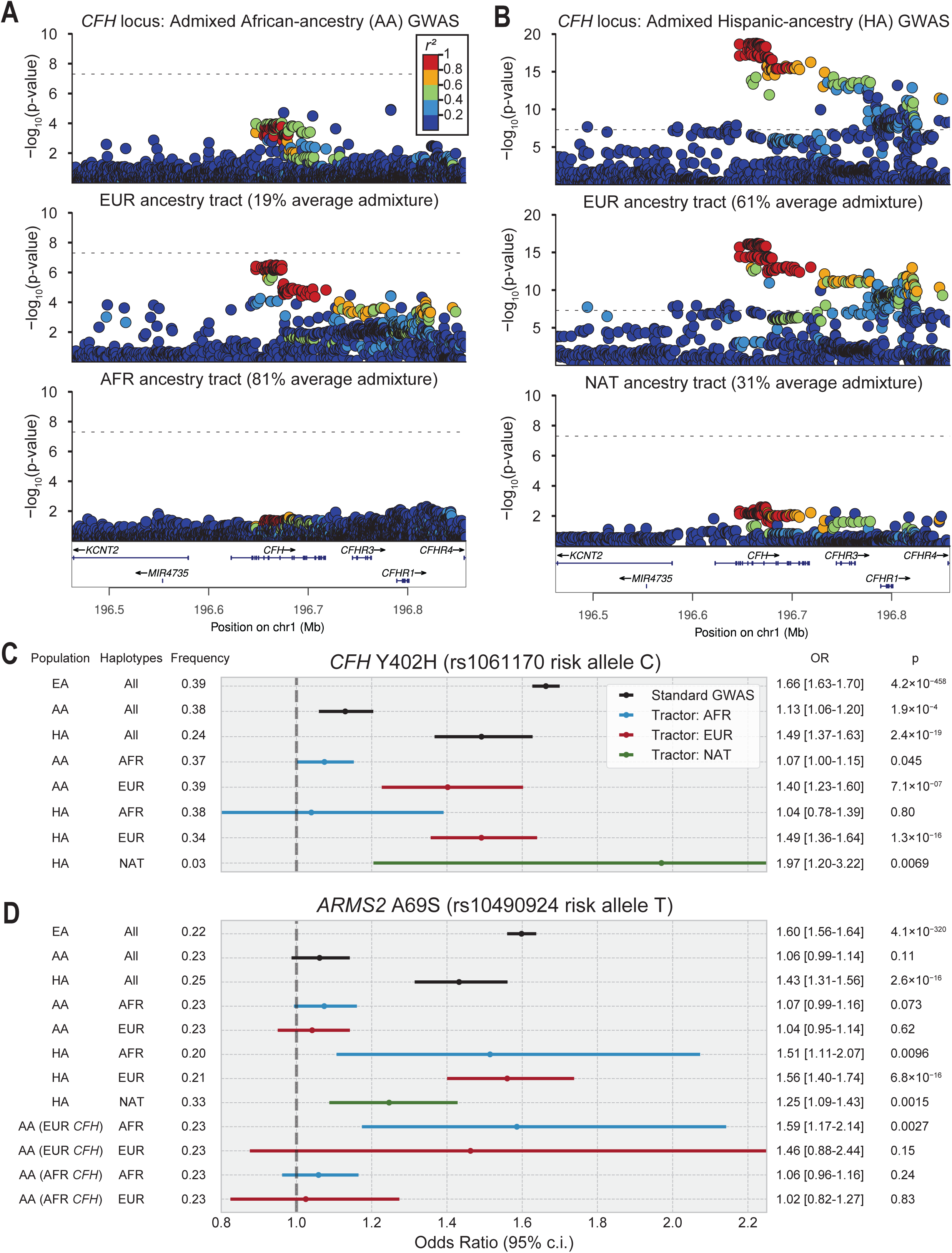
Local ancestry analysis of the *CFH* and *ARMS2/HTRA1* loci. (**A**) Regional association plots of AMD risk at *CFH* in a standard GWAS in AA individuals (top), and the EUR (middle) and AFR (bottom) tracts from the Tractor analysis. (**B**) Regional association plots of AMD risk at *CFH* in a standard GWAS in HA individuals (top), the EUR ancestry tract from the tractor analysis (middle) and the NAT ancestry tract (NAT) (bottom). (**C**) Forest plot comparing effect sizes and confidence intervals of the *CFH* Y402H risk allele in EA, AA, and HA populations, with Tractor analyses for AA (AFR and EUR tracts), and HA (EUR, NAT, and AFR tracts). (**D**) Forest plot comparing effect sizes and confidence intervals of the *ARMS2* A69S risk allele in EA, AA, and HA populations, with Tractor analyses for AA (AFR and EUR tracts), and HA (EUR, NAT, and AFR tracts). Additionally, Tractor effect size estimates in AA populations with either homozygous EUR ancestry at *CFH* (“EUR *CFH*”) or homozygous AFR ancestry at *CFH* (“AFR *CFH*”) are provided. Error bars are colored according to whether they correspond to a standard GWAS (black), or a Tractor GWAS, with blue for AFR tracts, red for EUR tracts, and green for NAT tracts.

Smaller *CFH* Y402H effect size observed in AFR could be explained through haplotype analysis. In particular, a protective deletion of *CFHR3* and *CFHR1* (*35*) has a higher frequency in AFR haplotypes (40% compared to 20% in EUR haplotypes) (*36*) and has been proposed as an explanation for lower rates of AMD observed in AA (*37*). As no SNPs tag the *CFHR3-CFHR1* deletion with r^2^ > 0.8 in AFR haplotypes, we called the deletion directly from genotype intensities and phased the deletion with the genotypes. We found that the deletion is largely out of phase with the *CFH* Y402H risk allele (D’=0.87) in AFR haplotypes; thus, it cannot fully explain the reduced effect size. Stratified by local ancestry, haplotypes in AA were analyzed using two models (*38, 39*), excluding SNPs that are rare in AFR haplotypes **(Table S19)**. While both models suggest that the putative risk haplotype may have a lower frequency in AFR compared to EUR, neither model demonstrated enrichment in cases with homozygous AFR local ancestry; they did, however, find haplotypes with enrichment in cases with homozygous EUR local ancestry. Thus, our analysis suggests a smaller effect size in AFR haplotypes, which may contribute to the lower incidence of AMD in AA; we acknowledge that the *CFH* locus comprises a complex genetic architecture (*40*).

### Genetic risk at *ARMS2* is lower in admixed Black subjects, but does not vary by haplotype ancestry

Similarly, we dissected the local ancestry contribution of genetic risk at the *ARMS2- HTRA1* locus, which also demonstrated marked cross-ancestry heterogeneity. In contrast to *CFH*, we did not find significant differences in the proportion of European haplotypes at *ARMS2* in admixed African or Hispanic subjects using Fisher’s Exact Test **(Table S20)**. We tested the *ARMS2* locus using the Tractor method (**Fig. 6d**). Within AA subjects, OR at the *ARMS2* A69S polymorphism were close to 1 and not significant in both the AFR and EUR tracts. However, within admixed HA subjects, EUR, NAT, and AFR local ancestry tracts surprisingly all showed similar statistically significant OR. We hypothesized that risk at *ARMS2* was partially dependent on genetic architecture at *CFH*, and isolated admixed AA subjects with EUR/EUR local ancestry at *CFH*. We observed OR in AFR and EUR tracts similar to the EA and HA GWAS, with the AFR tract reaching statistical significance (p=0.0026). In contrast, OR were close to 1 in both AFR and EUR tracts in subjects with homozygous AFR local ancestry at *CFH*. Thus, our results suggest that the causal variant at *ARMS2* has similar effect size across ancestries, and the lack of association in AA may be due to epistasis or pathway effects related to the conspicuous lack of complement loci in the AA GWAS. Supporting this hypothesis, we modeled epistatic interaction between the *CFH* and *ARMS2* risk alleles in MVP EA **(Table S21)** and observed a significant association of AMD with the interaction term (OR=1.11 [1.08-1.14]; p=1.6×10^-13^).

### An MHC Class II haplotype is associated with AMD in African and European ancestries

As the AA GWAS demonstrated a primary association with the MHC and not *CFH* and *ARMS2* as observed in other ancestries, we dissected the MHC signal by testing HLA alleles imputed with HIBAG (*41*). Full summary statistics are provided in **Tables S22-S24**. Within MVP EA (32,567 cases and 130,444 controls), we replicated HLA allele associations (*42*) with DQB*02:01 (p=1.2×10^-7^), DQB*02:02 (p=4.7×10^-10^), and DRB*03:01 (p=2.5×10^-7^); however, the DQB*02:01 and DRB*03:01 alleles were no longer significant in the fully adjusted model, especially after adjustment for the nearby *CFB/C2* SNP rs429608. Eight alleles were significant after Bonferroni correction, with four novel associations reaching genome-wide significance (A*11:01, B*14:01, DRB1*07:01, and DQA1*02:01). Within AA subjects (2,296 cases and 28,478 controls), three Class II alleles were highly significant: DRB1*07:01 (1.52 [1.37-1.68]; p=5.2×10^-15^), DQA1*02:01 (1.39 [1.26-1.52]; p=2.4×10^-11^), and DQB1*02:02 (1.39 [1.27-1.52]; p=2.1×10^-12^).

These three alleles are known to form a haplotype (HLA-DRB1*07:01-DQA1*02:01-DQB1*02:02) that is associated with autoimmune conditions such as celiac disease (*43*) and asparaginase hypersensitivity (*44, 45*). In an additive model, the risk haplotype was highly associated with AMD, with OR=1.51 [1.37-1.66; p=1.8×10^-16^] in AA and replicated with a smaller effect size, OR=1.16 [1.12-1.20; p=2.1×10^-15^], in EA, with all other haplotypes as reference. A PheWAS of the risk haplotype in AA led to significant enrichment of diagnostic signals for drusen (p=1.2×10^-26^) and dry AMD (p=4.7×10^-9^), which were corroborated in the EA analysis (p=6.7×10^-4^ and p=2.4×10^-3^, respectively). Additionally, we replicated the celiac disease association in the EA cohort (p=1.0×10^-12^). Antibodies directed against HLA-DR showed immunoreactivity in drusen (*46*), supporting biological plausibility.

### Multi-tissue TWAS of AMD identifies novel AMD-associated genes

To pair the GWAS signals with functional gene units, predictive models of gene expression (*47*) were used to perform a multi-tissue transcriptome-wide association study (TWAS), based on our EA discovery meta-analysis (N=45,045 cases, 275,806 controls). We performed TWAS using the high-powered dorsolateral prefrontal cortex (DLPFC) dataset from the PsychENCODE Consortium (PEC) (*48*) **(Table S25)**, as well as retina (*49*) **(Table S26)**, and 35 other tissue models built from GTEx v8 (*50*) and STARNET (*51*). We then meta-analyzed tissue p-values using ACAT (*52*) **(Table S27)**. AMD-related gene expression was highly correlated across tissues **(Fig. S9a)**. Consistent with the high protein and gene expression similarity among retina and brain (Human Protein Atlas, proteinatlas.org) (*53*), we found that the retina TWAS clustered together with the TWAS in DLPFC and other brain tissues **(Fig. S9b)**.

Applying the DLPFC model, 156 genes and 326 transcripts were significant at the FDR-corrected p<0.05 association threshold (**Fig. 3a**). Novel TWAS genes include regulatory genes for protein glycosylation (*MAN2C1*, *MPI*, *TMEM199*, and *CTSA*), lipid metabolism (*CHD9*), cell cycle (*FRK*), and neurogenesis and neural differentiation (*NTN5* and *NIF3L1*). Significant genes not at a genome-wide significant GWAS locus include *CAB39*, *CUL1* antisense transcript *AC005229.4* (ENSG00000273314), and *WAC-AS1* **(Fig. S10)**. Along with the novel GWAS locus near *CAND2*, *CUL1* and *WAC* are components of the ubiquitin-proteasome system, suggesting a larger role for that pathway in AMD than previously appreciated.

At *CD46/CD55* we observed significant downregulation of the *CD55-*203 transcript and upregulation of *CD46* overall, with differential regulation of some *CD46* transcripts (**Fig. 5c**), likely corresponding to a strong splice QTL at the index SNP (rs2724360) observed in many tissues in GTEx (p=1.3×10^-25^ in DFPLC). We further performed retina summary-based MR (SMR) experiments **(Table S28)**, which provide support for a causal link between AMD risk variants and expression of *CD46*, but not *CD55*.

We then conducted gene set enrichment analysis (GSEA) on significant TWAS genes in the DLPFC-based model **(Table S29; Fig. S11a)** and the tissue meta-analysis **(Table S30; Fig. S11b)**. In addition to complement cascade, humoral immunity, and HDL cholesterol regulation pathways, we observed significant enrichment in genes belonging to pathways related to regulation of immune and inflammatory responses, and death receptor-mediated apoptosis. We further performed semantic clustering of significant GO terms in the tissue meta-analysis GSEA, which identified two major themes associated with AMD pathology: immune functions and lipid homeostasis **(Fig. S12)**. Finally, we conducted gene-based rare variant burden analyses of the MVP EA cohort **(Table S31)**, which confirmed the risk-increasing effect of rare *CFH* and *CFI* mutations, and performed enrichment using our TWAS results **(Fig. S13)**. In addition to *CFH* and *CFI,* genes prioritized by this analysis include *CETP, ABCA7, ELP5* and *B3GLCT*.

## Discussion

We performed the largest GWAS meta-analysis to date and the first GWAS of AMD in AA and HA populations using data from veterans participating in MVP, nearly quadrupling the number of cases and doubling associated loci. Importantly, we uncovered marked differences in risk between major AMD loci across populations even at loci with known large effect sizes, especially *CFH* and *ARMS2/HTRA1*, which may partially explain lower rates of AMD in these groups. We submit that the overall reduction in risk in AA is due to tempered involvement of the complement pathway, substantiated through overall weak PRS transferability. In contrast to other ancestries, our AA GWAS had a primary peak at *HLA-DQB1*, corresponding to an MHC Class II risk haplotype which was replicated with a smaller effect size in EA subjects. This heterogeneity in genetic architecture may reflect the role of natural selection in shaping immune responses across ancestries (*54, 55*). However, our PRS-PheWAS experiment using the AA GWAS as a base, which demonstrated associations between the AA AMD PRS and increases in both HDL cholesterol and neutrophil counts, points to shared disease mechanisms.

We dissected cross-ancestry heterogeneity at the *CFH* and *ARMS2* loci using haplotype-based local ancestry analyses, “painting” the ancestral origin of each chromosome. These analyses revealed that AFR haplotypes have a smaller marginal effect size at the *CFH* risk allele, compared to EUR and NAT haplotypes in the same individuals, in both AA and HA populations. In contrast with *CFH*, the lack of association at *ARMS2* in AA is not due to ancestry-based haplotype-specific effects. In AA individuals, both AFR and EUR haplotypes demonstrated similarly small effect sizes. However, in HA individuals, which show high risk at *ARMS2*, AFR, EUR, and NAT haplotypes all showed similarly large marginal effect sizes. We infer from these observations a diminished role for the *ARMS2/HTRA1* locus in the absence of genetic risk at *CFH*. We further leveraged the statistical power offered by MVP to confirm a robust interaction effect between the *CFH* and *ARMS2* risk alleles in EA.

A general conclusion from years of investigative efforts is that the genetic architectures at the broader *CFH* and *ARMS2/HTRA1* loci are multifaceted (*40, 56*). At *CFH*, both common and rare variation (*8*) as well as structural variation (*35*) plays a role. At *ARMS2/HTRA1*, broad linkage disequilibrium between variants has made fine-mapping and identification of causal mechanisms difficult. Current literature points to a tissue-specific cis-regulatory element within *ARMS2* that is disrupted by the risk allele, leading to downregulation of *HTRA1* (*57*). HTRA1, a serine protease with reported roles in TGF-β signaling (*58*) and regulation of monocyte elimination (*59*), has also been shown to cleave neutrophil elastase inhibitor α-1 antitrypsin (*SERPINA1*) (*60*), producing NET inhibitory peptides, or NIPs (*19, 61*). NETs activate complement (*62–64*), and CFH itself may both regulate NETosis (*65*) and bind to NETs (*66*), potentially connecting the biology of the two principal AMD risk loci. Supporting this pathway, we discovered that two low-frequency pathogenic mutations in *SERPINA1* that cause α-1 antitrypsin deficiency are associated with AMD, but with opposite directions of effect. Additionally, consistent with histological (*67*) and observational (*68*) evidence, our AMD PRS-PheWAS experiments in both EA and AA showed association with increased neutrophil counts. Thus, integration of epidemiologic and genetic data from European and non-European populations synergizes and unifies evidence, and refines risk models.

Other loci to emerge in our GWAS meta-analysis continue to expand on known AMD themes, including numerous novel loci related to the complement cascade, lipid metabolism, angiogenesis, TGF-β signaling, photoreceptor function, apoptosis, and inflammation. We also identified the first pigmentation loci associated with AMD: *HERC2/OCA2*, *TYR*, and *TRPM1*. We explored this theme further by demonstrating for the first time a protective effect of pigmentation on AMD in EA using MR, supporting a causal relationship where long-standing observational evidence has been inconsistent (*69*).

To pinpoint causal changes in genetically regulated gene expression, we performed multi-tissue TWAS and SMR experiments. Our analysis adds transcriptomic imputation models from new cohorts (PsychENCODE and STARNET), expands sample sizes in GTEx (GTEx v8 vs. GTEx v7), leverages a higher-powered GWAS, and introduces expression information at the transcript level in the DLPFC model. A comparison with previous TWAS studies (*49, 70*) is summarized in **Table S32**. TWAS enabled us to refine several loci, such as at *CD46/CD55*, where we identified specific *CD46* transcripts associated with AMD. Our TWAS analysis also enabled us to identify numerous additional novel AMD-associated genes across several pathways, including ubiquitin-proteasome system, protein glycosylation, cell cycle regulation, and neural differentiation.

While our analysis demonstrated a very high genetic correlation with IAMDGC (95%), one limitation is that our phenotyping approach used only ICD code-based diagnoses and demographic parameters (*71*), which limited our ability for deep phenotyping. An important future direction will be to review ophthalmologic imaging data to assess disease severity and pathology in patients diagnosed with AMD and associate findings with specific genetic risk factors, such as the HLA risk haplotype in AA. To date, most large-scale imaging studies of AMD have focused on EA populations, and as our results demonstrate, increasing diversity is important.

In conclusion, our study expands the number of genetic loci associated with AMD and fills in important gaps in the literature regarding the genetics of AMD in non-European ancestries. Moreover, it illustrates the importance of considering diverse admixed genomes and using local ancestry-aware analyses (*34*), which can leverage differences in effect size, frequency, and LD to tease apart mechanisms of genetic risk and improve risk prediction.

## Materials and Methods

### Samples

A general description of the MVP sample may be found in Gaziano et al. (*9*). Composition of the cohorts studied here is available in **Table S1.** Informed consent was obtained from all participants, and all studies were performed with approval from the Institutional Review Boards at participating centers, in accordance with the Declaration of Helsinki.

### MVP

We used ICD9/ICD10 codes in VA electronic health records to define AMD case/control status, using Algorithm 4 in Halladay et al. (*71*) in which AMD cases and controls were required to be 50 and 65 years of age, respectively. Briefly, AMD cases had ICD9/ICD10 codes indicating AMD from at least two separate eye clinic visits; controls had either no eye clinic visits or no AMD diagnoses. The three EA cohorts represent three separate releases of the MVP genotype data.

A description of the custom MVP Affymetrix Biobank chip, and general quality control (QC) procedures for genetic data, have been published (*72*). Briefly, the MVP sample was genotyped for a custom Affymetrix Biobank chip, containing 725,000 variants. Samples were imputed to the 1000 Genomes Phase 3 panel. AA and HA samples were additionally imputed to the African Genome Resources imputation panel from the Sanger Institute (*73*) to improve imputation of African haplotypes. Imputed genotypes were available for a total of 658,000 Veterans across all ethnic groups. We used HARE categories (*74*) AFR (African American), EUR (European American) and HIS (Hispanic American) to harmonize genetic ancestry with self-identified race/ethnicity. Principal components for ancestry were determined within HARE groups.

### Genentech

#### Study Design and Populations

We performed whole-genome sequencing of DNA derived from blood samples obtained from patients with choroidal neovascularization (CNV) participating in clinical trials for Ranibizumab (NCT00891735 [HARBOR], NCT00061594 [ANCHOR] and NCT00056836 [MARINA]) and from patients with geographic atrophy (GA) participating in clinical trials for Lampalizumab (NCT02247479 [CHROMA], NCT02247531 [SPECTRI], NCT01229215 [MAHALO]) and an observational study (NCT02479386 [PROXIMA]). Patients had to consent for genetic analysis for inclusion eligibility and these cohorts were selected for inclusion based on available phenotypic information and DNA availability for whole-genome sequencing.

Two independent sets of non-AMD controls were used for the GA risk analysis and the CNV risk analysis. Samples and data for non-AMD controls without CNV were obtained from clinical trial cohorts of asthma and RA. Samples and data for non-AMD controls without GA were obtained from clinical trial cohorts of asthma, colorectal cancer, COPD, inflammatory bowel disease, IPF and RA. No healthy controls were used because no whole-genome sequencing data was available. All non-AMD controls were aged 50 or older and of European ancestry.

All patients (AMD and non-AMD controls) included in this study provided written informed consent for whole-genome sequencing of their DNA. Ethical approval was provided as per the original clinical trials.

#### DNA analysis

The whole-genome sequencing data was generated to a read depth of 30X using the HiSeq platform (Illumina X10, San Diego, CA, USA) processed using the Burrows-Wheeler Aligner (BWA) / Genome Analysis Toolkit (GATK) best practices pipeline. Whole genome sequencing short reads were mapped to hg38 / GRCh38 (GCA_000001405.15), including alternate assemblies, using BWA version 0.7.9a-r786 to generate BAM files. All sequencing data was subject to quality control and was checked for concordance with SNP fingerprint data collected before sequencing. After filtering for genotypes with a GATK genotype quality greater than 90, samples with heterozygote concordance with SNP chip data of less than 75% were removed. Sample contamination was determined with VerifyBamID software. And samples with a freemix parameter of more than 0.03 were excluded. Joint variant calling was done using the GATK best practices joint genotyping pipeline to generate a single variant call format (VCF) file. The called variants were then processed using ASDPEx to filter out spurious variant calls in the alternate regions.

#### Quality Control

Samples were then excluded if the call rate was less than 90%. Identity by descent analysis was used to detect and filter out relatedness in the dataset; samples were excluded if PI_HAT was 0.4 or higher. Samples were removed if they showed excess heterozygosity with more than three standard deviations of the mean. This resulted in 1,703 GA patients with 2,611 non-GA controls, and 1,175 CNV patients with 3,225 non-CNV controls.

Sample genotypes were set to missing if the Genotype Quality score was less than 20 and SNPs were removed if the missingness was higher than 5%. SNPs were filtered if the significance level for the Hardy-Weinberg equilibrium test was less than 5×10^-8^. The allele depth balance test was performed to test for equal allele depth at heterozygote carriers using a binomial test; SNPs were excluded if the p-value was less than 1×10^-5^.

### GWAS

#### MVP EA

GWAS on the first two EA tranches of the MVP cohort were conducted in PLINK 2.0, adjusting for sex and six ancestry-specific principal components.

#### MVP AA and HA

Individuals were classified by genetic ancestry using the HARE method (*74*) which harmonizes self-reported ethnicity with genetic ancestry. We conducted GWAS in MVP AA and HA populations with REGENIE v1.6.7 (*75*) using sex and 10 ancestry-specific PCs as covariates. We used the approximate Firth mode (--firth --approx) with a p-value threshold of 0.05 to control type I error rate in rare SNPs. We additionally used option --firth-se to generate standard errors computed from the Firth effect size and likelihood ratio test p-value for meta-analysis purposes.

#### MVP EA replication GWAS

We conducted a GWAS in a third tranche of MVP data that later became available to us. To ensure that novel associations in the EA meta-analysis were robust to population structure, we conducted the EA replication GWAS in REGENIE v2.2.4 (*75*) using sex and ten ancestry-specific PCs as covariates. We used the approximate Firth mode (-- firth --approx) with a p-value threshold of 0.05 to control type I error rate in rare SNPs. We additionally used option --firth-se to generate standard errors computed from the Firth effect size and likelihood ratio test p-value for meta-analysis purposes.

#### IAMDGC

Phenotyping and GWAS analysis in IAMDGC is described in detail elsewhere (*8*). Briefly, European-ancestry subjects were genotyped on a custom Illumina HumanCoreExome array and imputed to the 1000 Genomes Phase 1 panel. GWAS analyses were conducted using EPACTS using DNA type (whether whole genome-amplified or not) and two PCs as covariates.

#### GERA

Phenotyping and sample collection in GERA are described in detail elsewhere (*42, 76*). Cases and controls were selected from European-ancestry subjects. Samples were genotyped on an Affymetrix Axiom array and imputed to the 1000 Genomes panel. GWAS were conducted in PLINK 1.07 using age, gender, PCs 1-10 (calculated within European-ancestry subjects), and percentage of Ashkenazi ancestry as covariates.

#### UK Biobank

Cases and controls were selected from subjects of at least 65 years of age. Samples were genotyped on the UK BiLEVE Axiom Array or the UK Biobank Axiom Array and imputed to the Haplotype Reference Consortium (HRC) reference panel. GWAS analyses were conducted in PLINK 1.9 using sex and the first two PCs as covariates.

#### Genentech GA and CNV

Common variant (MAF>=1%) genome-wide association studies (GWAS) were conducted to separately assess GA risk and CNV risk as compared to independent non-AMD control cohorts. PLINK was used to perform logistic regression using an additive model, adjusting for age, sex and the first three principal components.

### Statistical Analysis

#### Epidemiological analyses

Observational association analyses were conducted using the statsmodels (version 0.12.2) Python module. Case-control analyses were performed with logistic regression. Time-to-AMD-diagnosis analysis was performed using a Cox proportional hazards model. The index date was defined as the date of first recorded visit to the VA healthcare system. An event was defined as a subject receiving AMD diagnoses on two unique days. The time-to-event was defined as the time from the first recorded visit to the first recorded AMD diagnosis. Subjects with fewer than two unique days with AMD diagnoses were right-censored by the date of the last recorded VA healthcare system visit.

#### GWAS Meta-analysis

GWAS meta-analyses were conducted using the inverse variance-weighted fixed effects scheme as implemented in METAL (*77*).

#### Additional QC

We imposed the following criteria for the GWAS and meta-analysis: imputation quality score ≥ 0.5, minor allele frequency ≥ 0.005, and *p* ≥ 1×10^-50^ from an exact test for Hardy-Weinberg proportions.

#### Locus definition

Association loci were defined using the FUMA GWAS online tool (*78*) with default options (merging together LD blocks closer than 250 kb into a locus).

#### IAMDGC-based polygenic risk score analysis

We derived AMD PRS scores for subjects in MVP across all ancestries using the IAMDGC GWAS (*8*) as a base. Weights were derived from the base GWAS using PRS-CS (*10*) with a global shrinkage prior of 1×10^-4^ (the default recommended for less-polygenic traits), using the UK Biobank European-ancestry LD reference. Imputed SNPs meeting R^2^ > 0.8 and minor allele frequency > 1% QC criteria were included in the model. Scores were then normalized to zero mean and unit variance within each ancestry group. *AA GWAS-based polygenic risk score analysis.* To better understand the pleiotropy of AMD genetic risk in African ancestries, we used the AA summary statistics as the base for scoring polygenic risk in unrelated MVP AA subjects that were not included as cases or controls in the GWAS cohort. As our phenotyping approach required subjects to have visited a VA eye clinic and been evaluated by an eye specialist, the PRS was evaluated in the remaining unrelated AA subjects with no such history. Weights were derived from the base MVP AA GWAS using PRS-CS (*10*) with a global shrinkage prior of 1×10^-4^ (the default recommended for less-polygenic traits), using the UK Biobank African-ancestry LD reference. Scores were then normalized to zero mean and unit variance.

#### Conditional association analysis

We determined a set of mutually independent genome-wide significant variants using the conditional analysis procedure implemented in GCTA-COJO (*79*). A subset of 100,000 MVP EA individuals (as classified by the HARE method) formed the reference population for LD structure. Independent significant variants were chosen over all loci simultaneously by means of the --cojo-slct procedure for stepwise regression. Selected variants had association *p* < 5×10^-8^ while adjusting for all other variants and were not necessarily pairwise independent.

#### HLA analysis

Four-digit HLA alleles were imputed in all subjects in MVP using HIBAG (*41*) with the Axiom UK Biobank Array multi-ethnic model as reference. Allele calls with posterior probability less than 50% were filtered out. Alleles were tested additively relative to all other alleles at each locus using the logistf R package for Firth logistic regression (*80*). Two models were applied: the ‘standard’ model with adjustment for sex and PCs 1-10, and the ‘fully adjusted’ model with adjustment for sex, PCs 1-10, and major AMD risk alleles at *CFH*, *ARMS2*, *CFB/C2*, and *C3*. Alleles with a minor allele count less than 50 were not tested. P-values and confidence intervals are based on the penalized likelihood ratio test and profile penalized likelihood, while the standard error and z-score are based on the Wald method.

#### HLA-DRB1*07:01-DQA1*02:01-DQB1*02:02 haplotype analysis

Two SNPs (rs28383172 and rs7775228) were recently reported to tag the haplotype with high sensitivity and specificity (*81*), both of which are genotyped on the MVP array. We phased genotypes using SHAPEIT4 (*82*) and designated the risk haplotypes as those with G and C alleles, respectively.

#### Local ancestry analysis

We inferred local ancestry within our AA sample assuming two-way (AFR/EUR) admixture, and within our HA sample assuming three-way (AFR/EUR/NAT) admixture. The 1000 Genomes YRI (N=108) and CEU (N=99) populations, were used as the AFR and EUR reference, respectively, and 43 Native American samples from Mao et al. (*83, 84*) were used as the NAT reference. We used RFMIX version 2 (*33*) to generate local ancestry calls for phased genotypes. We then extracted ancestry-specific dosages from the imputed data into PLINK 2.0-compatible files (*85*) using custom scripts based on the Tractor workflow (*34*). For the AA analysis, EUR-specific dosages were put into a PGEN file, and AFR-specific dosages and EUR haplotype counts were interlaced in a zstandard-compressed table. For the HA analysis, EUR-specific dosages were put into a PGEN file, with AFR and NAT-specific dosages and EUR and AFR haplotype counts interlaced into a zstandard-compressed table. We used these files to conduct a local ancestry-aware GWAS using the PLINK 2.0 local covariates feature, obtaining ancestry-specific marginal effect size estimates.

#### CFHR3-CFHR1 deletion calling

We calculated log-R ratio intensities for 12 probesets spanning the *CFHR3* and *CFHR1* deletion. We then clustered intensities by constructing a two-dimensional UMAP embedding (*86*), which separated samples into 0, 1, and 2-copy number clusters. In order to conduct haplotype analyses, the deletion was phased with the genotype data using SHAPEIT 4.2 (*82*), and the locus was re-imputed using Minimac4 (*87*) to obtain accurate deletion calls alongside imputed genotypes.

#### Rare variant gene-based analysis

We conducted gene-based rare variant burden association analyses of the entire MVP EA cohort (N=34,046 cases, 135,775 controls). We considered only variants genotyped on the MVP 1.0 array (*72*), which is enriched in protein-altering rare variants, and applied a recent technical advance (*88*) that improves the positive predictive value of rare genotype calls. We further restricted the included markers to ultra-rare variants (MAF < 0.25% in controls) classified as “high-impact” (*89*). We then defined a series of three “masks” over which the burden test was performed: mask 1 including frameshift and nonsense loss of function (LoF) variants only, mask 2 including LoF + missense variants, and mask 3 including LoF + missense + splice site variants. Burden test association analyses were conducted in REGENIE v2.02 with Firth logistic regression (*75*).

#### Relatedness

For experiments in unrelated samples, we used KING version 2.2.7 (*90*) to generate a maximal set of samples that are unrelated within two degrees.

#### Summary-based MR

We performed summary-based MR experiments on retina eQTL data from EyeGEx using the Summary-based Mendelian Randomization (SMR) software tool (*91*).

#### Two-sample MR

We performed MR on selected pigmentation traits and AMD using the TwoSampleMR R package (*92*). Instruments were derived from genome-wide significant loci reported in the exposure trait GWASs and clumped using the TwoSampleMR clump_data tool with default options (*r^2^*< 0.001) to ensure no correlation between instruments.

#### Genetic correlation

Genetic correlation analyses were performed using LDSC 1.01 (*11*) using the provided European-ancestry LD scores derived from the 1000 Genomes project.

#### Fine-mapping

We performed Bayesian fine-mapping of each genome-wide significant locus in the European-ancestry meta-analysis and trans-ancestry meta-analysis using FINEMAP 1.4 (*30*). Pairwise SNP correlations were calculated directly from imputed dosages on 320,831 European-ancestry samples in MVP using LDSTORE 2.0. The maximum number of allowed causal SNPs at each locus was set to 10 (the default used in the FinnGen fine-mapping pipeline: https://github.com/FINNGEN/finemapping-pipeline). Finally, we performed multivariate multiple QTL (mmQTL) analysis as previously described (*31*), leveraging the multi-ancestry RNA-seq-based brain gene expression reference dataset derived from 2,119 donors (*31*) and the GWAS summary statistics from the AMD multi-ancestry discovery meta-analysis.

#### PheWAS

We performed PheWAS and LabWAS in unrelated subjects as implemented in the R PheWAS package version 0.12 (*93*). For each ICD code, cases were defined as at least 2 counts of the code on separate days, and controls were 0 counts. Subjects with 1 count were excluded. Additionally, we applied phenotype-based control exclusion (i.e., exclusion of controls that were cases in similar phenotypes) and we excluded females in male-specific phenotypes and vice versa. After this process, only phenotypes with 100 or more cases were considered. For labs, we used the median value in regression analyses, and required that each individual have at least two lab measurements to be included. Labs with measurements in at least 50 individuals were considered. Lab measurements were then normalized using inverse rank normal transformation (IRNT). All ICD code-based phenotypes were tested with logistic regression, at the Bonferroni-corrected p-value of 3×10^-5^. All 69 lab-based phenotypes were tested with linear regression, at Bonferroni-corrected p-value of 7×10^-4^. Both analyses adjusted for age, age-squared, sex (for non-sex specific phenotypes), and 20 ancestry-specific PCs.

### TWAS

#### Transcriptomic imputation model construction

Transcriptomic imputation models are constructed as previously described (*47, 94*) for tissues of the GTEx v8 (*50*), STARNET (*51*) and PsychENCODE (*48, 95*) cohorts. For GTEx and STARNET cohorts, we consider adipose tissue: subcutaneous (GTEx & STARNET) and visceral (GTEx & STARNET); arterial tissue: aorta (GTEx & STARNET), coronary artery (GTEx) and mammary artery (STARNET); Blood (GTEx & STARNET); brain (GTEx): anterior cingulate cortex, frontal pole cortex, hippocampus and substantia nigra; adrenal gland (GTEx); colon (GTEx): sigmoid and trasverse; esophagus (GTEx): gastroesophageal junction, mucosa and mascularis; pancreas (GTEx); stomach (GTEx); terminal ileum (GTEx); heart (GTEx): atrial appendage and left ventricle; liver (GTEx & STARNET), skeletal muscle (GTEx & STARNET); mammary tissue (GTEx); ovary (GTEx); lung (GTEx); skin (GTEx): not sun and sun exposed; spleen (GTEx). The genetic datasets of the GTEx (*50*), STARNET (*51*) and PsychENCODE (*95*) cohorts are uniformly processed for quality control (QC) steps before genotype imputation as previously described (*47, 94*). We restrict our analysis to samples with European ancestry as previously described (*47*). Genotypes are imputed using the University of Michigan server (*87*) with the Haplotype Reference Consortium (HRC) reference panel (*73*). Gene expression information is derived from RNA-seq gene level counts which are adjusted for known and hidden confounders, followed by quantile normalization. For GTEx, we use publicly available, quality-controlled, gene expression datasets from the GTEx consortium (http://www.gtexportal.org/). RNA-seq data for STARNET were obtained in the form of residualized gene counts from a previously published study (*51*). For the dorsolateral prefrontal cortex from PsychENCODE we used post-quality-control RNA-seq data that were fully processed, filtered, normalized, and extensively corrected for all known biological and technical covariates except the diagnosis status (*48*) as previously described (*94*). Finally, we constructed a retinal transcriptomic imputation model based on reference data from Ratnapriya et al. (*49*) comprising imputed genotypes and expression data for 406 individuals. Genotypes and gene expression data were subjected to additional quality control steps as described below. For population classification we used individuals of known ancestry from 1000 Genomes. We excluded variants in regions of high linkage disequilibrium, variants with minor allele frequency < 0.05, variant with high missingness (>0.01), and variants with Hardy-Weinberg equilibrium p < 1×10^-10^; the remaining variants were pruned (--indep-pairwise 1000 10 0.02 with PLINK (*96*)) and PCA was performed with PLINK 2 (*85*). We used the first (PC1), second (PC2) and third (PC3) ancestral principal components to define an ellipsoid based on 1000Gp3v5 EUR samples (*97*) and samples within 3 SD from the ellipsoid center were classified as EUR; based on this definition of EUR samples, we excluded one non-European ancestry individual. In the remaining samples (n = 405), we performed additional sample-level quality control by retaining non-related samples (--king-cutoff 0.0884 with PLINK 2 (*85*)) with sample-level missingness < 0.015 for variants with variant-level missingness < 0.02, and heterozygosity rate of < 3SD away from the mean; of note, no samples were excluded by these steps. For the next step of our pipeline, we performed outlier testing in the gene expression data; we performed PCA on raw gene counts and excluded individuals located more than 4 SD away from the mean of the ellipsoid defined by PC1 to PC3. The latter led to the exclusion of twelve samples, giving us a final sample size of 393 individuals for the training of the model. In this final set of individuals, we performed variant-level quality control of the genotypes by removing variants with greater than 0.05 minor allele frequency and 0.02 missingness rate; only variants present in the reference panel of the Haplotype Reference Consortium were retained to ensure good representation of variants in the target GWAS (*73*). After conversion of genotypes to dosages, missing values were replaced with twice the variant’s minor allele frequency. Expression data corresponding to the retained individuals were prepared as follows: genes with > 0.5 counts per million in at least 30% of samples were retained, expression values were normalized by voom (*98*), residualized for 10 PEER factors (*99*) and quantile normalized. For training, we used PrediXcan (*100*) for the construction of the retinal transcriptomic imputation model due to a lack of SNP epigenetic annotation information; for all other models, we used EpiXcan (*47*).

#### Multi-tissue transcriptome-wide association study (TWAS)

We performed the gene-trait association analysis as previously described (*47*). Briefly, we applied the S-PrediXcan method (*101*) to integrate the summary statistics and the transcriptomic imputation models constructed above to obtain gene-level association results. Results were corrected for multiple testing with the Benjamini & Hochberg (FDR) method (*102*). P-values across tissues were meta-analyzed using ACAT (*52*) ≤ 0.05 and predictive r^2^ > 0.01 (to control both for significance and variance explained).

#### Gene set enrichment analysis for TWAS results

To investigate whether the genes associated with AMD exhibit enrichment for biological pathways, we use gene sets from MsigDB 5.1 (*103*) and filter out non-protein coding genes, genes located at MHC as well as genes whose expression cannot be reliably imputed. In addition, we assay enrichment for genesets deriving from the rare variant analysis as above. Statistical significance is evaluated with one-sided Fisher’s exact test and the corrected p-values are obtained by the Benjamini-Hochberg (FDR) method (*102*). Semantic enrichment analysis of the TWAS results was performed as follows: a) gene ontology (GO) data (*104, 105*) were retrieved on 2021-07-10; b) we run Fisher-based gene set enrichment analysis as above and kept enrichments with FDR-corrected p-value ≤ 0.1; c) we used GOGO (*106*) to generate the directed acyclic graphs (DAGs) and estimated all possible pairwise semantic similarities when both GO terms of each pair fell within one of the three DAG domains: molecular function ontology (MFO), biological process ontology (BPO), and cellular component ontology (CCO); d) to characterize/illustrate the semantic clusters deriving from the significantly enriched gene sets, we converted semantic similarity between GO terms to distance and determined the optimal number of clusters with NbClust (*107*) using the ward.D2 method (*108*) according to the Silhouette criterion (*109*); e) finally, we built a word cloud for each semantic cluster by mining text terms (*110*) from its members’ GO term titles.

## Data Availability

The full summary level association data from the individual population association analyses in MVP will be available via the dbGaP study accession number phs001672.

## Acknowledgements

The unsung heroes of this publication are the U.S. Veterans who participated in this study, and who contributed time and effort to improve the lives of all Veterans and of populations worldwide. Key individuals who collected, curated or cleaned data for the Million Veteran Program are listed in the Supplementary Information. Dr. Igo passed away during the drafting of this manuscript and is remembered for his tireless contributions. We are further grateful to Catherine Tcheandjieu and Elizabeth Atkinson for advice on local ancestry inference and Tractor implementation, and to Jonathan Haines for comments on an earlier version of the manuscript. Finally, we wish to acknowledge the UK Biobank Eye and Vision Consortium who helped collect the eye phenotypes in the UK Biobank. The UK Biobank Eye and Vision Consortium is supported by grants from Moorfields Eye Charity, The NIHR Biomedical Research Centre at Moorfields Eye Hospital NHS Foundation Trust and UCL Institute of Ophthalmology, the Alcon Research Institute and the International Glaucoma Association (UK). This research has been conducted using the UK Biobank Resource under Application Number 2112. This project was supported by the Clinical and Translational Science Collaborative of Cleveland, which is funded by the National Institutes of Health, National Center for Advancing Translational Science, Clinical and Translational Science Award grant UL1TR002548. The content of this paper is solely the responsibility of the authors and does not necessarily represent the official views of the Department of Veterans Affairs or any agency of the United States federal government.

