## Supplementary Figures&legends and Table captions for "Distinctive cross-ancestry genetic architecture for age-related macular degeneration"

**Legends for Supplemental Figures**

**Fig. S1. Detailed study overview.** (**A**) PRS-PheWAS and PRS-LabWAS analyses using the external European-ancestry IAMDGC GWAS of AMD (*8*) as a base for scoring subjects in MVP. (**B**) Ancestry-specific GWAS analyses, and multi-ancestry meta-analysis, with corresponding secondary analyses including TWAS and Mendelian randomization (EA-only), and fine-mapping (EA and trans-ancestry). An unrelated MVP EA tranche was used as a replication cohort, and meta-analyzed into final EA and multi-ancestry meta-analyses. (**C**) HLA fine-mapping and rare variant analysis.

**Fig. S2. PRS distribution of AMD cases and controls.** (**A**) EA samples in MVP; (**B**) AA samples in MVP; and (**C**) HA samples in MVP. The PRS was constructed using weights derived from the IAMDGC GWAS (*8*).

**Fig. S3. Regional association plots of novel loci from the European-ancestry replication meta-analysis (57,290 cases, 324,430 controls).** The 1000 Genomes EUR panel was used as the reference for plotting LD $r^2$.

**Fig. S4. QQ plots of the multi-ancestry GWAS and meta-analysis.** (**A**) MVP AA GWAS, (**B**) MVP HA GWAS, (**C**) EA meta-analysis, and (**D**) multi-ancestry meta-analysis. SNPs with minor allele frequency greater than 0.5% are plotted and included in the genomic control inflation calculation.

**Fig. S5. Conditional analysis of *PLTP* and *MMP9* signals in the 20q13 locus.** Regional association plots of: (**A**) the EA replication meta-analysis, demonstrating a primary signal near *PLTP*; (**B**) the IAMDGC study (*8*), demonstrating a primary signal near *MMP9*; (**C**) the EA meta-analysis conditioned on the index SNP from the IAMDGC study (rs1888235); (**D**) the IAMDGC study conditioned on the index SNP from the EA meta-analysis (rs17447545). LD $r^2$ values were calculated in the MVP EA cohort. The dashed line denotes genome-wide significance ($5 \times 10^{-8}$).

**Fig. S6. Genome-wide significant loci from the AA and HA GWAS in MVP.** (**A**) Regional association plot of the *HLA-DQB1* locus from the AA GWAS (2,302 cases, 29,223 controls). The 1000 Genomes AFR panel was used as the reference for plotting LD $r^2$. (**B-C**) Regional association plots of the *CFH* and *ARMS2-HTRA1* loci from the HA GWAS (1,656 cases, 10,819 controls). The 1000 Genomes AMR panel was used as the reference for plotting LD $r^2$.

**Fig. S7. Novel genome-wide significant loci from the multi-ancestry meta-analysis.** Regional association plots of novel loci from the multi-ancestry replication meta-analysis (61,258 cases, 364,472 controls) beyond that reported for the EA meta-analysis. The 1000 Genomes EUR panel was used as the reference for plotting LD $r^2$.

**Fig. S8. Co-localization of GWAS and eQTL signals identifies candidate causal variants.** Shown are significant co-localized signals (co-localization posterior probability > 0.01) from the multi-ancestry discovery meta-analysis using the mmQTL (*31*) method.

**Fig. S9. Characterization of TWAS results across tissues.** (**A**) Pearson correlation of AMD association z-scores across the 38 tissues analyzed from GTEx, EyeGEx, STARNET, and PEC. (**B**) Hierarchical clustering of tissues based on AMD associations using Ward's linkage method.

**Fig. S10. Regional Miami plots of genes with novel TWAS associations.** The TWAS was based on the DLPFC brain model jointly testing genes and transcript isoforms. Results are based on the EA discovery meta-analysis (45,045 cases, 275,806 controls). The GWAS p-values are shown above the y=0 line, and the TWAS p-values are shown below it. SNPs are colored according to linkage disequilibrium r-squared with the lead SNP (EA panel). Lines connecting SNP predictors of genetically regulated gene expression are colored according to the significance level of the SNP: orange, $p \leq 5\times10^{-8}$; cyan, $p \leq 1\times10^{-5}$; gray, $p > 1\times10^{-5}$.

**Fig. S11. Gene set enrichment analysis (GSEA) of TWAS genes.** (**A**) GSEA of TWAS using the DLPFC transcripts (genes and isoforms) model, and (**B**) the multi-tissue meta-analysis. Shown are the top ten enrichments by effect size (odds ratio) in both analyses.

**Fig. S12. Semantic clustering of enriched gene ontology (GO) terms in the AMD multi-tissue transcriptome-wide association study (TWAS) identifies immune and lipid homeostasis functions.** First, we performed enrichment of AMD TWAS associated genes (FDR < 0.05 in ACAT meta-analysis of all tissues) for GO terms corresponding to ontologies of molecular functions (MFO), biological processes (BPO), and cellular components (CCO). Semantic clustering of the GO terms that had enrichment FDR < 0.1 (Fisher's exact test) and could be clustered identified 7 clusters (BPO1-BPO5, CCO1, and MFO1). For each of these clusters we provide a word cloud based on the description of the GO terms where words that appear more frequently are larger; clusters are ordered from left to right and top to bottom from the most significant to the least significant based on their meta-analyzed p-value (ACAT). Major themes that are associated with AMD are immune functions and lipid homeostasis.

**Fig. S13. Enrichment of rare variant burden associations in TWAS meta-analysis genes.** TWAS meta-analysis genes were those with FDR < 0.05; only protein-coding genes were considered. *** FDR < 0.001, ** FDR < 0.01, * FDR < 0.05 (12 rare variant gene sets were considered from the rare variant analysis). Mask 1: Loss of function mutations only; Mask 2: Loss of function + missense mutations; Mask 3: Loss of function + missense + splice mutations.

**Captions for Supplemental Tables**

**Table S1.** Sample demographics for the GWAS cohorts comprising the European-ancestry meta-analysis and multi-ancestry meta-analysis.

**Table S2**. (**A**) Effects of demographic parameters on AMD case/control status in the MVP EA, AA and HA samples, estimated in single regression models. (**B**) Effects of demographic parameters on AMD case/control status in the MVP EA, AA and HA samples, estimated in multiple regression models. (**C**) Multi-ancestry time-to-AMD-diagnosis analysis using a Cox proportional-hazards model.

**Table S3**. PheWAS summary statistics of the IAMDGC-derived PRS in European-ancestry subjects in MVP. The PRS was normalized to the unit normal distribution.

**Table S4**. LabWAS summary statistics of the IAMDGC-derived PRS in European-ancestry subjects in MVP. The PRS was normalized to the unit normal distribution and labs were inverse rank normal transformed (IRNT).

**Table S5.** Index SNPs from genome-wide significant loci in the European-ancestry replication meta-analysis (N=57,290 cases, 324,430 controls).

**Table S6.** Comparison of effect sizes at the index SNPs across the IAMDGC, GERA, MVP Replication GWAS.

**Table S7.** Conditionally independent genome-wide significant markers from the European replication meta-analysis.

**Table S8.** Genetic correlation of AMD with selected eye-related traits.

**Table S9.** Genetic correlation of AMD with blood-based biomarkers, hematological parameters, and other traits. Trait GWAS were from the Pan-UKB project unless noted otherwise. Genetic correlation analysis was conducted separately in the EA meta-analysis and EA replication GWAS, and rG effect sizes were meta-analyzed using inverse variance-weighted fixed effects meta-analysis.

**Table S10.** Mendelian randomization analyses of pigmentation traits on AMD. Traits were tested using five methods implemented in the TwoSample MR package: Inverse variance weighted, MR Egger, simple mode, weighted mode, and weighted median. Horizontal pleiotropy was tested via the MR Egger intercept.

**Table S11.** Index SNPs from genome-wide significant loci in the African-ancestry GWAS (N=2,302 cases, 1,656 controls).

**Table S12.** LabWAS summary statistics of the MVP AA GWAS-derived PRS in unrelated African-ancestry subjects in MVP not used in the GWAS cohort. The PRS was normalized to the unit normal distribution and labs were inverse rank normal transformed (IRNT). A PheWAS was also performed, with no significant associations.

**Table S13.** Index SNPs from genome-wide significant loci in the Hispanic/Latino-ancestry GWAS (N=1,656 cases, 10,819 controls).

**Table S14.** Index SNPs from genome-wide significant loci in the multi-ancestry replication meta-analysis (N=61,248 cases, 364,472 controls).

**Table S15.** SNPs with posterior probability > 0.01 from fine-mapping the European-ancestry discovery meta-1analysis (N=45,045 cases, 275,806 controls). Fine-mapping was conducted at each genome-wide significant locus using FINEMAP (*30*) software.

**Table S16.** SNPs with posterior probability > 0.01 from fine-mapping the multi-ancestry discovery meta-analysis (N=49,003 cases, 315,848 controls). Fine-mapping was conducted at each genome-wide significant locus using FINEMAP (*30*) software.

**Table S17.** Candidate causal variants from eQTL co-localization analysis using the mmQTL method (*31*). Shown are SNPs with co-localization posterior probability (CLPP) greater than 0.01. a) eQTL co-localization with the trans-ancestry fine-mapping. b) eQTL co-localization with the European-ancestry fine-mapping.

**Table S18.** Local ancestry analysis of haplotypes at *CFH* in admixed MVP populations. a) Enrichment of European-ancestry haplotypes (EUR) in AA AMD cases. b) Enrichment of individuals with homozygous European ancestry at *CFH* (EUR/EUR) in AA AMD cases. c) Enrichment of EUR haplotypes, compared to non-EUR haplotypes in HA AMD cases. d) Enrichment of EUR haplotypes in HA AMD cases, compared to African (AFR) and Native American (NAT) haplotypes.

**Table S19.** Haplotype analysis of the *CFH* locus in admixed African-ancestry (AA) subjects. Case and control frequencies were derived for all haplotypes and stratified by individuals with homozygous African local ancestry (AFR/AFR) or homozygous European local ancestry (EUR/EUR). a) Haplotype model used in Cipriani et al (*38*). b) Haplotype model used in Pappas et al (*39*).

**Table S20.** Local ancestry analysis of haplotypes at the *ARMS2* locus in admixed MVP populations. (**A**) No significant enrichment of European-ancestry haplotypes (EUR) in AA AMD cases. (**B**) No significant enrichment of individuals with homozygous European ancestry at *ARMS2* (EUR/EUR) in AA AMD cases. (**C**) No significant enrichment of EUR haplotypes, compared to non-EUR haplotypes in HA AMD cases. (**D**) Slight enrichment of Native American (NAT) haplotypes in HA AMD cases, compared to African (AFR) and EUR.

**Table S21.** Modeling epistatic interaction between the *CFH* Y402H and *ARMS2* A69S risk alleles in MVP EA subjects. (**A**) Summary statistics of the model including the A69S*Y402H multiplicative interaction term. (**B**) Summary statistics of the model excluding the multiplicative interaction term. (**C**) Likelihood ratio test comparing the two models.

**Table S22.** HLA fine-mapping summary statistics for unrelated EA subjects in MVP. Association with AMD was tested for four-digit alleles at *HLA-A*, *B*, *C*, *DPB1*, *DQA1*, *DQB1*, and *DRB1* imputed by HIBAG software (*41*), considered additively with all other alleles as reference. Association was tested using two models: 1) sex and ten within-ancestry PCs only, and 2) sex, ten within-ancestry PCs, and risk allele dosages for *CFH* (rs1061170), *ARMS2* (rs10490924), *CFB/C2* (rs429608), and *C3* (rs2230199). Firth logistic regression (*80*) was used for both models; p-values and confidence intervals are based on the penalized likelihood ratio test. Alleles with minor allele count less than 50 were excluded from analysis.

**Table S23.** HLA fine-mapping summary statistics for unrelated AA subjects in MVP. Association with AMD was tested for four-digit alleles at *HLA-A*, *B*, *C*, *DPB1*, *DQA1*, *DQB1*, and *DRB1* imputed by HIBAG software (*41*), considered additively with all other alleles as reference. Association was tested using two models: 1) sex and ten within-ancestry PCs only, and 2) sex, ten within-ancestry PCs, and risk allele dosages for *CFH* (rs1061170), *ARMS2* (rs10490924), *CFB/C2* (rs429608), and *C3* (rs2230199). Firth logistic regression (*80*) was used for both models; p-values and confidence intervals are based on the penalized likelihood ratio test. Alleles with minor allele count less than 50 were excluded from analysis.

**Table S24.** HLA fine-mapping summary statistics for unrelated HA subjects in MVP. Association with AMD was tested for four-digit alleles at *HLA-A*, *B*, *C*, *DPB1*, *DQA1*, *DQB1*, and *DRB1* imputed by HIBAG software (*41*), considered additively with all other alleles as reference. Association was tested using two models: 1) sex and ten within-ancestry PCs only, and 2) sex, ten within-ancestry PCs, and risk allele dosages for *CFH* (rs1061170), *ARMS2* (rs10490924), *CFB/C2* (rs429608), and *C3* (rs2230199). Firth logistic regression (*80*) was used for both models; p-values and confidence intervals are based on the penalized likelihood ratio test. Alleles with minor allele count less than 50 were excluded from analysis.

**Table S25.** Transcriptome-wide association scan (TWAS) summary statistics using the brain DLPFC (PsychENCODE) tissue model (N=45,045 cases, 275,806 controls).

**Table S26.** Transcriptome-wide association scan (TWAS) summary statistics using the retina (EyeGEx) tissue model (N=45,045 cases, 275,806 controls).

**Table S27.** TWAS summary statistics from the ACAT meta-analysis of 37 tissue models (N=45,045 cases, 275,806 controls).

**Table S28.** Summary-based Mendelian randomization (SMR) summary statistics for retina gene expression instruments (EyeGEx) (N=45,045 cases, 275,806 controls).

**Table S29.** Gene-set enrichment analysis (GSEA) summary statistics based on the DLPFC TWAS.

**Table S30.** Gene-set enrichment analysis (GSEA) summary statistics based on the TWAS tissue meta-analysis.

**Table S31.** Summary statistics from the gene-based rare variant burden test. All SNPs were ultra-rare (frequency less than 0.25%) and genotyped on the MVP 1.0 array (*72*). Three masks were defined: mask1, containing only frameshift, nonsense, stop-gained, and stop-loss loss of function (LoF) SNPs; mask2 containing LoF SNPs plus high-impact missense variants; and mask3, containing LoF + missense + high-impact splice-site mutations. Burden tests over the masks were conducted in REGENIE (*75*).

**Table S32.** Comparison of the AMD TWAS results to previous studies (*49*, *70*).

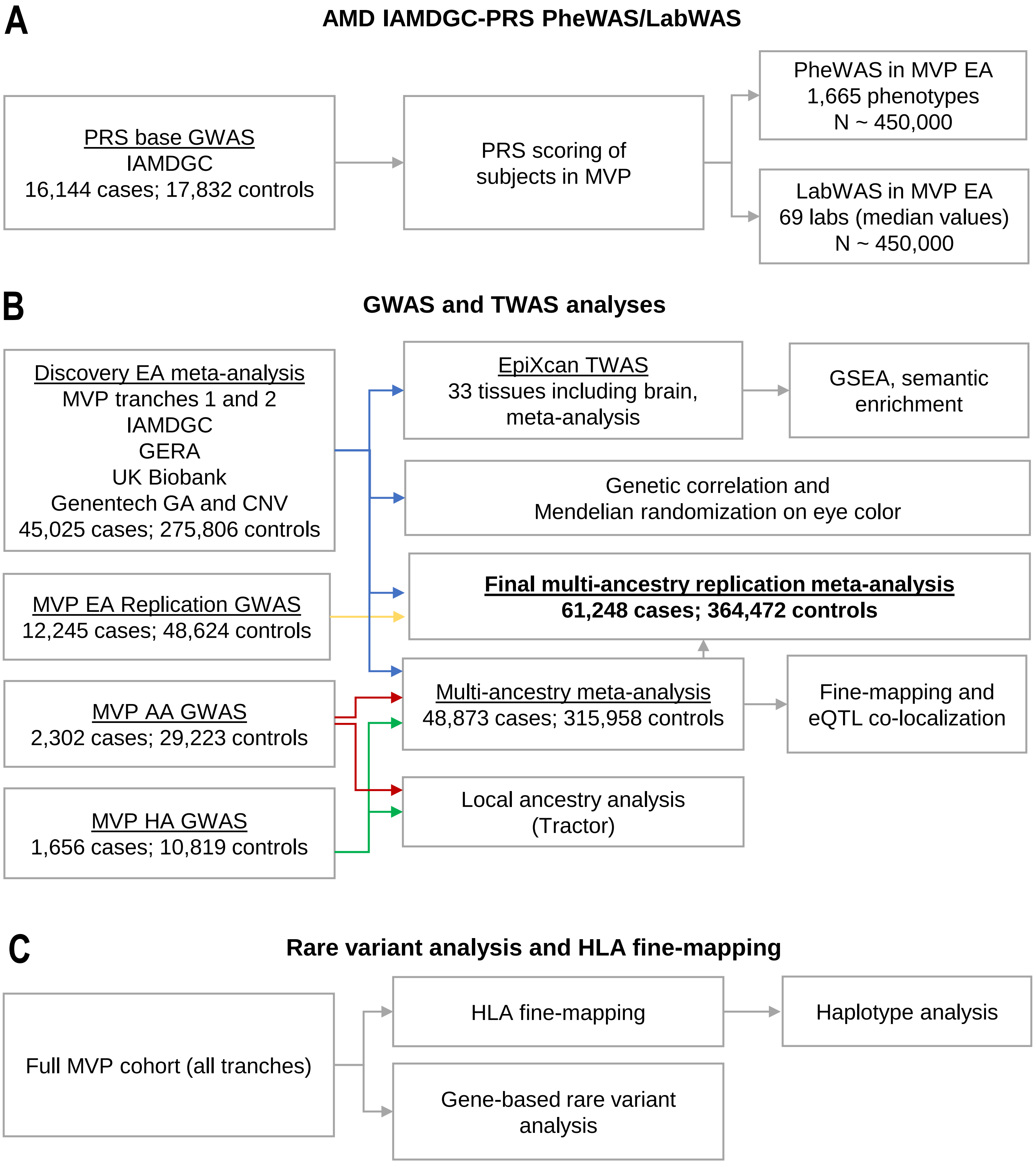


**Fig. S1. Detailed study overview.** (**A**) PRS-PheWAS and PRS-LabWAS analyses using the external European-ancestry IAMDGC GWAS of AMD (*8*) as a base for scoring subjects in MVP. (**B**) Ancestry-specific GWAS analyses, and multi-ancestry meta-analysis, with corresponding secondary analyses including TWAS and Mendelian randomization (EA-only), and fine-mapping (EA and trans-ancestry). An unrelated MVP EA tranche was used as a replication cohort, and meta-analyzed into final EA and multi-ancestry meta-analyses. (**C**) HLA fine-mapping and rare variant analysis.

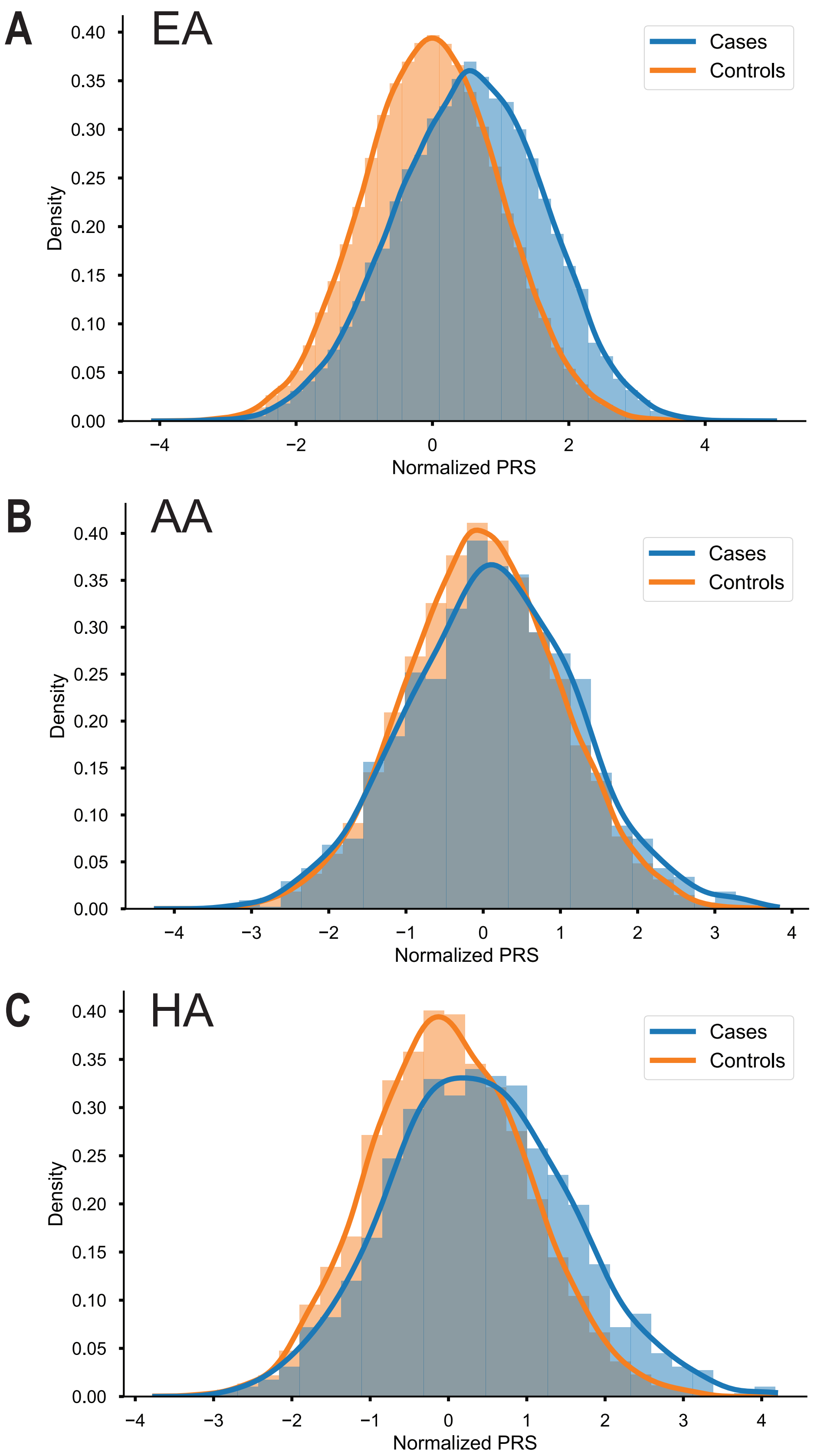


**Fig. S2. PRS distribution of AMD cases and controls.** (**A**) EA samples in MVP; (**B**) AA samples in MVP; and (**C**) HA samples in MVP. The PRS was constructed using weights derived from the IAMDGC GWAS (*8*).

**Fig. S3. Novel genome-wide significant loci from the European-ancestry meta-analysis.** Regional association plots of novel loci from the European-ancestry replication meta-analysis (N=57,290 cases, 324,430 controls). The 1000 Genomes EUR panel was used as the reference for plotting LD $r^2$.

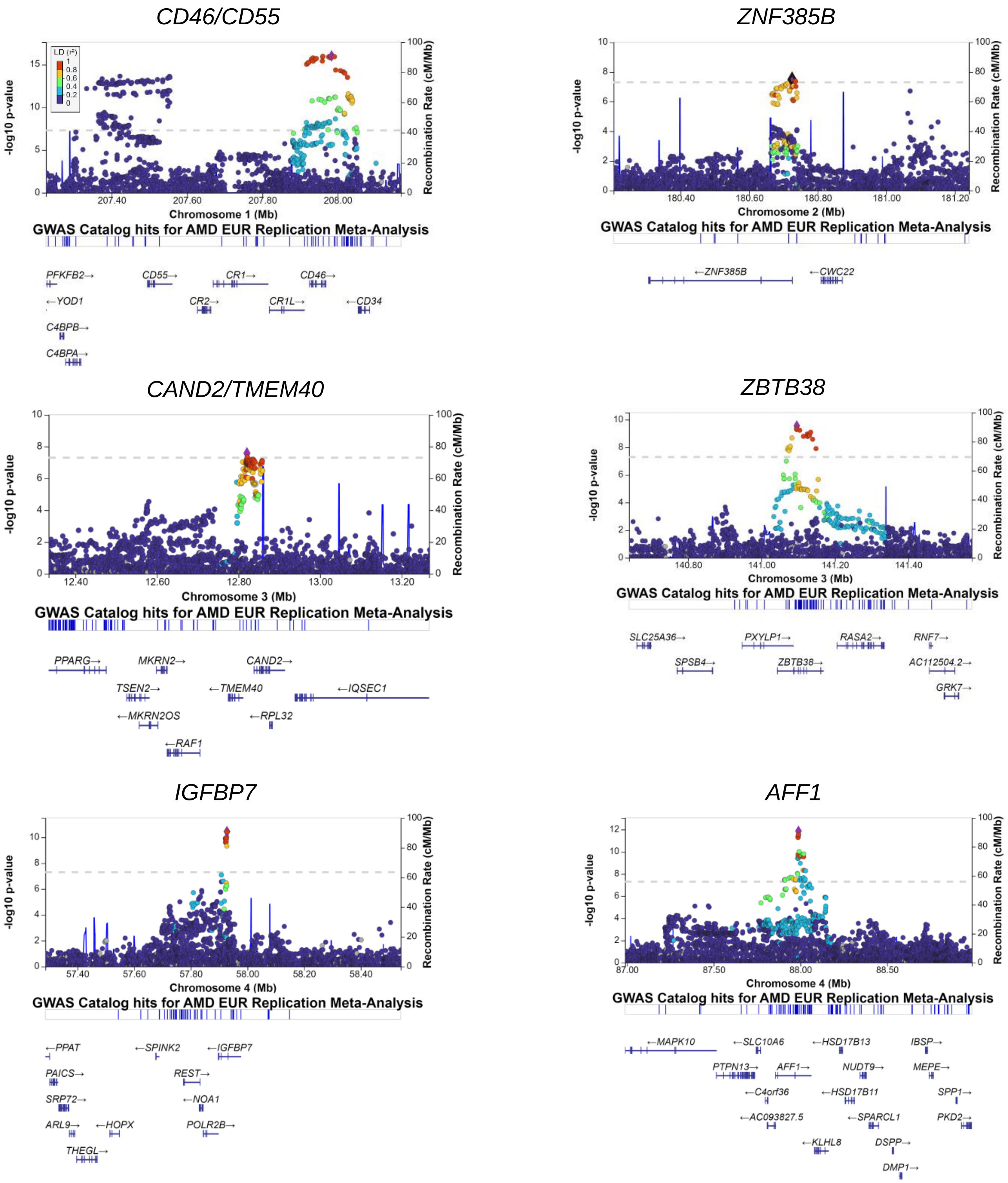

*ADAM19*

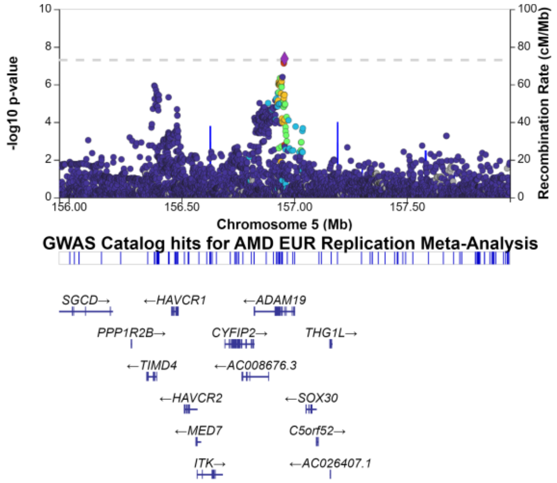


*HSDL2*

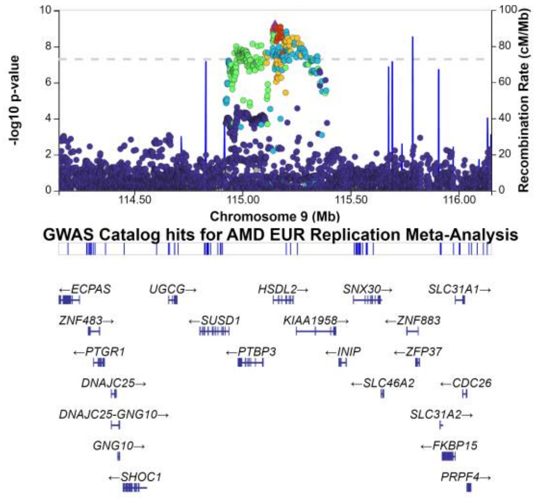


*ME3*

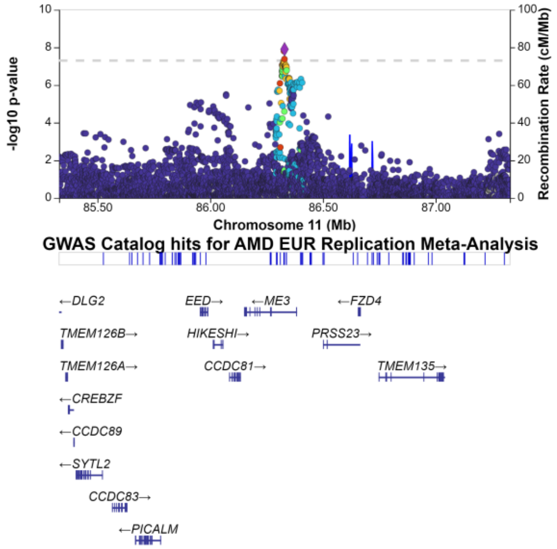


*TYR*

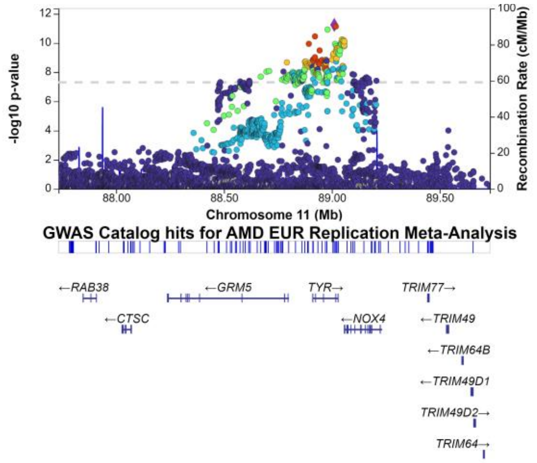


*LINC02343*

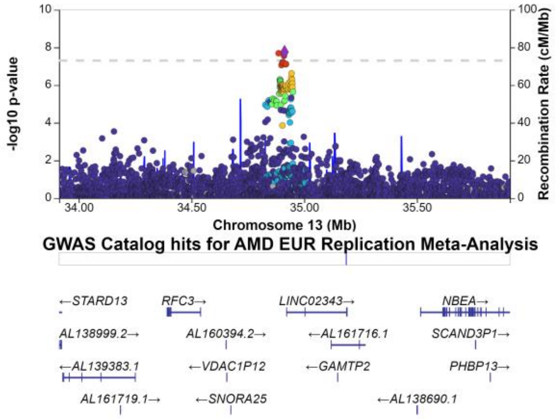


*SERPINA1*

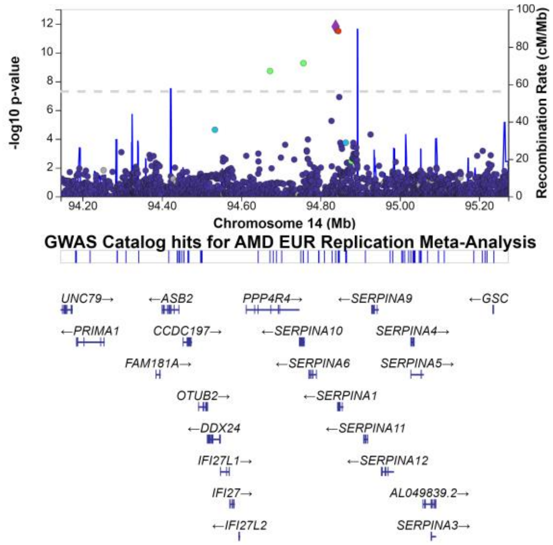

*HERC2/OCA2*

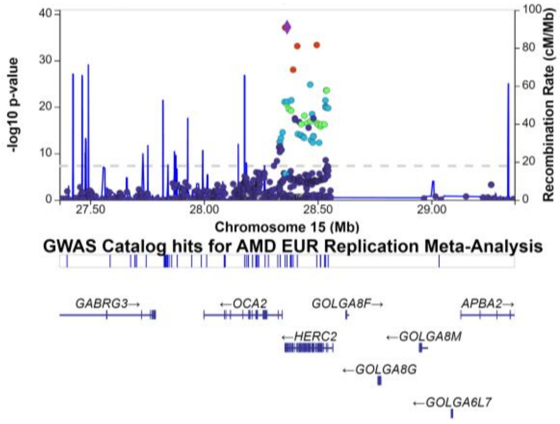


*TRMP1*

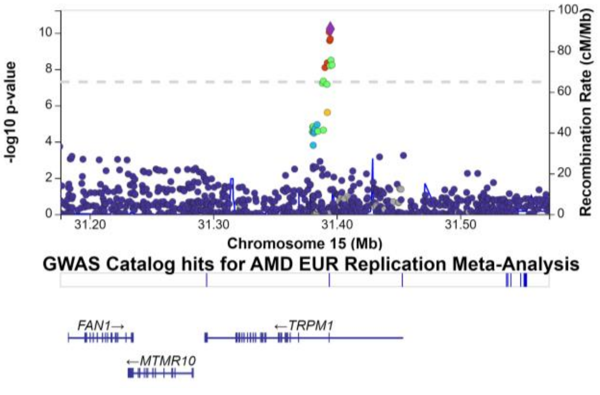


*MYO1E*

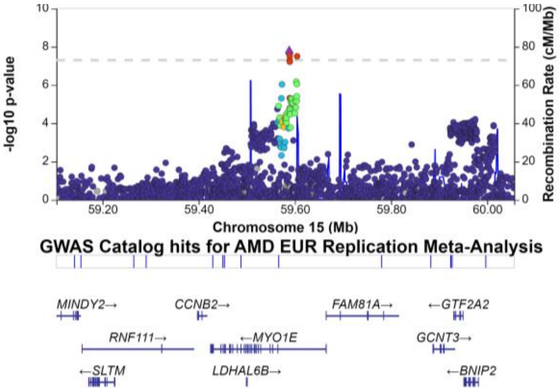


*SMAD3*

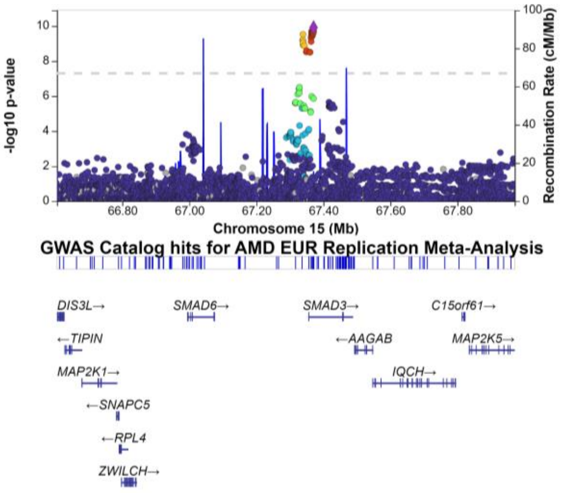


*CSK*

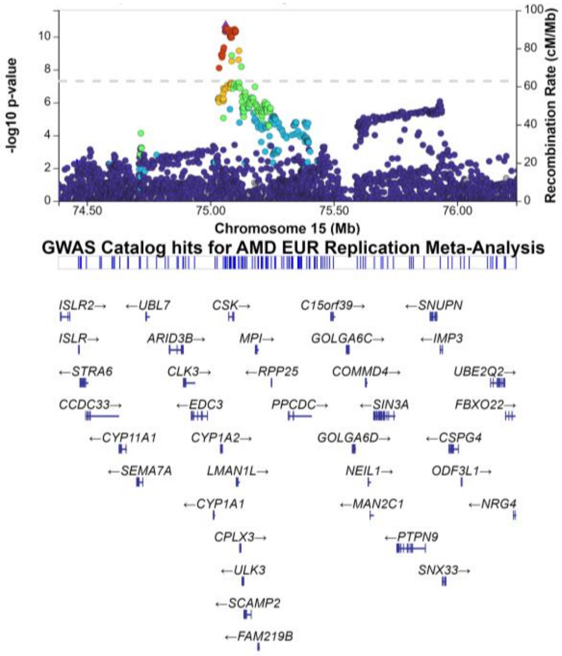


*RLBP1*

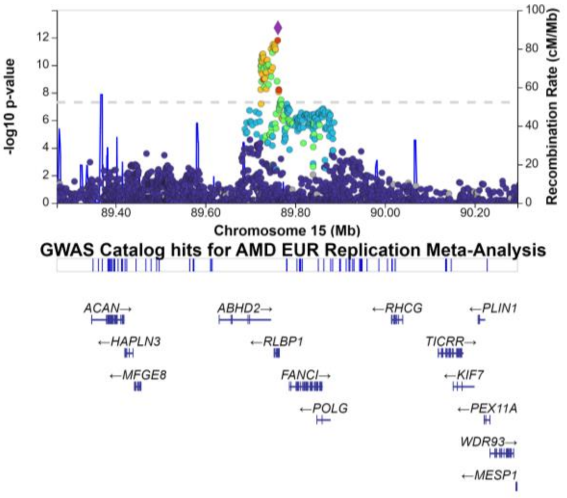

*CHD9*

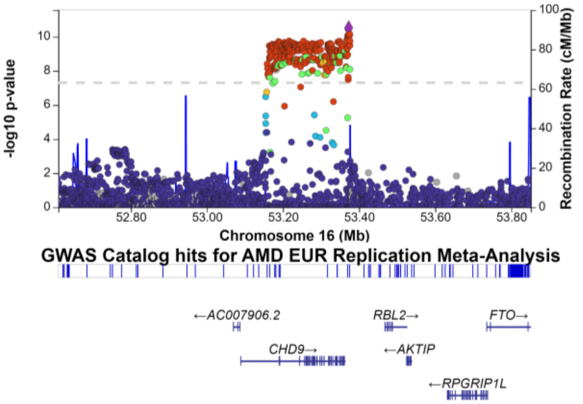


*CLUL1*

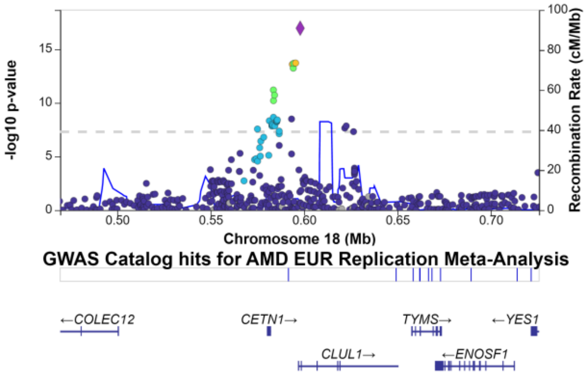


*ACAA2/LIPG*

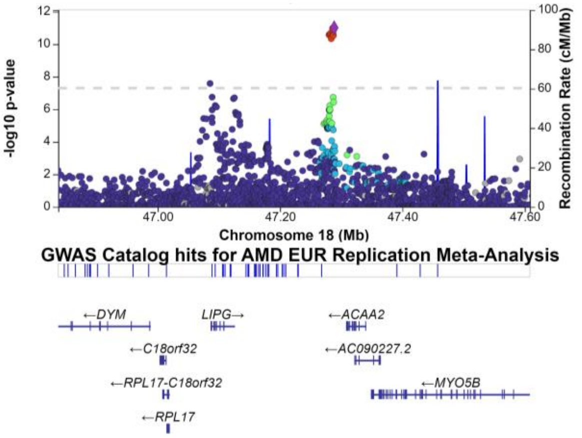


*TGFB1*

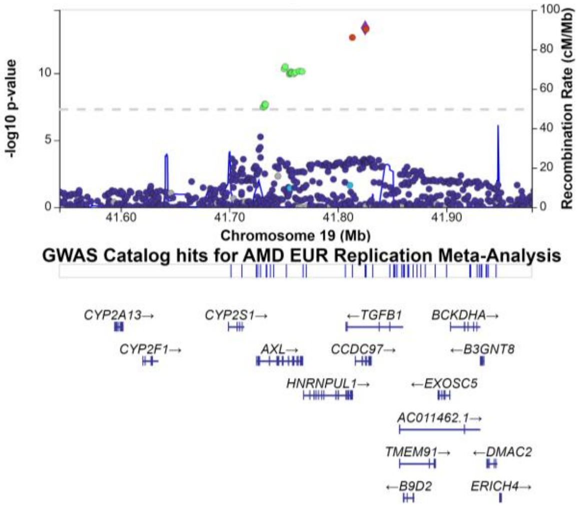


*RASIP1*

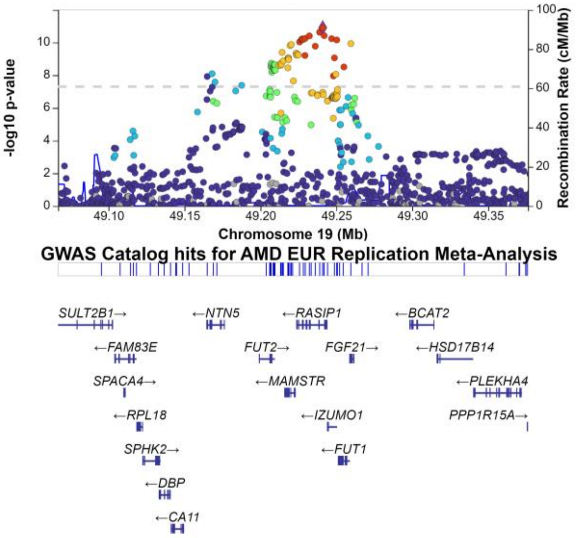


*MARK4/EXOC3L2*

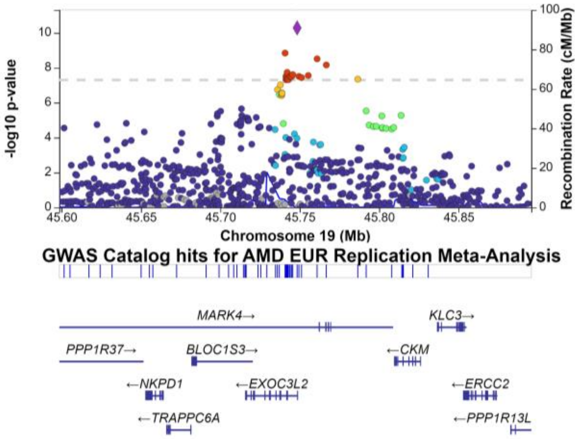

*RRAS*

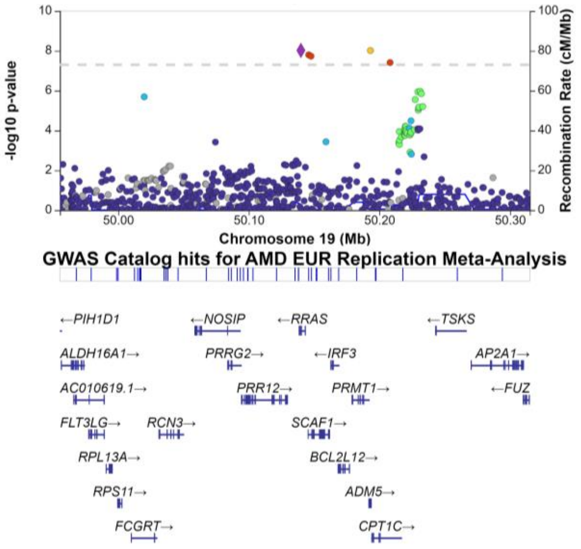


*LBP*

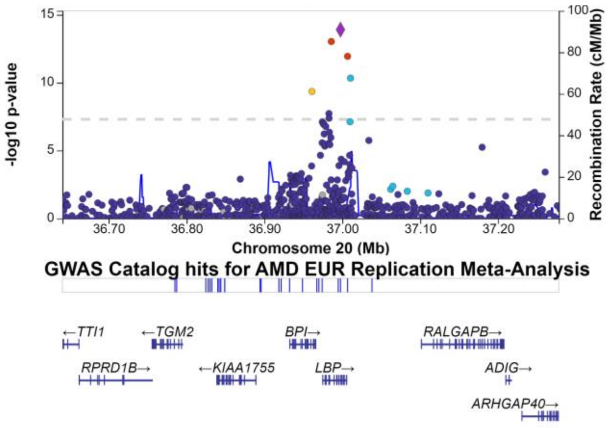


*PDGFB*

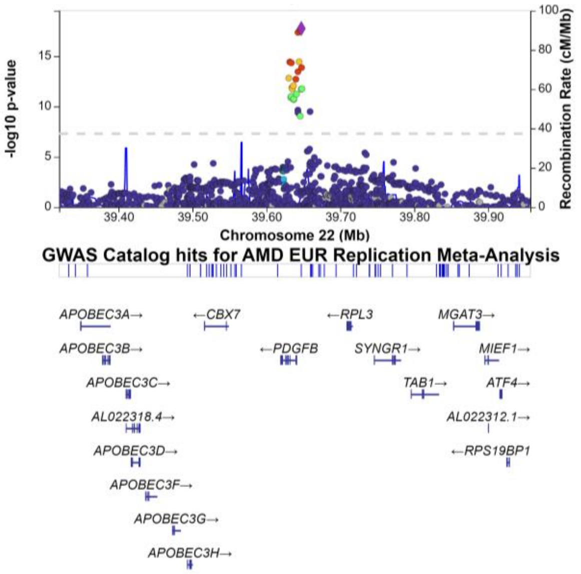

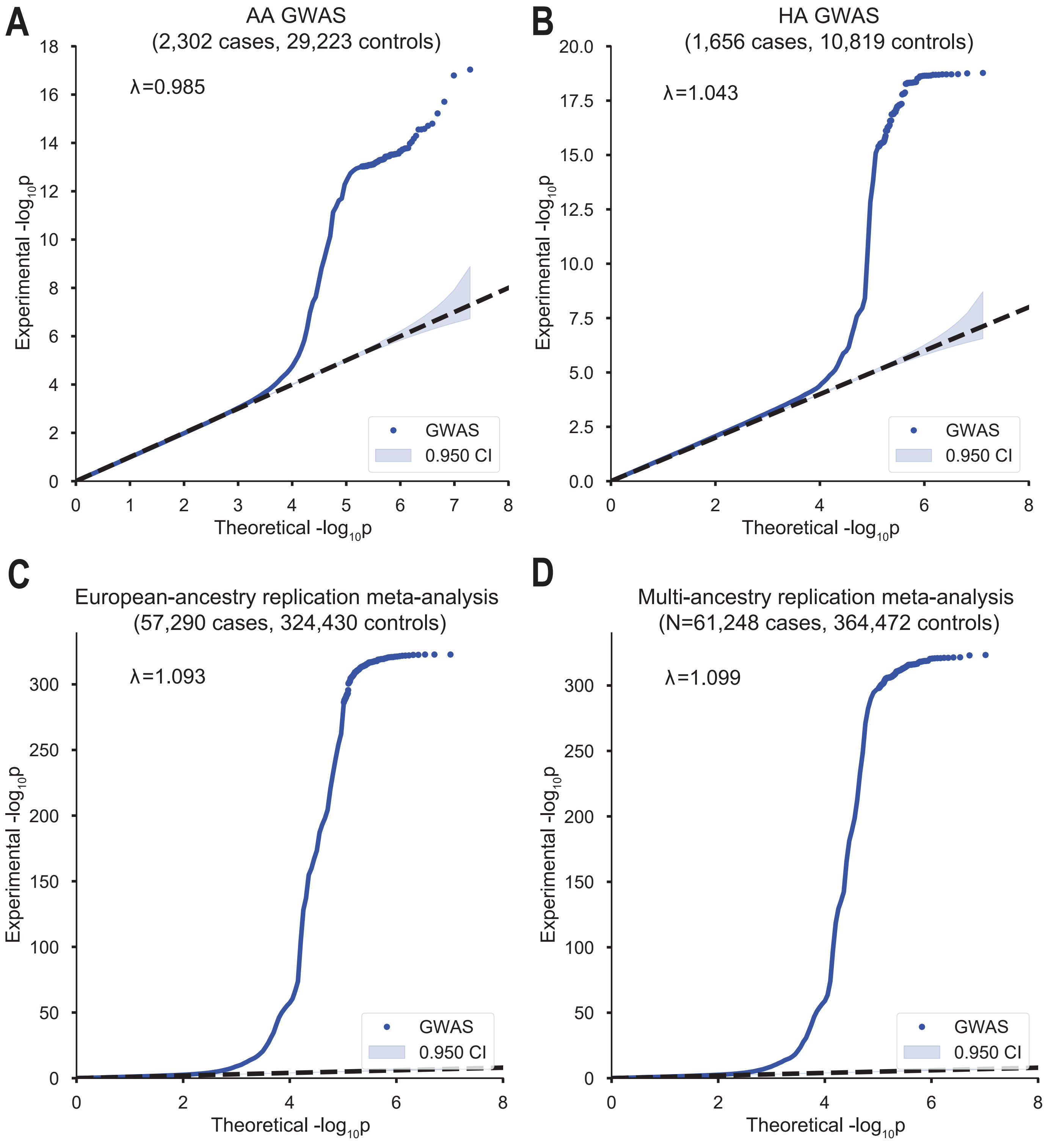


**Fig. S4. QQ plots of the multi-ancestry GWAS and meta-analysis.** (**A**) MVP AA GWAS, (**B**) MVP HA GWAS, (**C**) EA meta-analysis, and (**D**) multi-ancestry meta-analysis. SNPs with minor allele frequency greater than 0.5% are plotted and included in the genomic control inflation calculation.

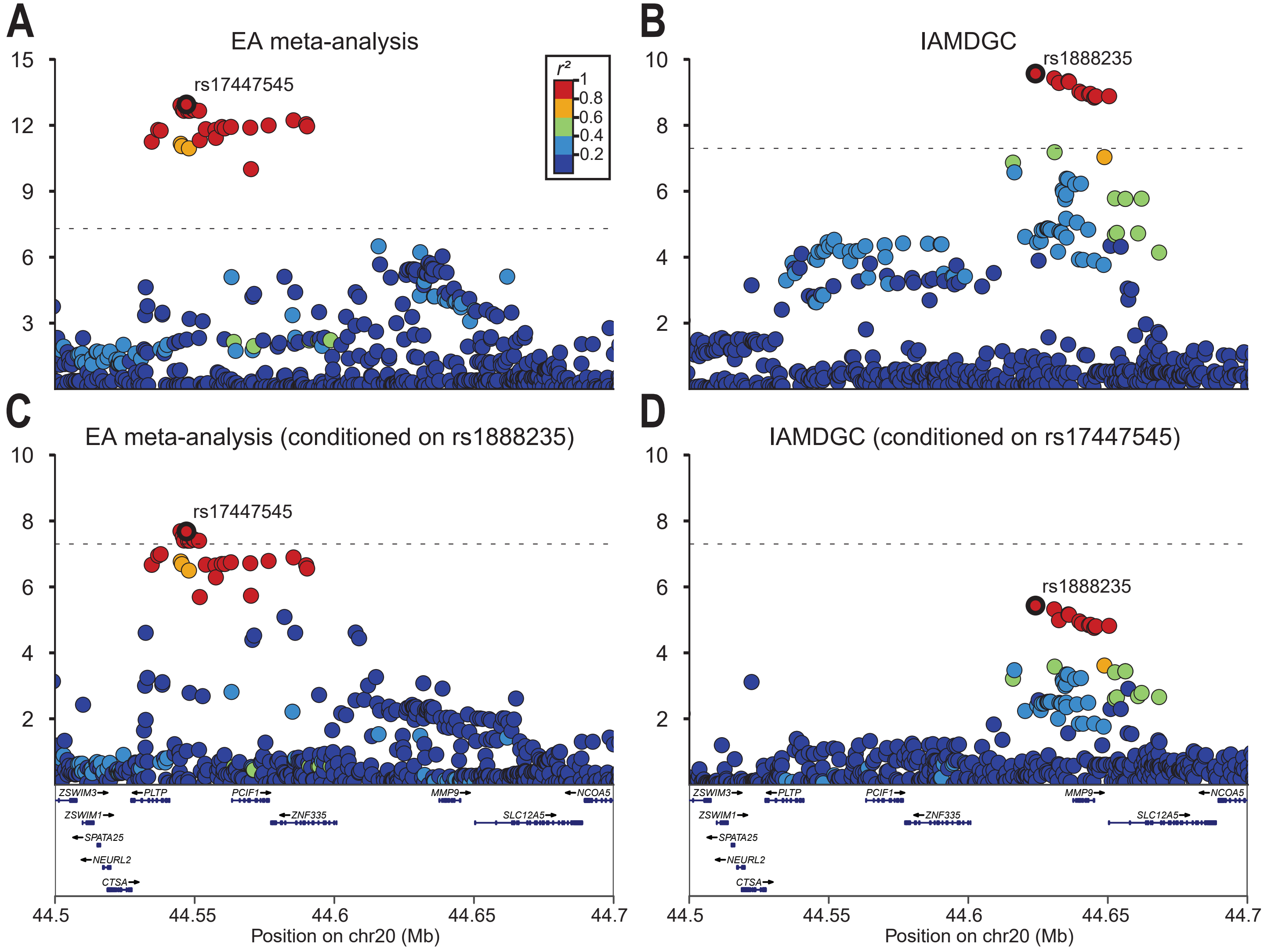


**Fig. S5. Conditional analysis of *PLTP* and *MMP9* signals in the 20q13 locus.** Regional association plots of: (**A**) the EA replication meta-analysis, demonstrating a primary signal near *PLTP*; (**B**) the IAMDGC study (*8*), demonstrating a primary signal near *MMP9*; (**C**) the EA meta-analysis conditioned on the index SNP from the IAMDGC study (rs1888235); (**D**) the IAMDGC study conditioned on the index SNP from the EA meta-analysis (rs17447545). LD $r^2$ values were calculated in the MVP EA cohort. The dashed line denotes genome-wide significance ($5\times10^{-8}$).

### African-ancestry (AA) GWAS

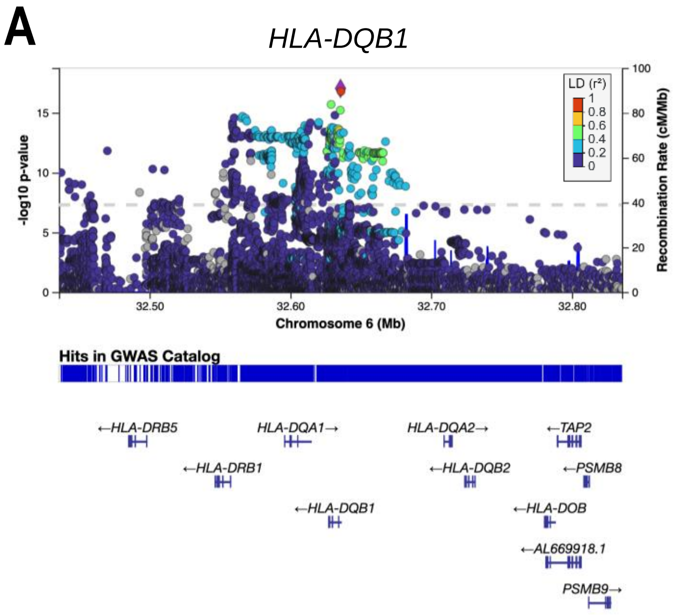


### Hispanic-ancestry (HA) GWAS

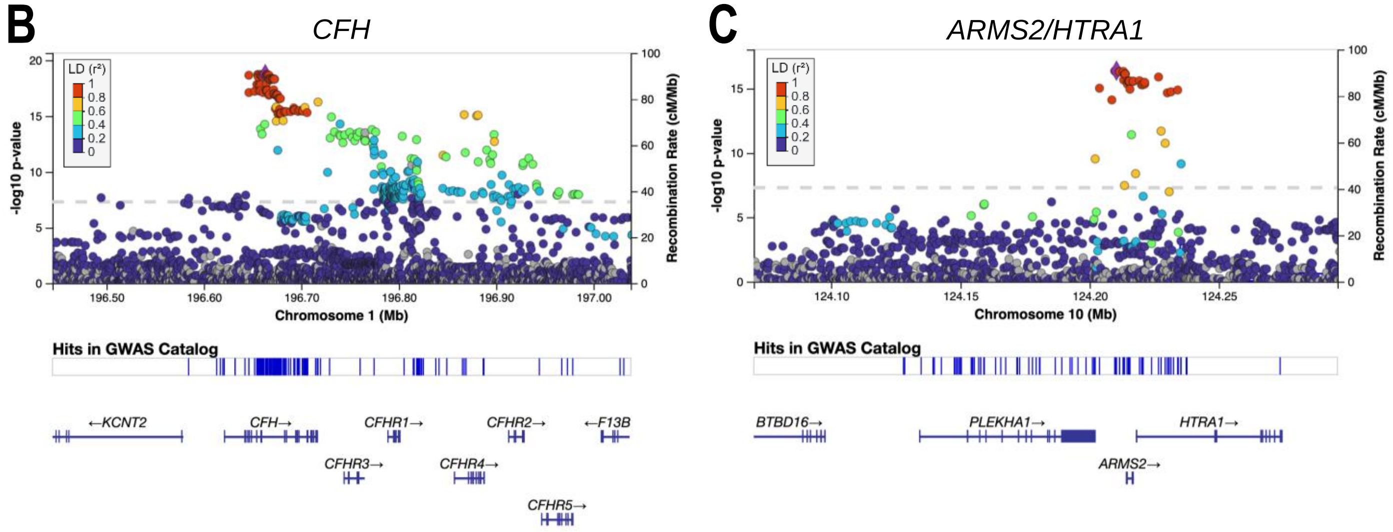


**Fig. S6. Genome-wide significant loci from the AA and HA GWAS in MVP.** (**A**) Regional association plot of the *HLA-DQB1* locus from the AA GWAS (2,302 cases, 29,223 controls). The 1000 Genomes AFR panel was used as the reference for plotting LD $r^2$. (**B-C**) Regional association plots of the *CFH* and *ARMS2-HTRA1* loci from the HA GWAS (1,656 cases, 10,819 controls). The 1000 Genomes AMR panel was used as the reference for plotting LD $r^2$.

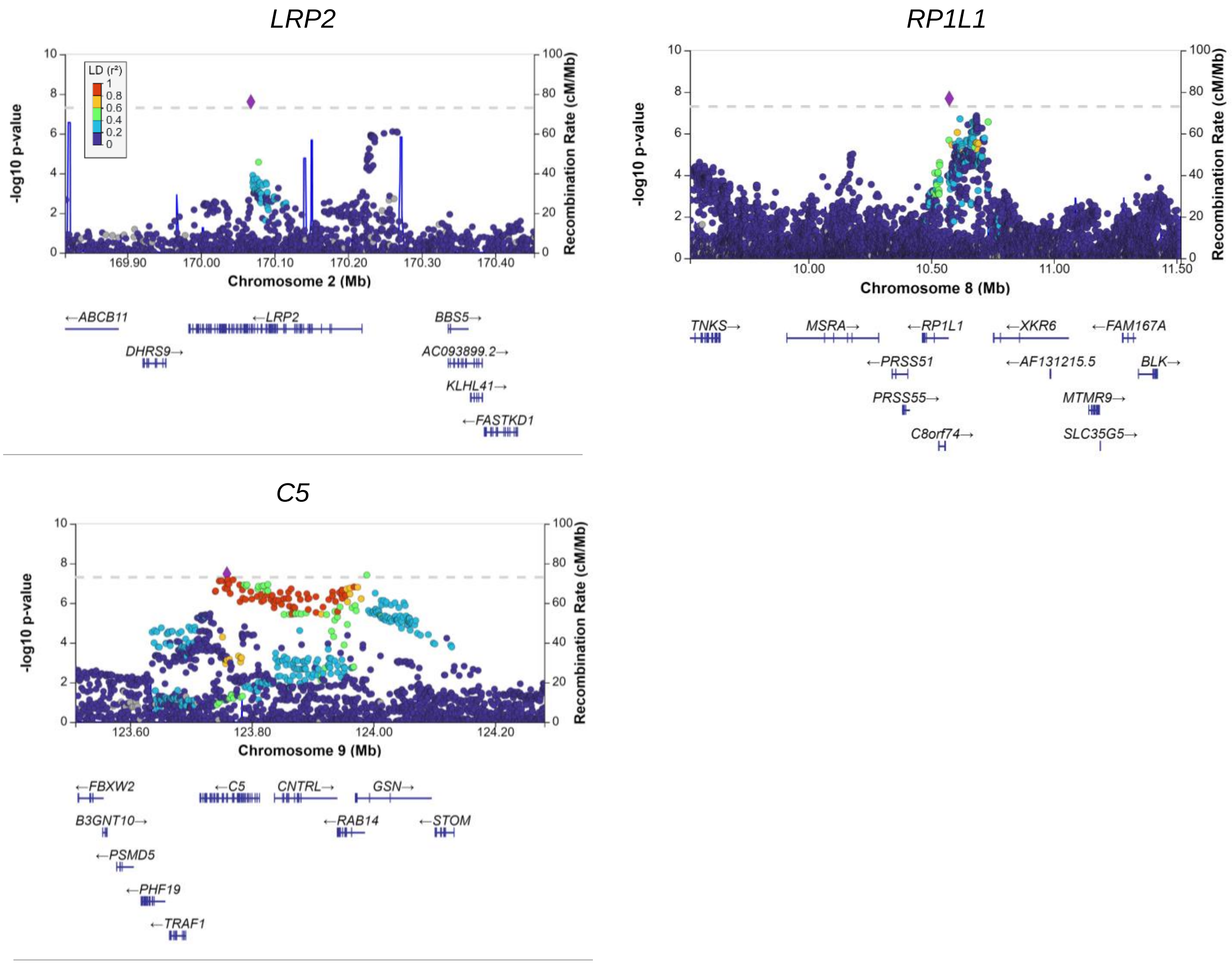


**Fig. S7. Novel genome-wide significant loci from the multi-ancestry meta-analysis.** Regional association plots of novel loci from the multi-ancestry replication meta-analysis (61,258 cases, 364,472 controls) beyond that reported for the EA meta-analysis. The 1000 Genomes EUR panel was used as the reference for plotting LD $r^2$.

**Fig. S8. Co-localization of GWAS and eQTL signals identifies candidate causal variants.** Shown are significant co-localized signals (co-localization posterior probability > 0.01) from the multi-ancestry discovery meta-analysis using the mmQTL (*31*) method.

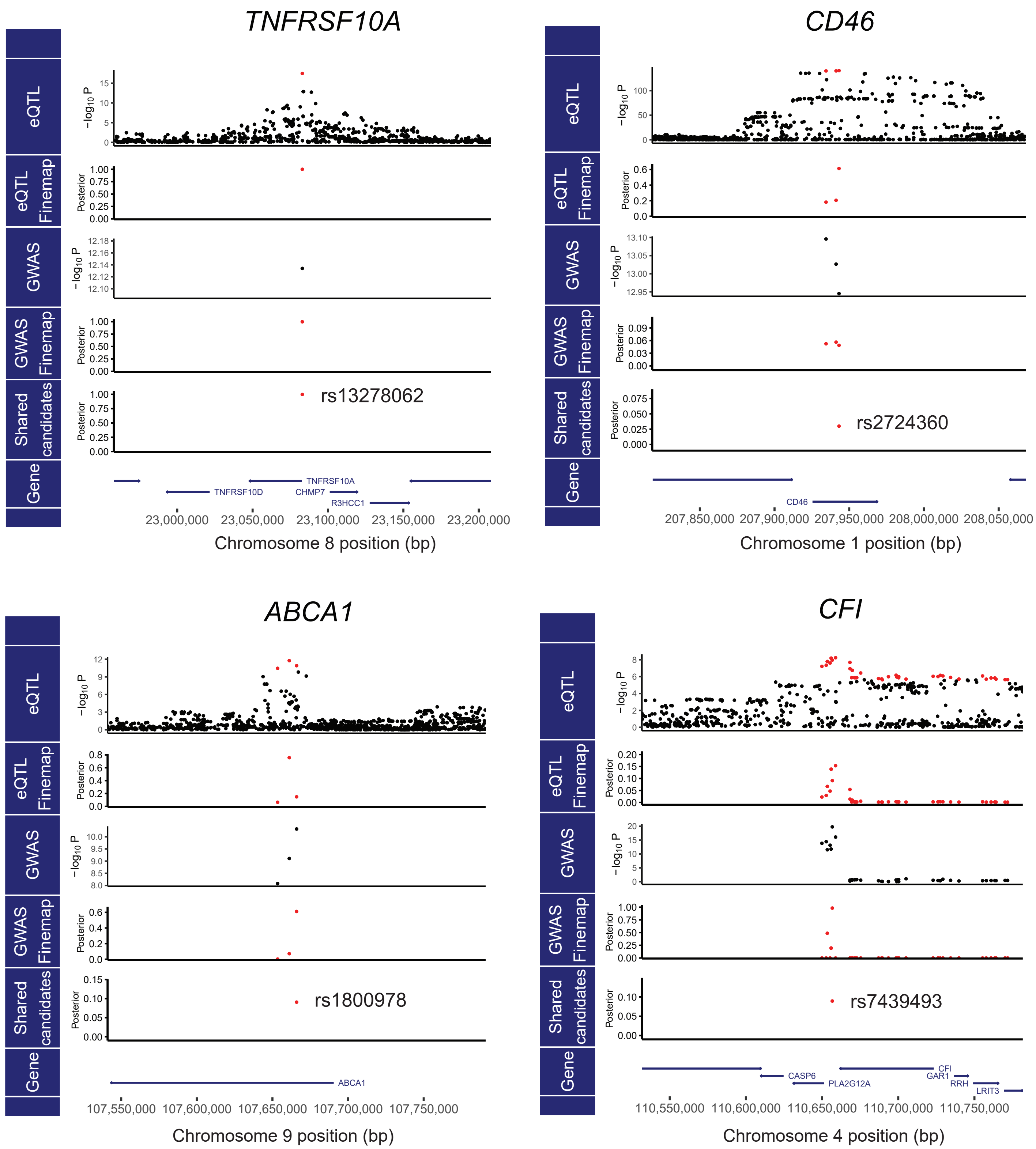

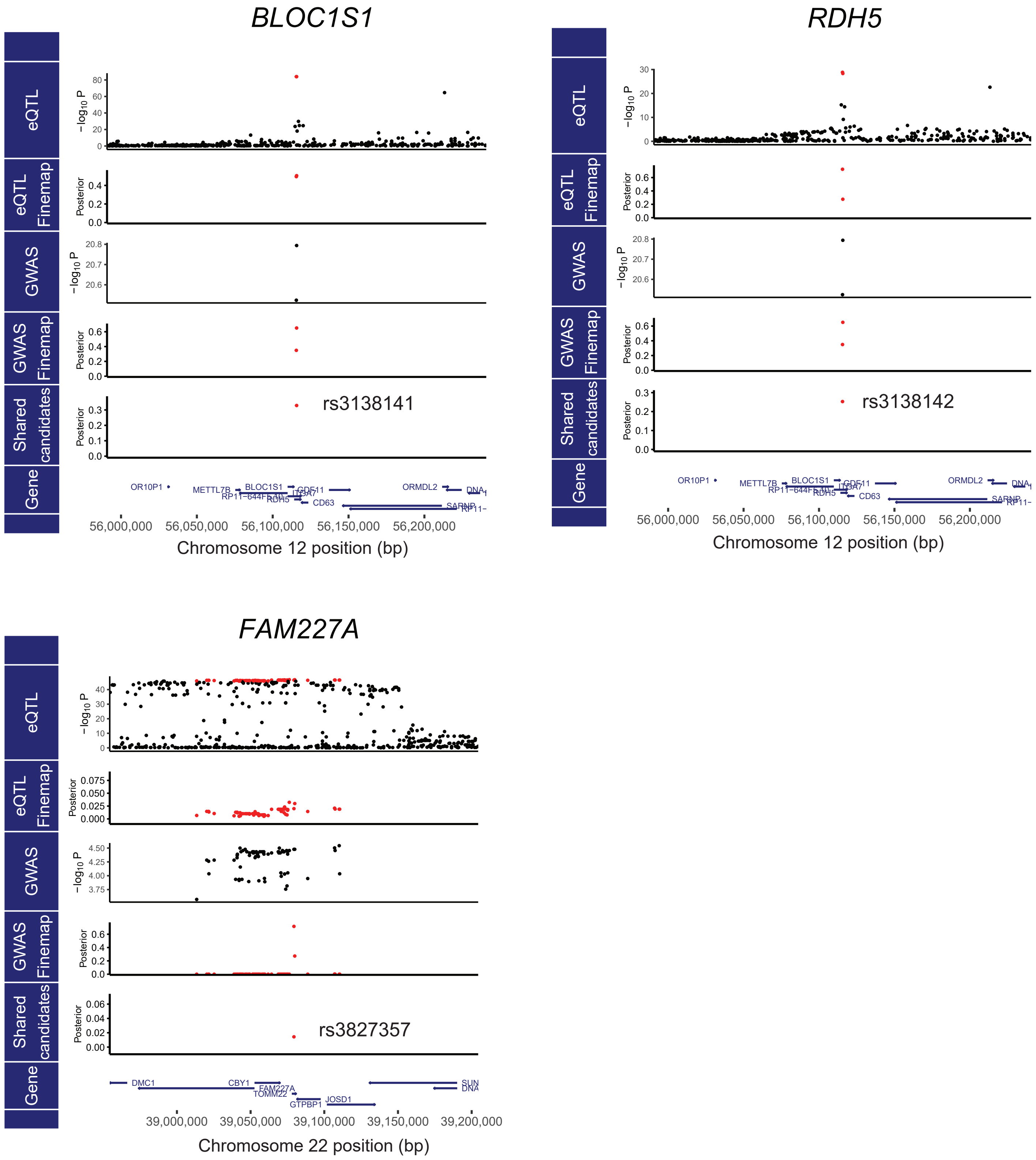

BLOC1S1
RDH5
FAM227A
eQTL
eQTL Finemap
GWAS
GWAS Finemap
Shared candidates
Gene
−log10 P
Posterior
rs3138141
rs3138142
rs3827357
OR10P1
METTL7B
BLOC1S1
RP11−644F5.10
RDH5
CDK2
ITGA7
CD63
GDF11
ORMDL2
SARNP
DNA
DMC1
CBY1
FAM227A
TOMM22
GTPBP1
JOSD1
SUN
56,000,000
56,050,000
56,100,000
56,150,000
56,200,000
Chromosome 12 position (bp)
39,000,000
39,050,000
39,100,000
39,150,000
39,200,000
Chromosome 22 position (bp)

A

B

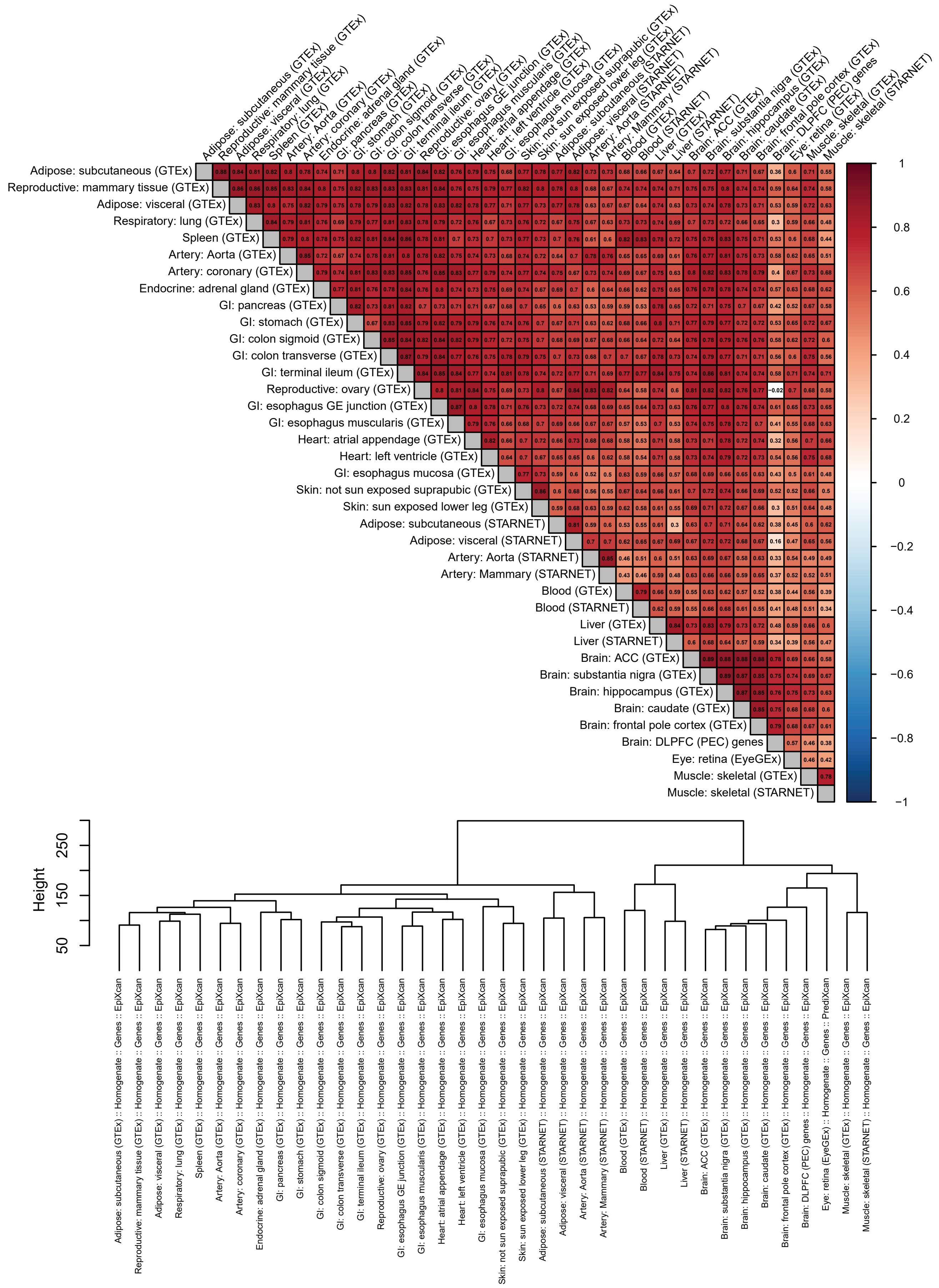


**Fig. S9. Characterization of TWAS results across tissues.** (**A**) Pearson correlation of AMD association z-scores across the 38 tissues analyzed from GTEx, EyeGEx, STARNET, and PEC. (**B**) Hierarchical clustering of tissues based on AMD associations using Ward's linkage method.

**Fig. S10. Regional Miami plots of genes with novel TWAS associations.** The TWAS was based on the DLPFC brain model jointly testing genes and transcript isoforms. Results are based on the EA discovery meta-analysis (N=45,045 cases, 275,806 controls). The GWAS p-values are shown above the y=0 line, and the TWAS p-values are shown below it. SNPs are colored according to linkage disequilibrium r-squared with the lead SNP (EA panel). Lines connecting SNP predictors of genetically regulated gene expression are colored according to the significance level of the SNP: orange, p≤5×$10^{-8}$; cyan, p≤1×$10^{-5}$; gray, p>1×$10^{-5}$.

**A**

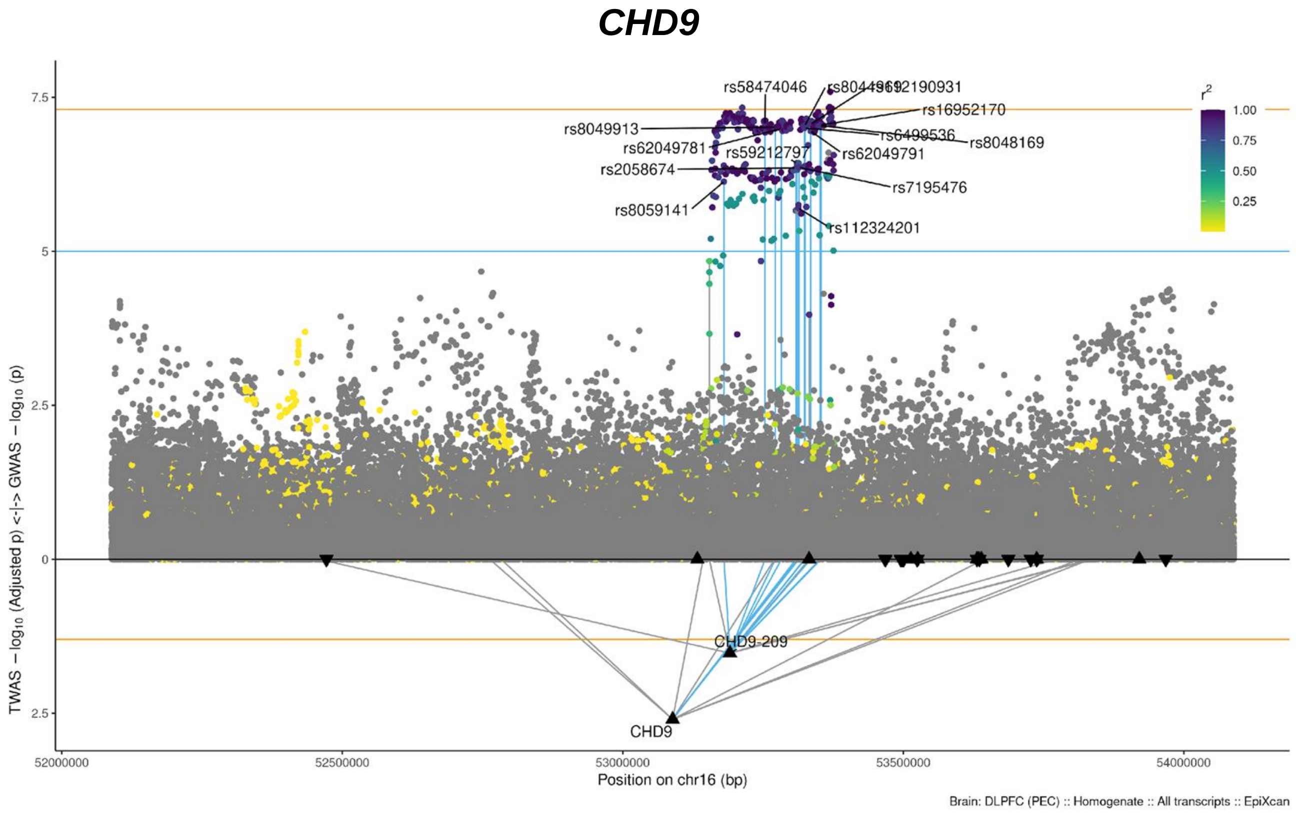

B

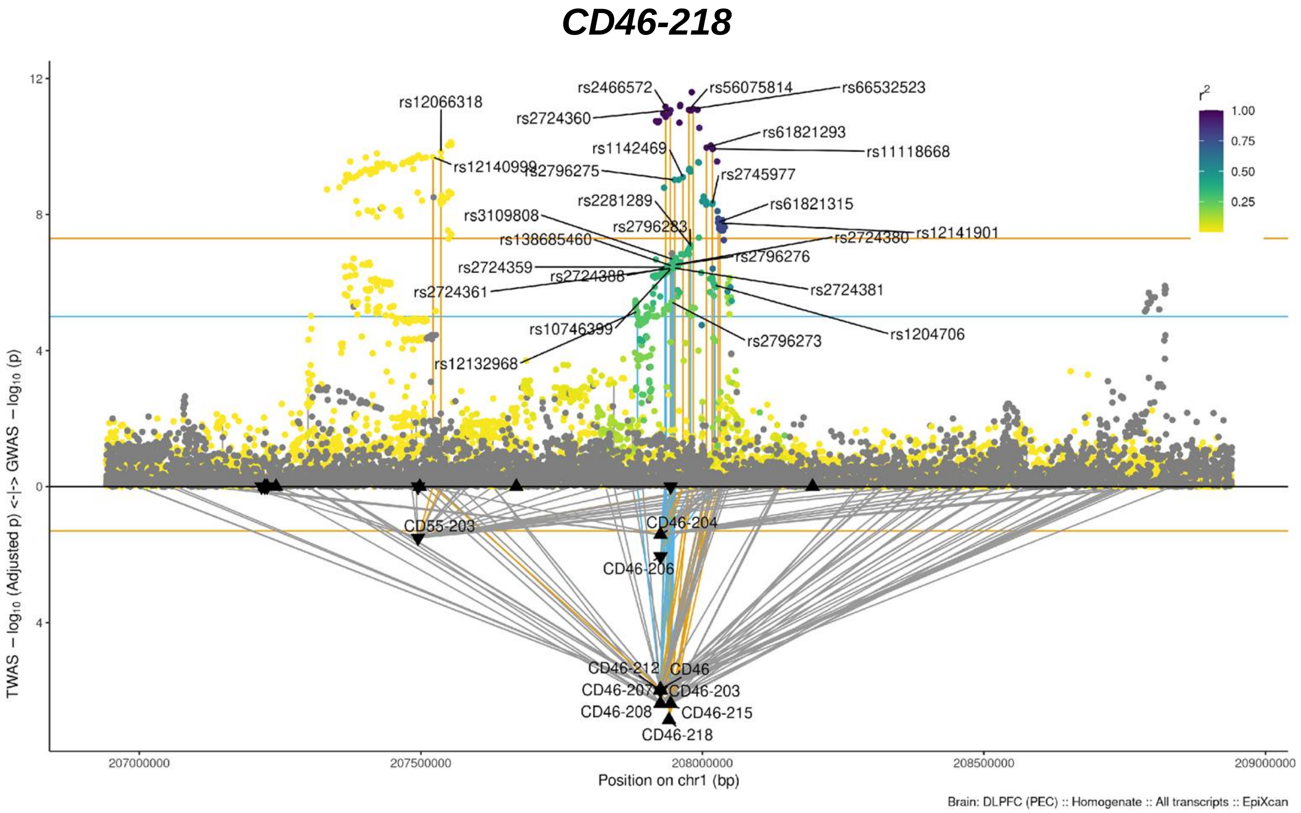

CD46-218
rs12066318
rs12140999
rs2466572
rs2724360
rs56075814
rs66532523
rs61821293
rs11118668
rs1142469
rs2796275
rs2745977
rs2281289
rs61821315
rs3109808
rs2796283
rs138685460
rs2724380
rs12141901
rs2724359
rs2796276
rs2724388
rs2724361
rs2724381
rs10746399
rs2796273
rs1204706
rs12132968
CD55-203
CD46-204
CD46-206
CD46-212
CD46
CD46-207
CD46-203
CD46-208
CD46-215
CD46-218
r^2
1.00
0.75
0.50
0.25
12
8
4
0
4
TWAS − log10 (Adjusted p) <-|-> GWAS − log10 (p)
207000000
207500000
208000000
208500000
209000000
Position on chr1 (bp)
Brain: DLPFC (PEC) :: Homogenate :: All transcripts :: EpiXcan


C

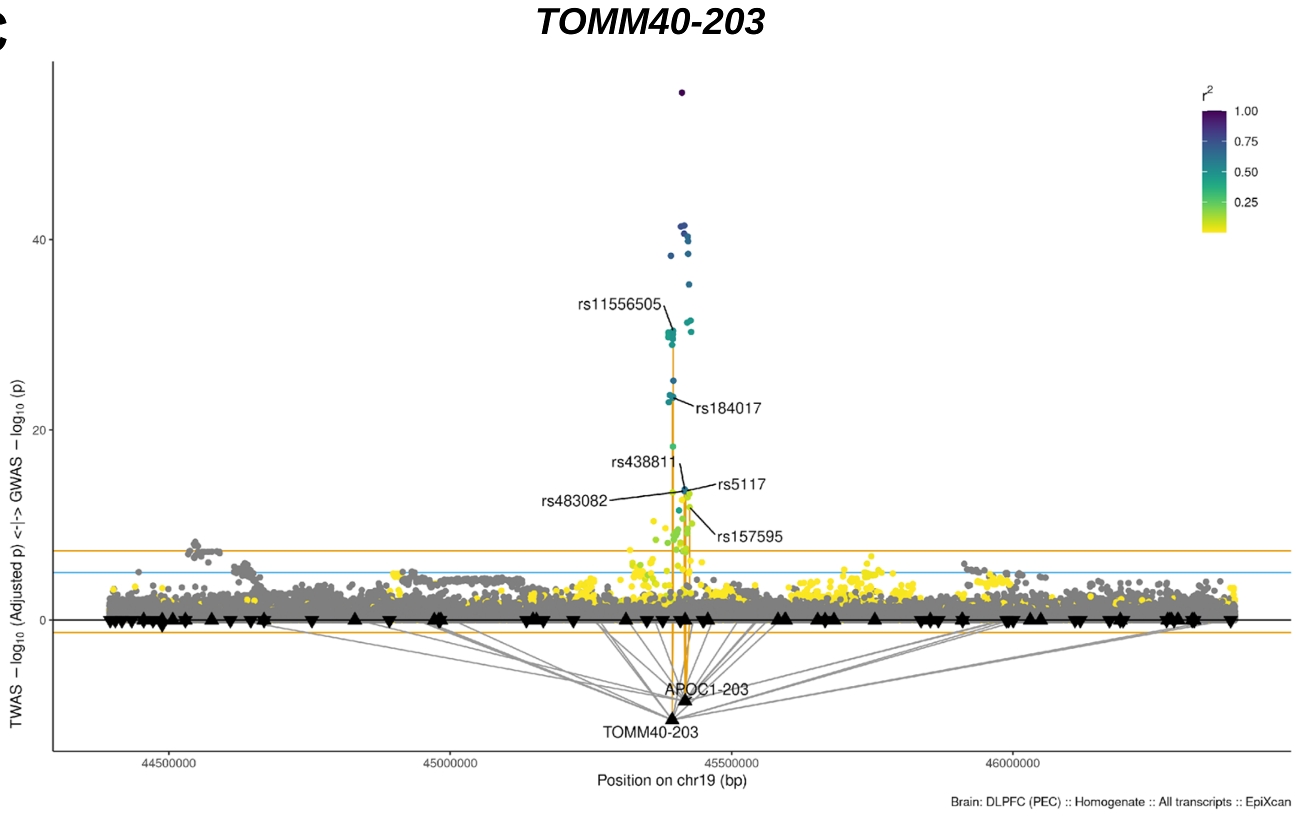

TOMM40-203
rs11556505
rs184017
rs438811
rs5117
rs483082
rs157595
APOC1-203
TOMM40-203
r^2
1.00
0.75
0.50
0.25
40
20
0
TWAS − log10 (Adjusted p) <-|-> GWAS − log10 (p)
44500000
45000000
45500000
46000000
Position on chr19 (bp)
Brain: DLPFC (PEC) :: Homogenate :: All transcripts :: EpiXcan

D

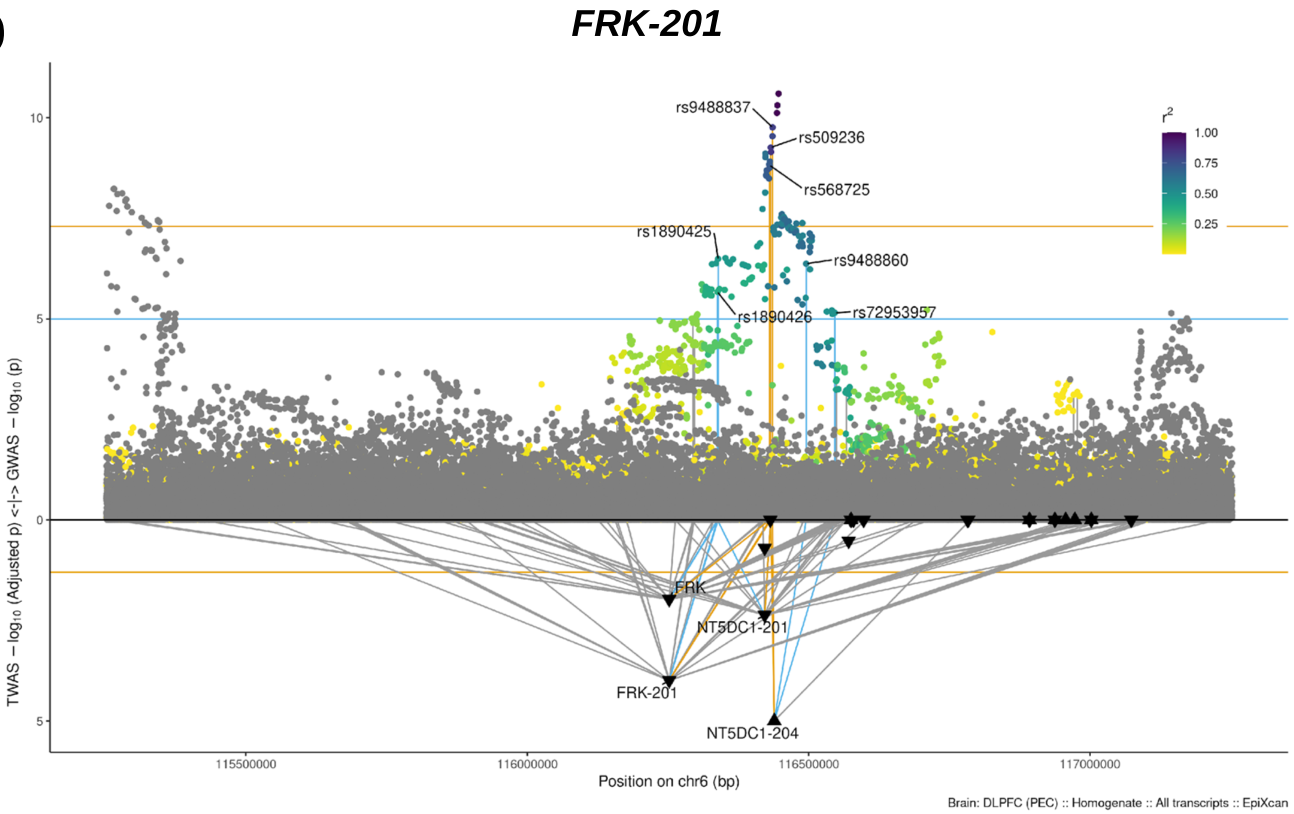
FRK-201
rs9488837
rs509236
rs568725
rs1890425
rs9488860
rs1890426
rs72953957
r2
1.00
0.75
0.50
0.25
10
5
0
5
FRK
NT5DC1-201
FRK-201
NT5DC1-204
TWAS − log10 (Adjusted p) <-|-> GWAS − log10 (p)
115500000
116000000
116500000
117000000
Position on chr6 (bp)
Brain: DLPFC (PEC) :: Homogenate :: All transcripts :: EpiXcan

E

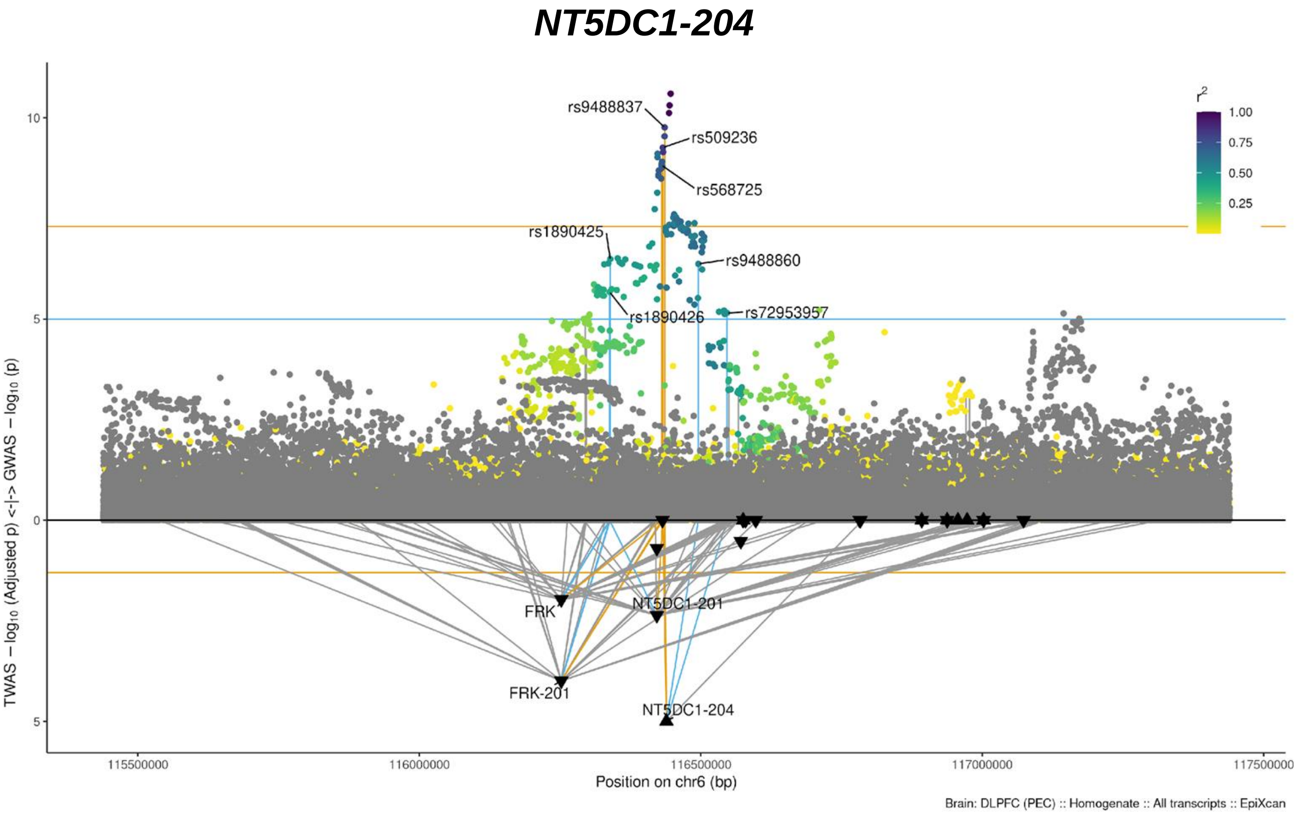
NT5DC1-204
rs9488837
rs509236
rs568725
rs1890425
rs9488860
rs1890426
rs72953957
r2
1.00
0.75
0.50
0.25
10
5
0
5
FRK
NT5DC1-201
FRK-201
NT5DC1-204
TWAS − log10 (Adjusted p) <-|-> GWAS − log10 (p)
115500000
116000000
116500000
117000000
117500000
Position on chr6 (bp)
Brain: DLPFC (PEC) :: Homogenate :: All transcripts :: EpiXcan

F

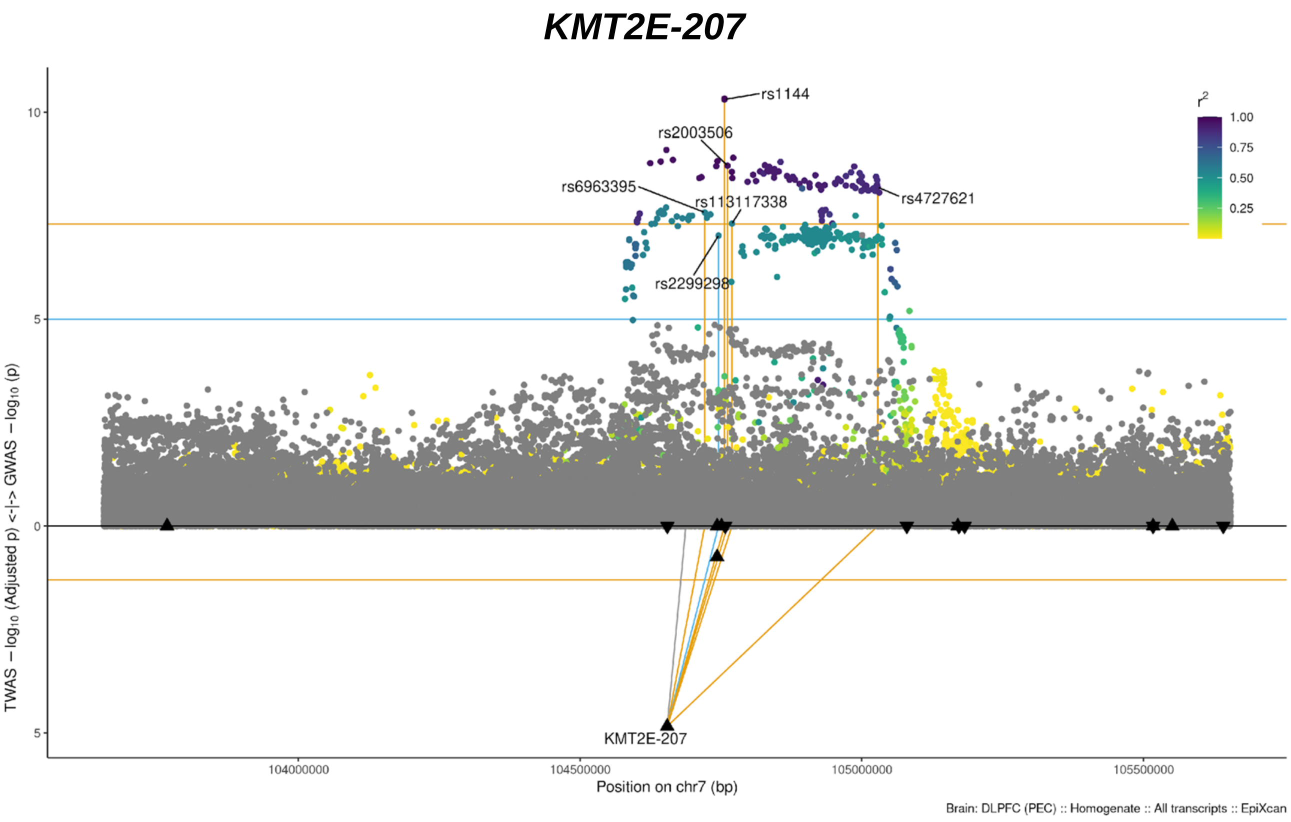
KMT2E-207
rs1144
rs2003506
rs6963395
rs113117338
rs4727621
rs2299298
KMT2E-207
r2
1.00
0.75
0.50
0.25
10
5
0
5
TWAS − log10 (Adjusted p) <-|-> GWAS − log10 (p)
104000000
104500000
105000000
105500000
Position on chr7 (bp)
Brain: DLPFC (PEC) :: Homogenate :: All transcripts :: EpiXcan

G

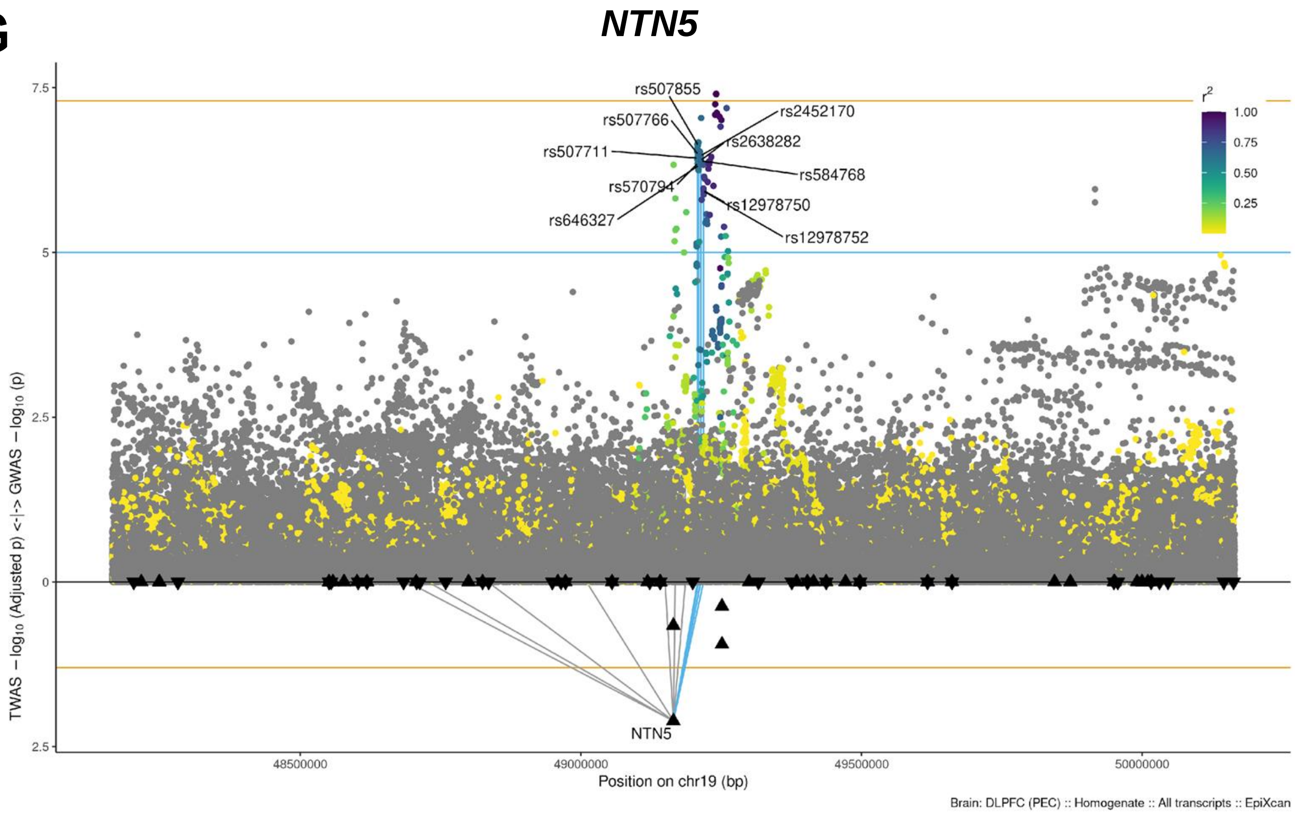
NTN5
rs507855
rs2452170
rs507766
rs2638282
rs507711
rs584768
rs570794
rs12978750
rs646327
rs12978752
NTN5
r2
1.00
0.75
0.50
0.25
7.5
5
2.5
0
2.5
TWAS − log10 (Adjusted p) <-|-> GWAS − log10 (p)
48500000
49000000
49500000
50000000
Position on chr19 (bp)
Brain: DLPFC (PEC) :: Homogenate :: All transcripts :: EpiXcan

**H**

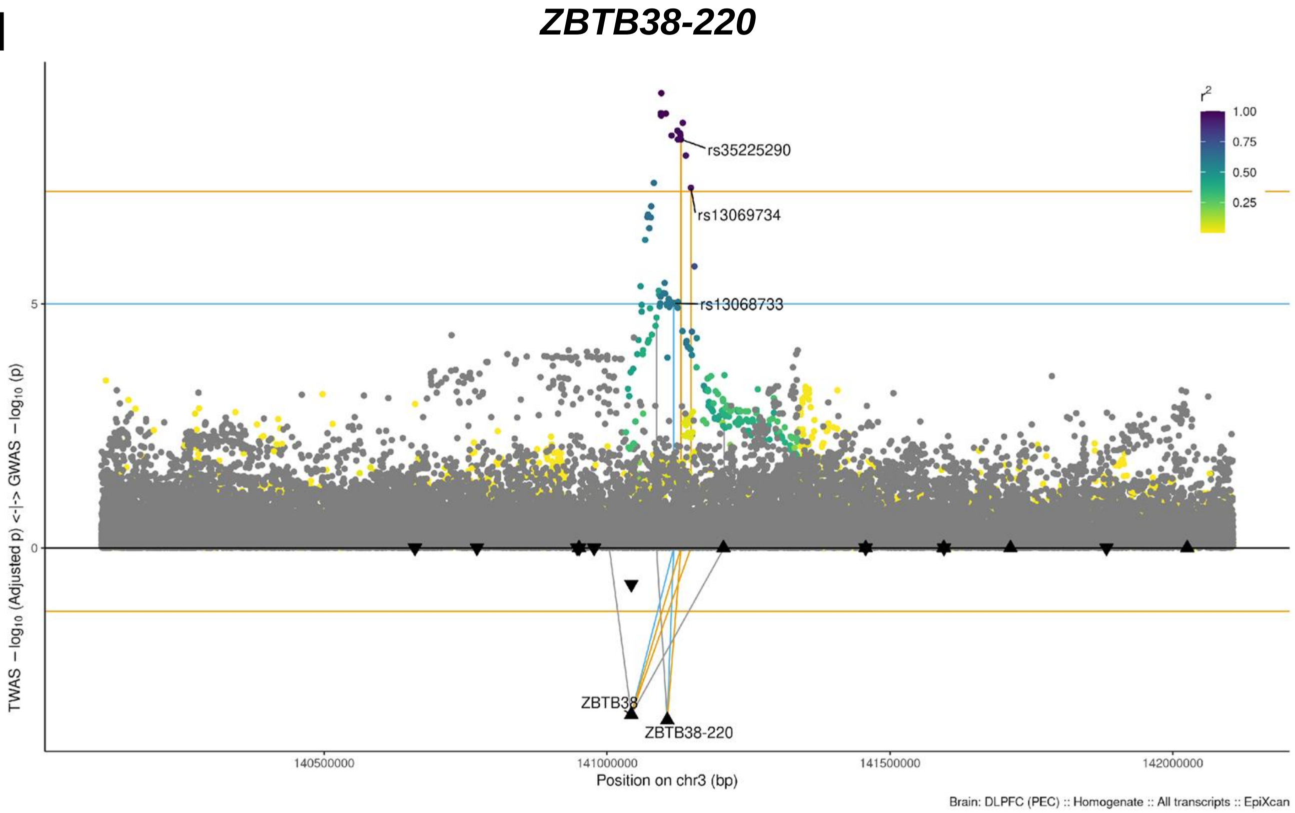
ZBTB38-220
TWAS − log₁₀ (Adjusted p) <-|-> GWAS − log₁₀ (p)
r²
1.00
0.75
0.50
0.25
rs35225290
rs13069734
rs13068733
ZBTB38
ZBTB38-220
5
0
140500000
141000000
141500000
142000000
Position on chr3 (bp)
Brain: DLPFC (PEC) :: Homogenate :: All transcripts :: EpiXcan

**I**

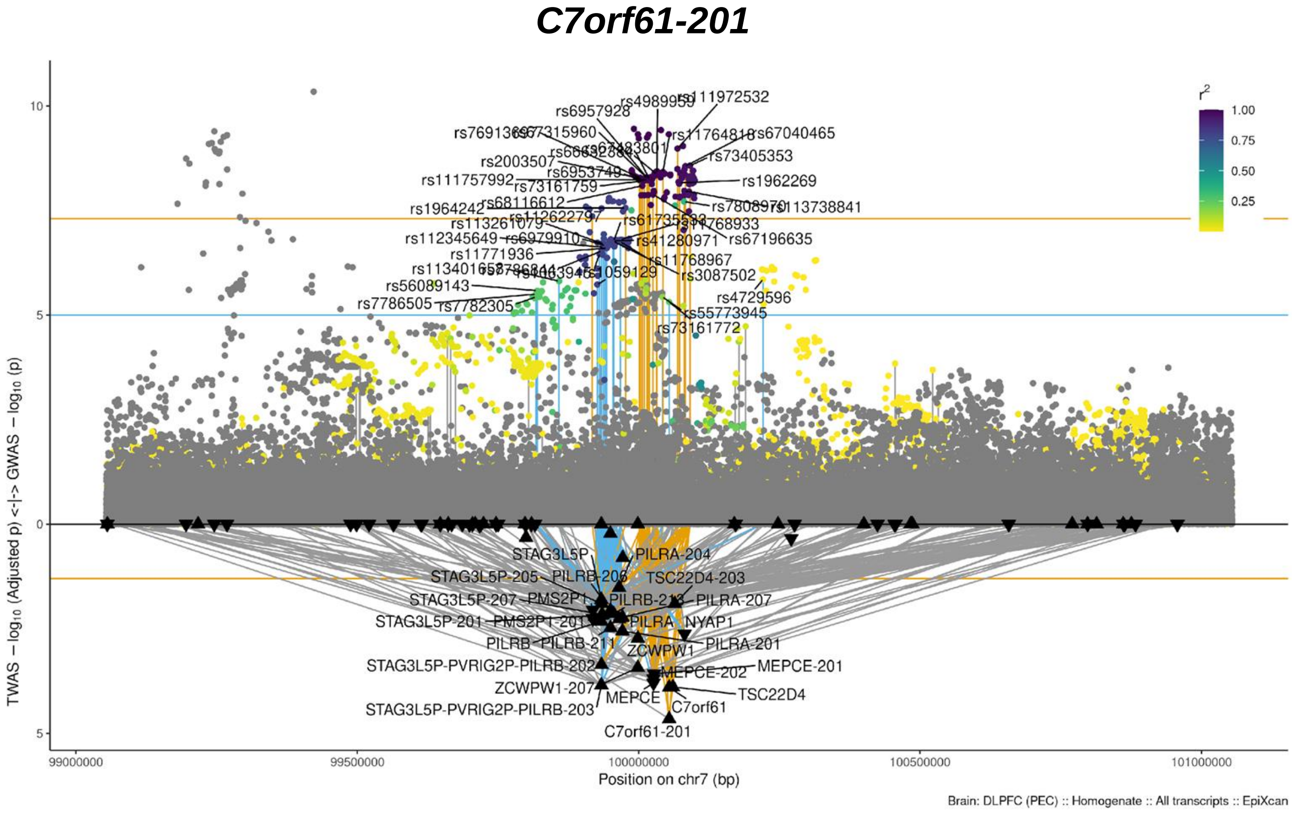
C7orf61-201
TWAS − log₁₀ (Adjusted p) <-|-> GWAS − log₁₀ (p)
r²
1.00
0.75
0.50
0.25
rs6957928
rs111972532
rs11764818
rs67040465
rs73405353
rs2003507
rs6953749
rs111757992
rs73161759
rs1962269
rs1964242
rs68116612
rs112622797
rs113261079
rs112345649
rs6979910
rs41280971
rs67196635
rs11771936
rs11768967
rs3087502
rs56089143
rs7786505
rs7782305
rs4729596
rs55773945
rs73161772
STAG3L5P
PILRA-204
STAG3L5P-205
PILRB-203
TSC22D4-203
STAG3L5P-207
PMS2P1
PILRB-213
PILRA-207
STAG3L5P-201
PMS2P1-201
PILRA
NYAP1
PILRB
PILRB-211
ZCWPW1
PILRA-201
STAG3L5P-PVRIG2P-PILRB-202
MEPCE-202
MEPCE-201
ZCWPW1-207
TSC22D4
MEPCE
C7orf61
STAG3L5P-PVRIG2P-PILRB-203
C7orf61-201
10
5
0
5
99000000
99500000
100000000
100500000
101000000
Position on chr7 (bp)
Brain: DLPFC (PEC) :: Homogenate :: All transcripts :: EpiXcan

J

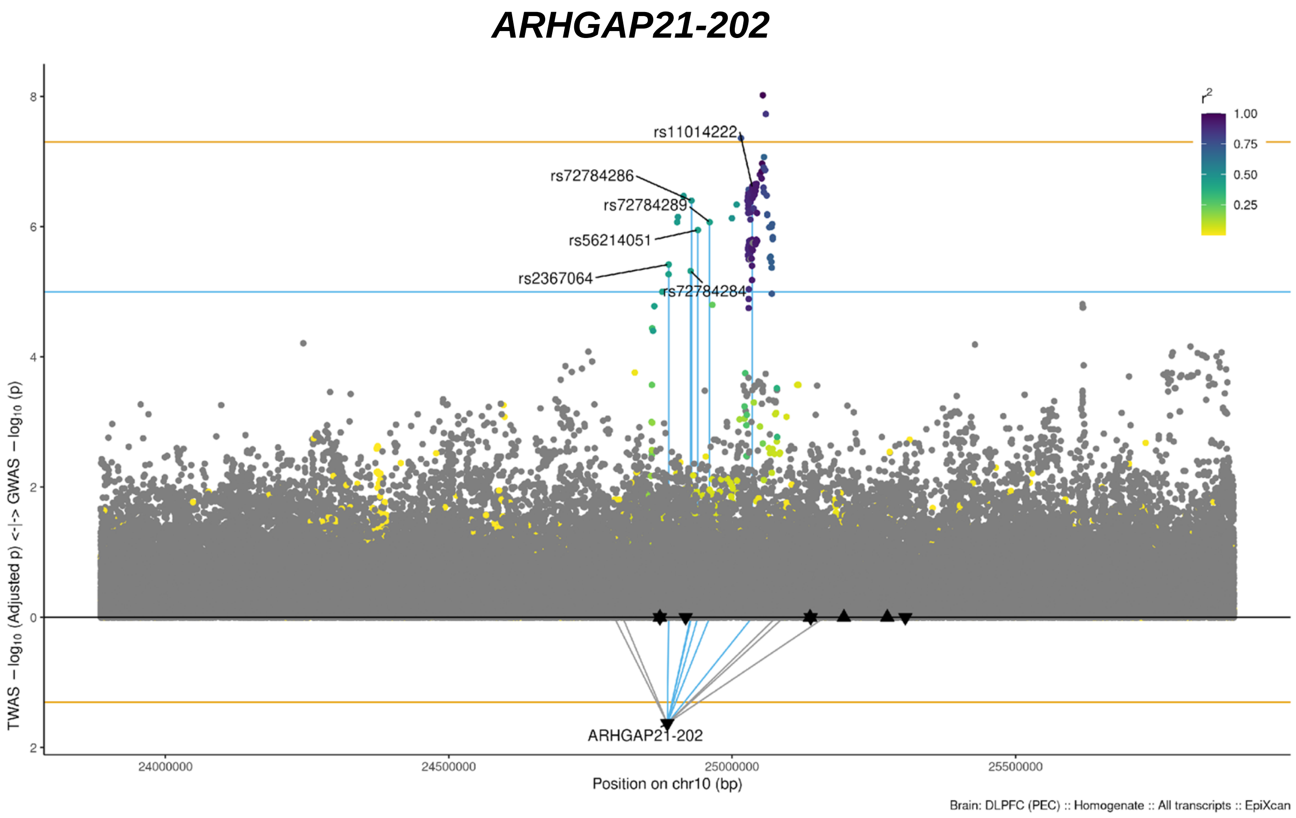
ARHGAP21-202
rs11014222
rs72784286
rs72784289
rs56214051
rs2367064
rs72784284
ARHGAP21-202
TWAS − log10 (Adjusted p) <-|-> GWAS − log10 (p)
Position on chr10 (bp)
24000000
24500000
25000000
25500000
8
6
4
2
0
2
r2
1.00
0.75
0.50
0.25
Brain: DLPFC (PEC) :: Homogenate :: All transcripts :: EpiXcan

K

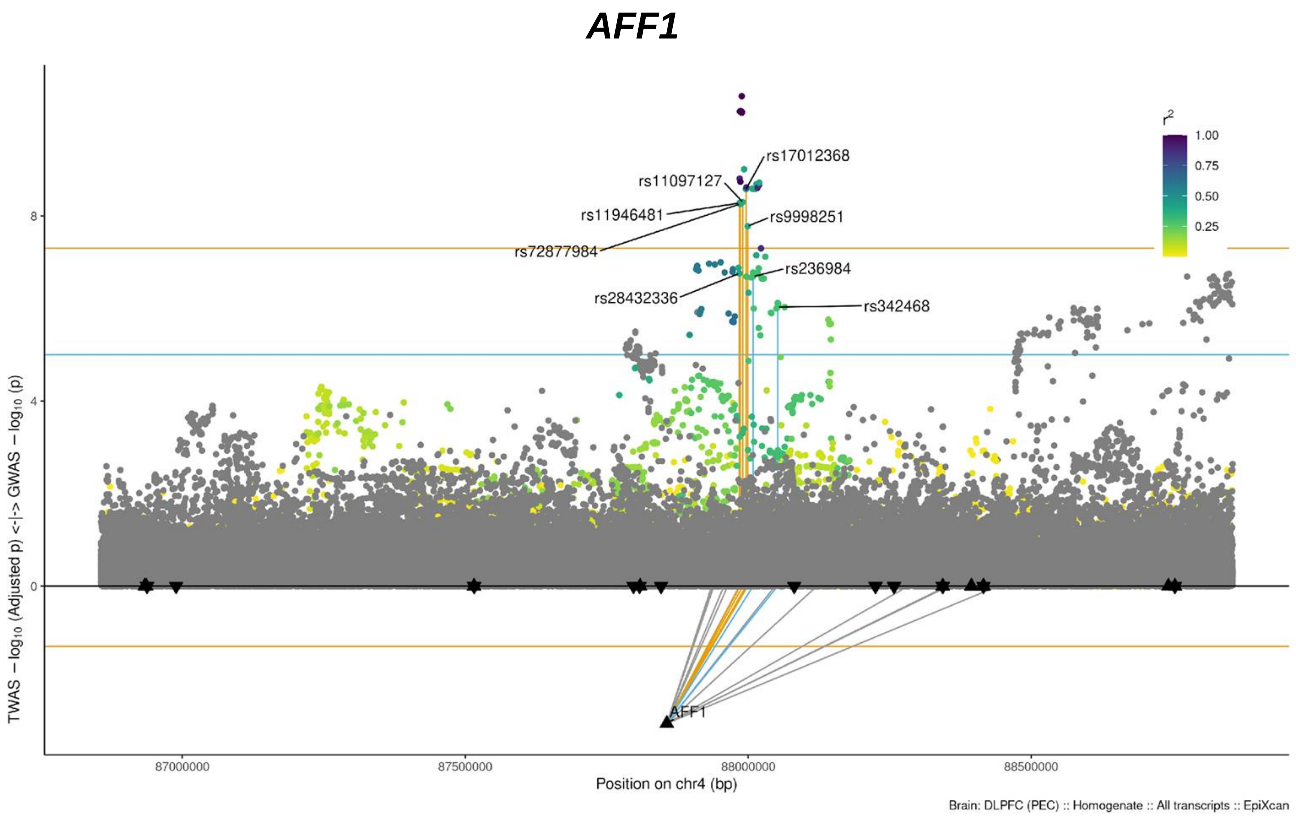
AFF1
rs17012368
rs11097127
rs11946481
rs9998251
rs72877984
rs236984
rs28432336
rs342468
AFF1
TWAS − log10 (Adjusted p) <-|-> GWAS − log10 (p)
Position on chr4 (bp)
87000000
87500000
88000000
88500000
8
4
0
r2
1.00
0.75
0.50
0.25
Brain: DLPFC (PEC) :: Homogenate :: All transcripts :: EpiXcan

L

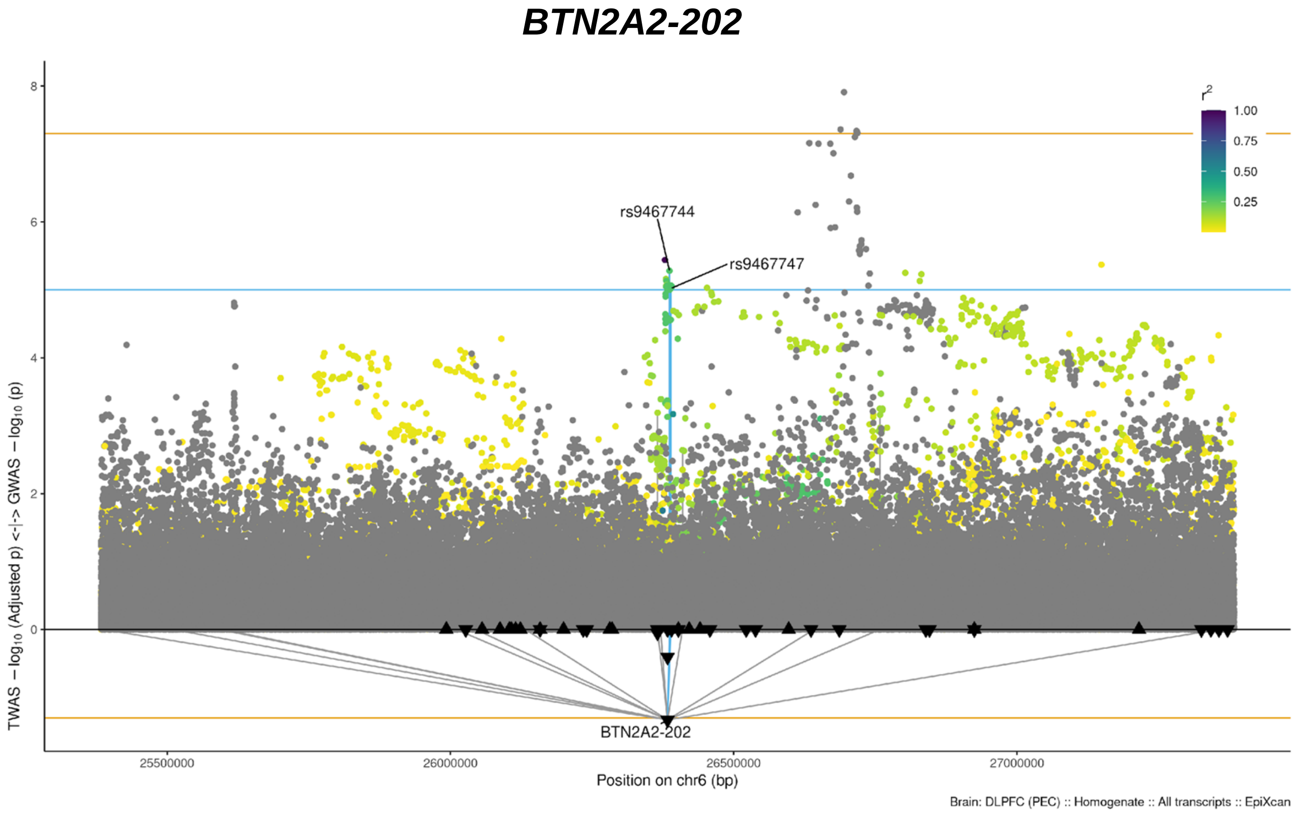
BTN2A2-202
r2
1.00
0.75
0.50
0.25
rs9467744
rs9467747
BTN2A2-202
TWAS − log10 (Adjusted p) <-|-> GWAS − log10 (p)
25500000
26000000
26500000
27000000
Position on chr6 (bp)
Brain: DLPFC (PEC) :: Homogenate :: All transcripts :: EpiXcan

M

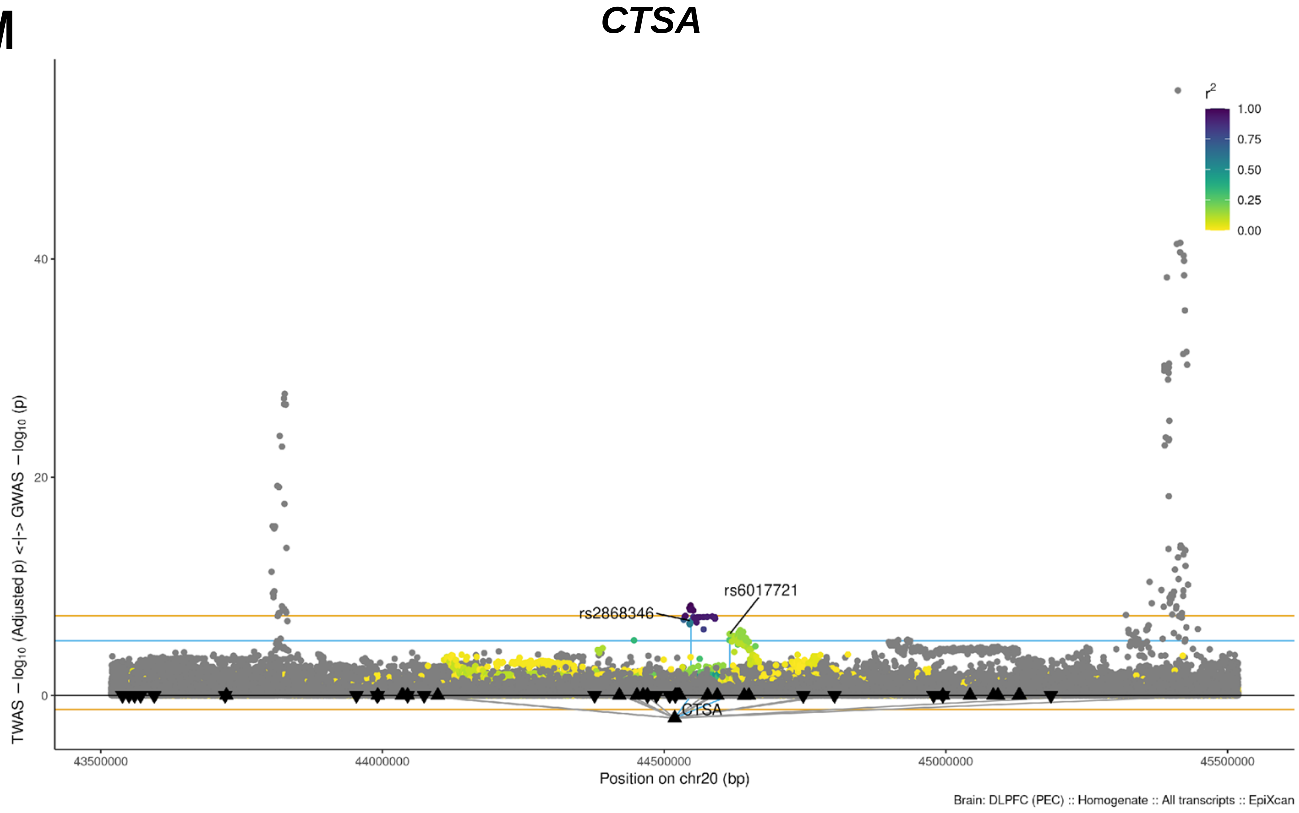
CTSA
r2
1.00
0.75
0.50
0.25
0.00
rs2868346
rs6017721
CTSA
TWAS − log10 (Adjusted p) <-|-> GWAS − log10 (p)
43500000
44000000
44500000
45000000
45500000
Position on chr20 (bp)
Brain: DLPFC (PEC) :: Homogenate :: All transcripts :: EpiXcan

N

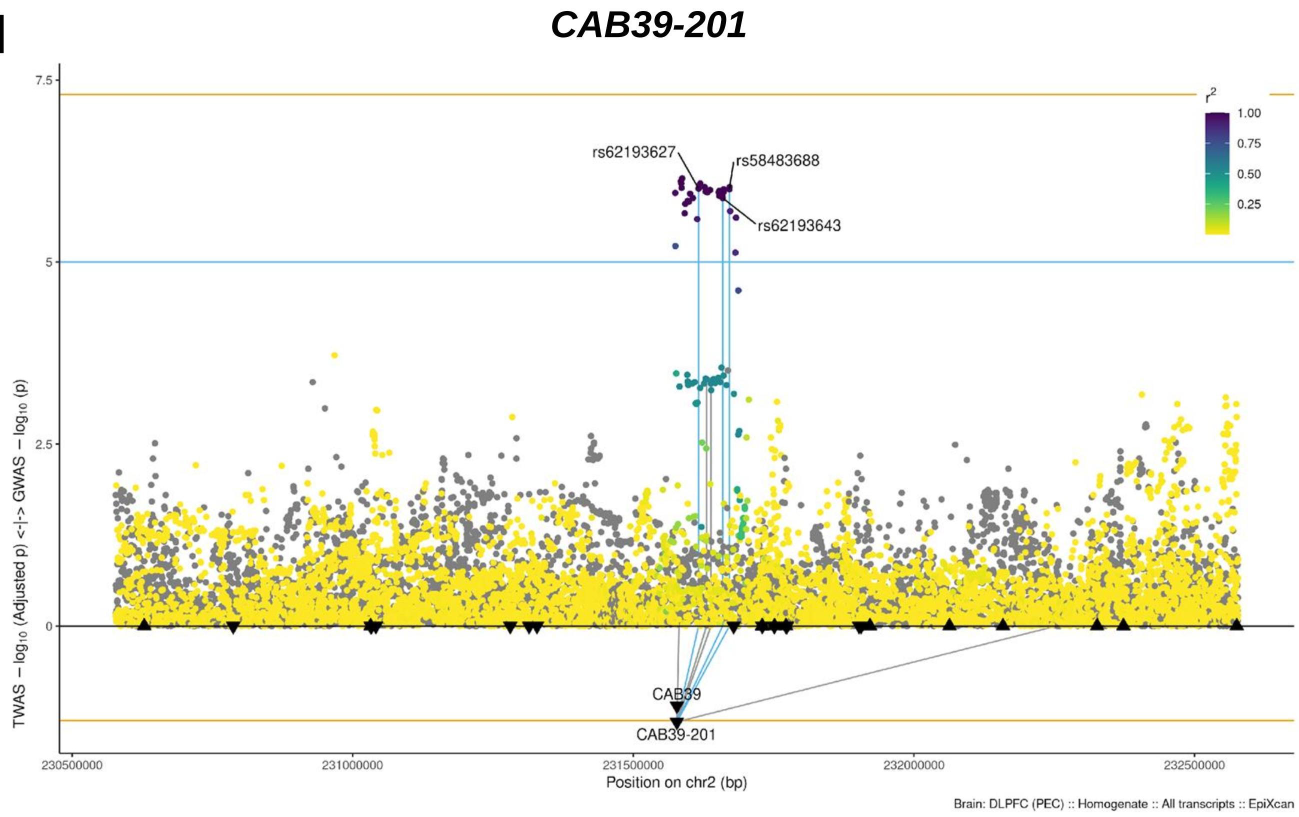
CAB39-201
rs62193627
rs58483688
rs62193643
CAB39
CAB39-201
TWAS − log10 (Adjusted p) <-|-> GWAS − log10 (p)
Position on chr2 (bp)
r2
Brain: DLPFC (PEC) :: Homogenate :: All transcripts :: EpiXcan

O

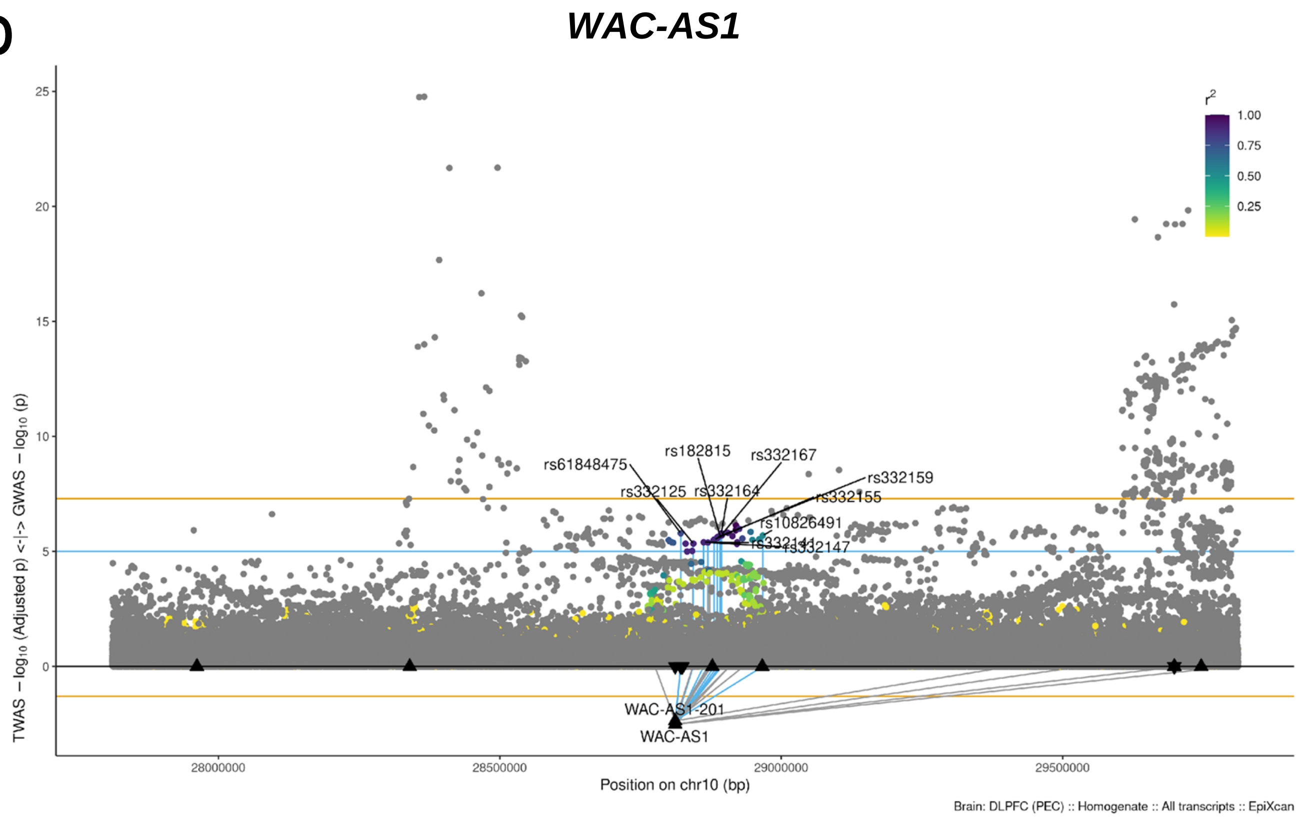
WAC-AS1
rs61848475
rs182815
rs332167
rs332159
rs332125
rs332164
rs332155
rs10826491
rs332147
WAC-AS1-201
WAC-AS1
TWAS − log10 (Adjusted p) <-|-> GWAS − log10 (p)
Position on chr10 (bp)
r2
Brain: DLPFC (PEC) :: Homogenate :: All transcripts :: EpiXcan

P

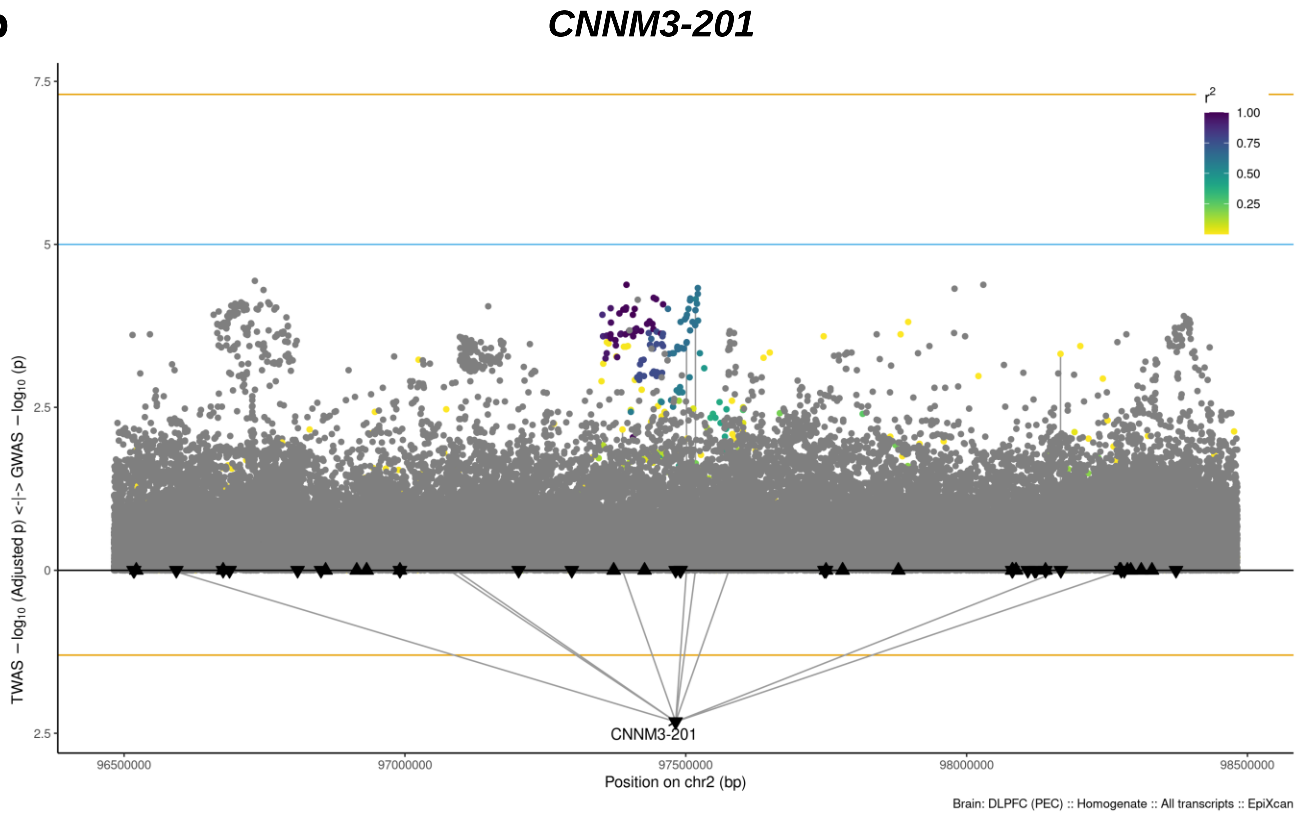
CNNM3-201
r2
1.00
0.75
0.50
0.25
7.5
5
2.5
0
2.5
TWAS − log10 (Adjusted p) <-|-> GWAS − log10 (p)
CNNM3-201
96500000
97000000
97500000
98000000
98500000
Position on chr2 (bp)
Brain: DLPFC (PEC) :: Homogenate :: All transcripts :: EpiXcan

Q

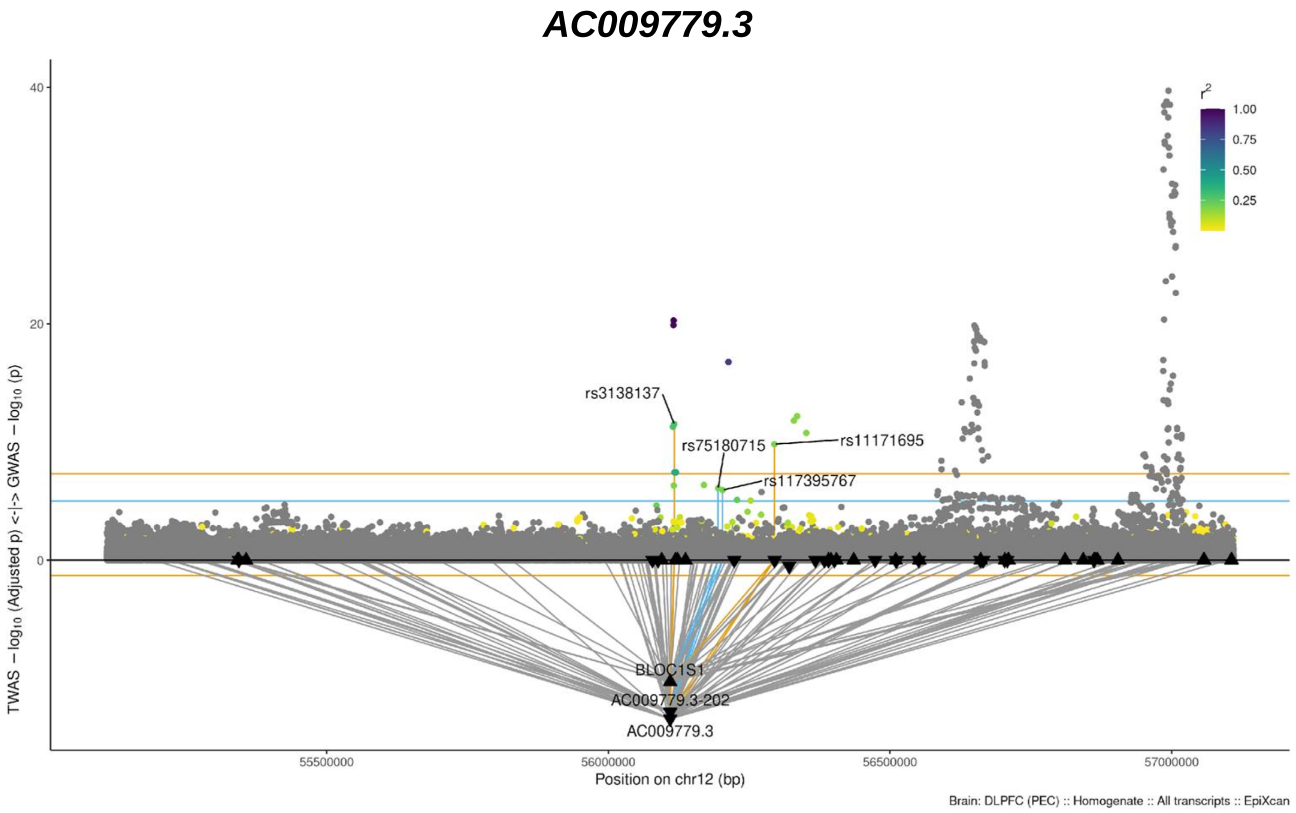
AC009779.3
r2
1.00
0.75
0.50
0.25
40
20
0
TWAS − log10 (Adjusted p) <-|-> GWAS − log10 (p)
rs3138137
rs75180715
rs11171695
rs117395767
BLOC1S1
AC009779.3-202
AC009779.3
55500000
56000000
56500000
57000000
Position on chr12 (bp)
Brain: DLPFC (PEC) :: Homogenate :: All transcripts :: EpiXcan

R

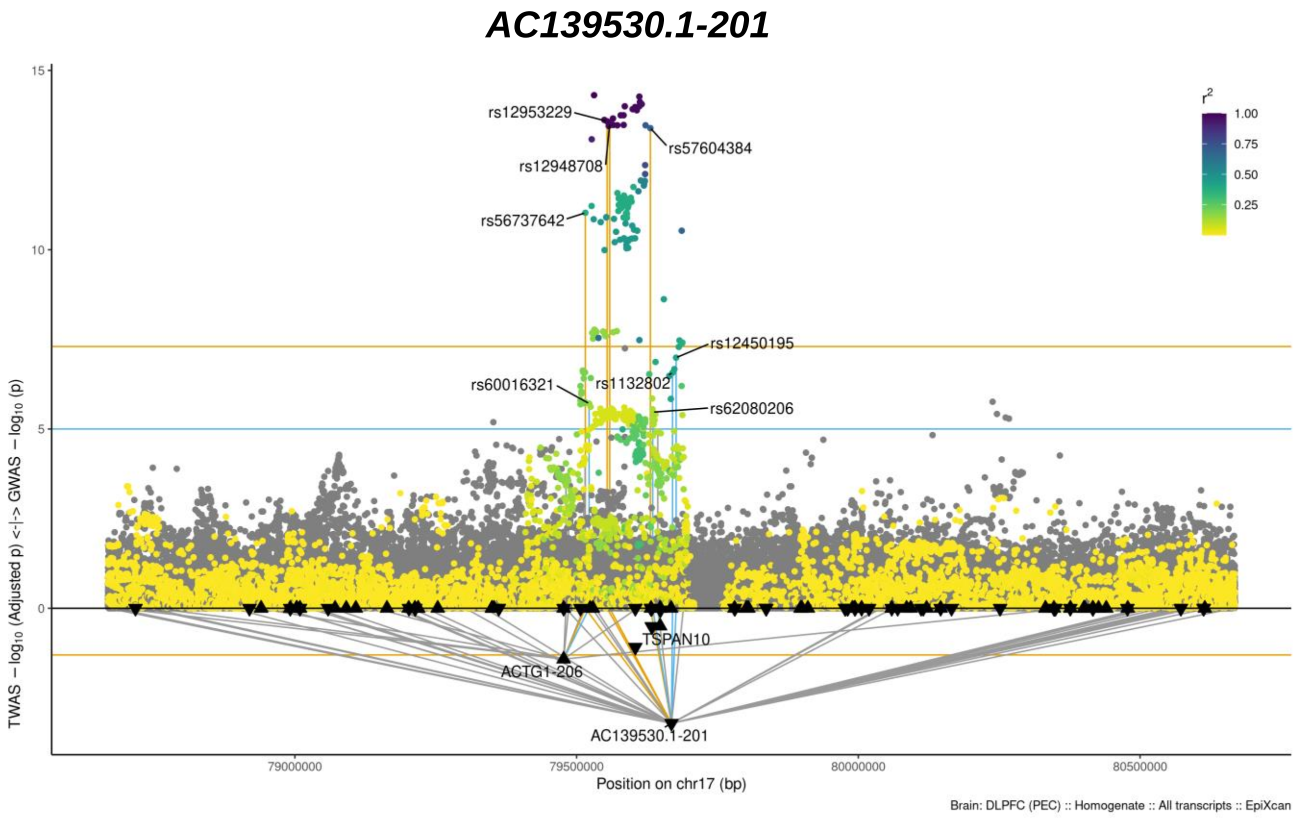
AC139530.1-201
rs12953229
rs12948708
rs57604384
rs56737642
rs12450195
rs60016321
rs1132802
rs62080206
TSPAN10
ACTG1-206
AC139530.1-201
r2
1.00
0.75
0.50
0.25
15
10
5
0
TWAS − log10 (Adjusted p) <-|-> GWAS − log10 (p)
79000000
79500000
80000000
80500000
Position on chr17 (bp)
Brain: DLPFC (PEC) :: Homogenate :: All transcripts :: EpiXcan

S

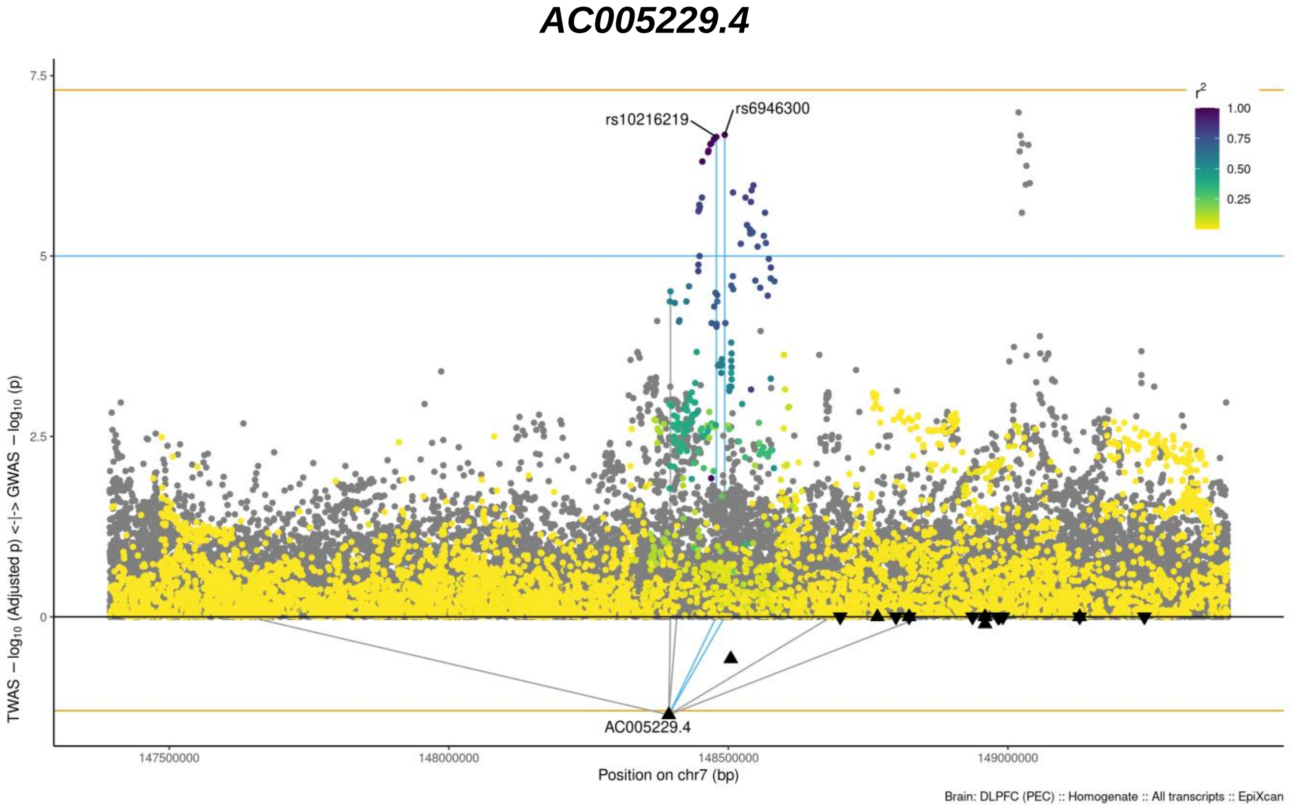
AC005229.4
rs10216219
rs6946300
AC005229.4
r2
1.00
0.75
0.50
0.25
7.5
5
2.5
0
TWAS − log10 (Adjusted p) <-|-> GWAS − log10 (p)
147500000
148000000
148500000
149000000
Position on chr7 (bp)
Brain: DLPFC (PEC) :: Homogenate :: All transcripts :: EpiXcan

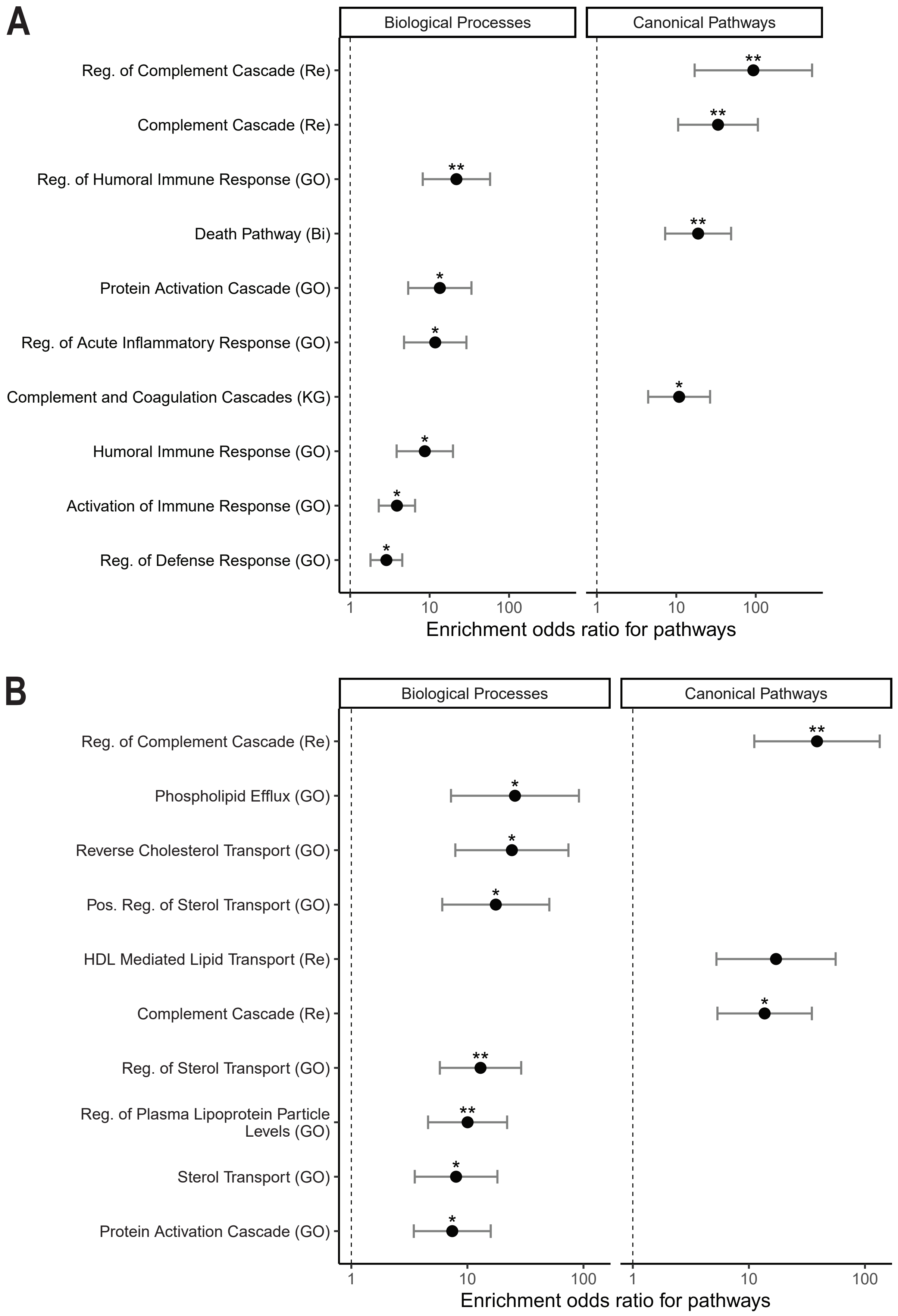


**Fig. S11. Gene set enrichment analysis (GSEA) of TWAS genes.** (**A**) GSEA of TWAS using the DLPFC transcripts (genes and isoforms) model, and (**B**) the multi-tissue meta-analysis. Shown are the top ten enrichments by effect size (odds ratio) in both analyses.

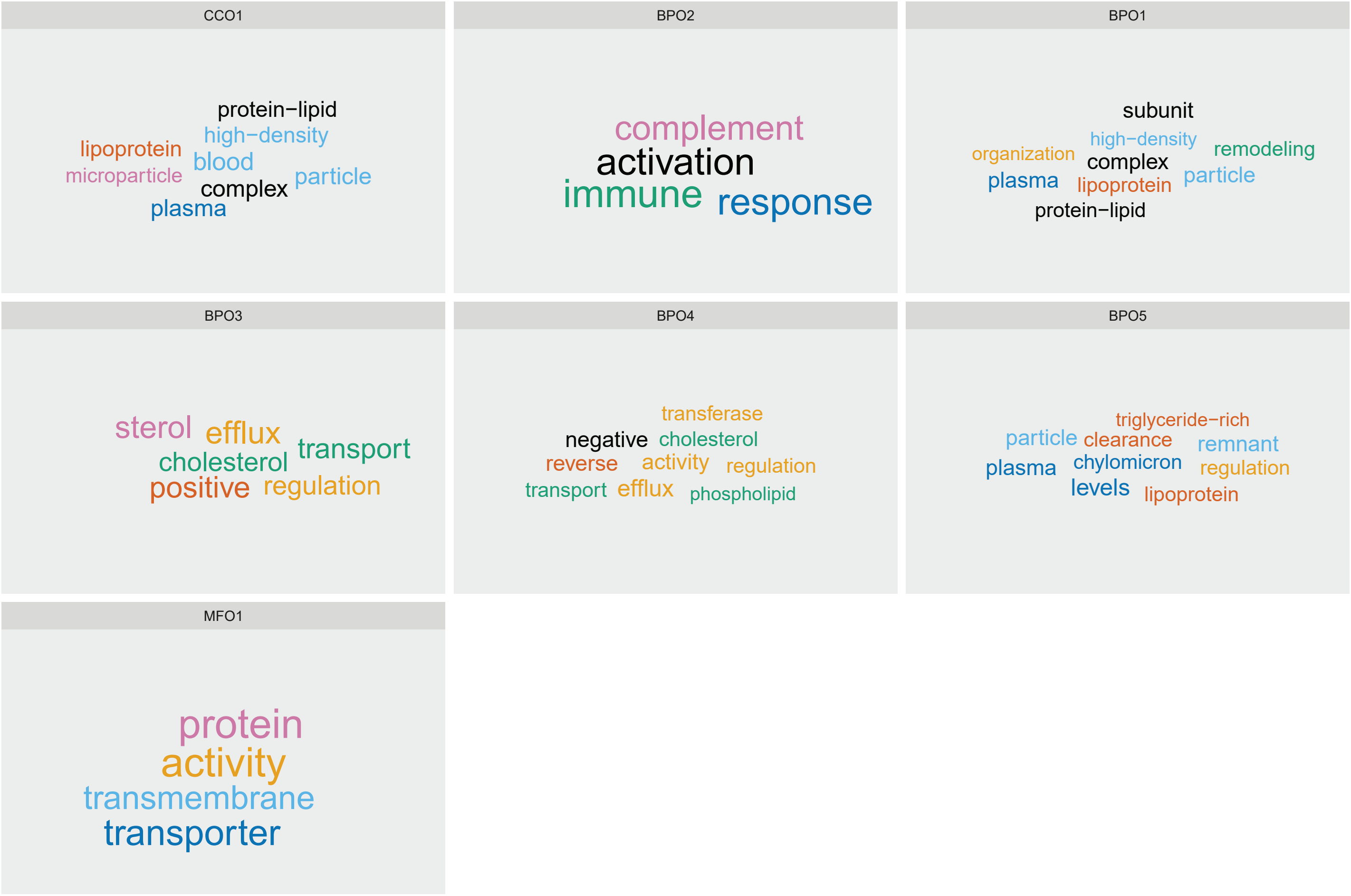


**Fig. S12. Semantic clustering of enriched gene ontology (GO) terms in the AMD multi-tissue transcriptome-wide association study (TWAS) identifies immune and lipid homeostasis functions.** First, we performed enrichment of AMD TWAS associated genes (FDR < 0.05 in ACAT meta-analysis of all tissues) for GO terms corresponding to ontologies of molecular functions (MFO), biological processes (BPO), and cellular components (CCO). Semantic clustering of the GO terms that had enrichment FDR < 0.1 (Fisher's exact test) and could be clustered identified 7 clusters (BPO1-BPO5, CCO1, and MFO1). For each of these clusters we provide a word cloud based on the description of the GO terms where words that appear more frequently are larger; clusters are ordered from left to right and top to bottom from the most significant to the least significant based on their meta-analyzed p-value (ACAT). Major themes that are associated with AMD are immune functions and lipid homeostasis.

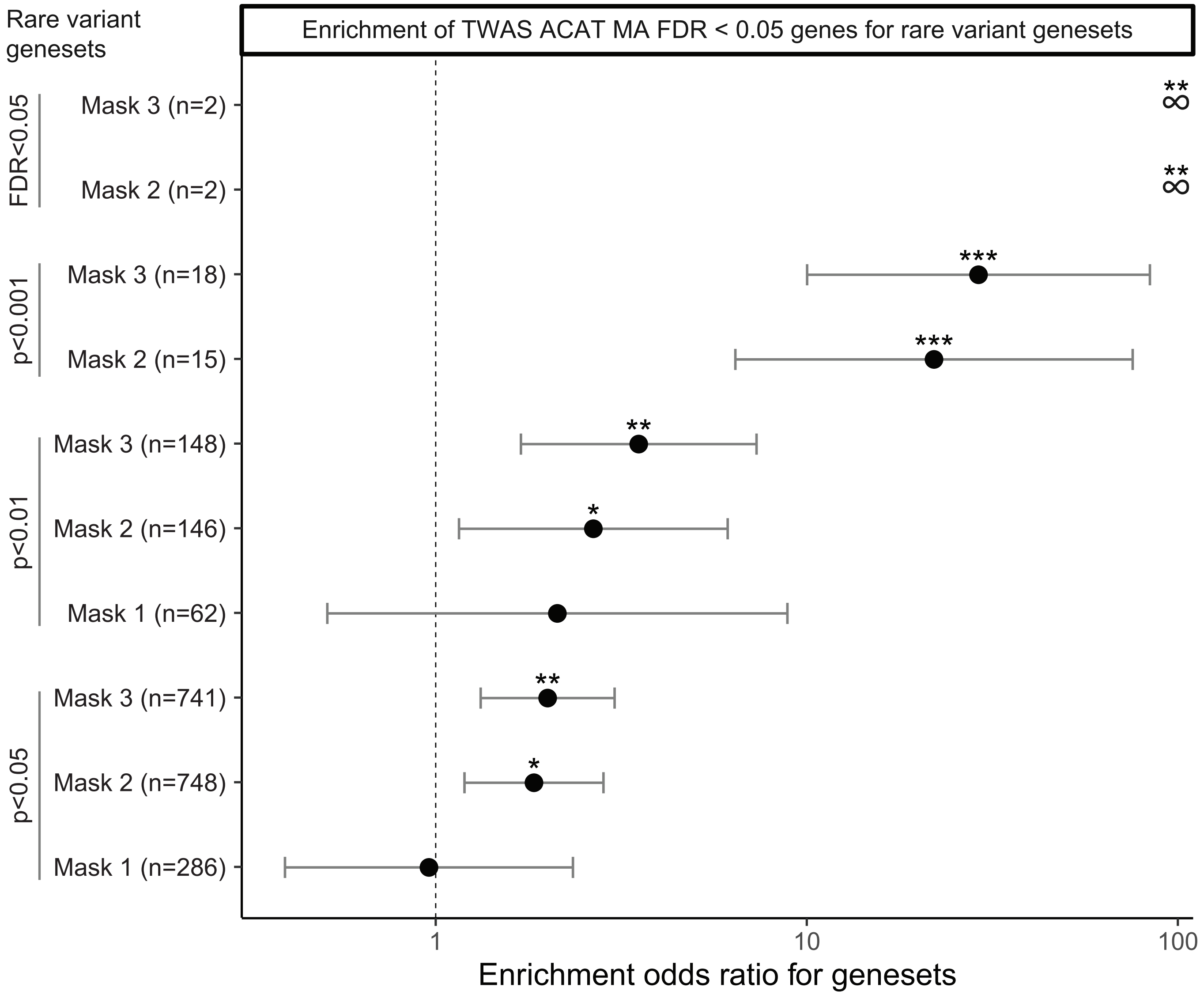


**Fig. S13. Enrichment of rare variant burden associations in TWAS meta-analysis genes.** TWAS meta-analysis genes were those with FDR < 0.05; only protein-coding genes were considered. *** FDR < 0.001, ** FDR < 0.01, * FDR < 0.05 (12 rare variant gene sets were considered from the rare variant analysis). Mask 1: Loss of function mutations only; Mask 2: Loss of function + missense mutations; Mask 3: Loss of function + missense + splice mutations.
