## Supplementary Information for "Distinctive cross-ancestry genetic architecture for age-related macular degeneration"

**Million Veteran Program: Consortium Acknowledgement for Manuscripts**

**MVP Executive Committee**

- Co-Chair: J. Michael Gaziano, M.D., M.P.H.
- Co-Chair: Rachel Ramoni, D.M.D., Sc.D.
- Jim Breeling, M.D. (ex-officio)
- Kyong-Mi Chang, M.D.
- Grant Huang, Ph.D.
- Sumitra Muralidhar, Ph.D.
- Christopher J. O'Donnell, M.D., M.P.H.
- Philip S. Tsao, Ph.D.

**MVP Program Office**

- Sumitra Muralidhar, Ph.D.
- Jennifer Moser, Ph.D.

**MVP Recruitment/Enrollment**

- Recruitment/Enrollment Director/Deputy Director, Boston
- Stacey B. Whitbourne, Ph.D.; Jessica V. Brewer, M.P.H.
- MVP Coordinating Centers
    - o Clinical Epidemiology Research Center (CERC), West Haven – John Concato, M.D., M.P.H.
    - o Cooperative Studies Program Clinical Research Pharmacy Coordinating Center, Albuquerque - Stuart Warren, J.D., Pharm D.; Dean P. Argyres, M.S.
    - o Genomics Coordinating Center, Palo Alto – Philip S. Tsao, Ph.D.
    - o Massachusetts Veterans Epidemiology Research Information Center (MAVERIC), Boston - J. Michael Gaziano, M.D., M.P.H.
    - o MVP Information Center, Canandaigua – Brady Stephens, M.S.
- Core Biorepository, Boston – Mary T. Brophy M.D., M.P.H.; Donald E. Humphries, Ph.D.
- MVP Informatics, Boston – Nhan Do, M.D.; Shahpoor Shayan
- Data Operations/Analytics, Boston – Xuan-Mai T. Nguyen, Ph.D.

**MVP Science**

- Genomics - Christopher J. O'Donnell, M.D., M.P.H.; Saiju Pyarajan Ph.D.; Philip S. Tsao, Ph.D.
- Phenomics - Kelly Cho, M.P.H, Ph.D.
- Data and Computational Sciences – Saiju Pyarajan, Ph.D.
- Statistical Genetics – Elizabeth Hauser, Ph.D.; Yan Sun, Ph.D.; Hongyu Zhao, Ph.D.

**MVP Local Site Investigators**

- Atlanta VA Medical Center (Peter Wilson) - Bay Pines VA Healthcare System (Rachel McArdle)
- Birmingham VA Medical Center (Louis Dellitalia)
- Cincinnati VA Medical Center (John Harley)
- Clement J. Zablocki VA Medical Center (Jeffrey Whittle)
- Durham VA Medical Center (Jean Beckham)
- Edith Nourse Rogers Memorial Veterans Hospital (John Wells)
- Edward Hines, Jr. VA Medical Center (Salvador Gutierrez)
- Fayetteville VA Medical Center (Gretchen Gibson)
- VA Health Care Upstate New York (Laurence Kaminsky)
- New Mexico VA Health Care System (Gerardo Villareal)
- VA Boston Healthcare System (Scott Kinlay)
- VA Western New York Healthcare System (Junzhe Xu)
- Ralph H. Johnson VA Medical Center (Mark Hamner)
- Wm. Jennings Bryan Dorn VA Medical Center (Kathlyn Sue Haddock)
- VA North Texas Health Care System (Sujata Bhushan)
- Hampton VA Medical Center (Pran Iruvanti)
- Hunter Holmes McGuire VA Medical Center (Michael Godschalk)
- Iowa City VA Health Care System (Zuhair Ballas)
- Jack C. Montgomery VA Medical Center (Malcolm Buford)
- James A. Haley Veterans' Hospital (Stephen Mastorides)
- Louisville VA Medical Center (Jon Klein)
- Louis Stokes Cleveland VA Medical Center (Frank Jacono)
- Manchester VA Medical Center (Nora Ratcliffe)
- Miami VA Health Care System (Hermes Florez)
- Michael E. DeBakey VA Medical Center (Alan Swann)
- Minneapolis VA Health Care System (Maureen Murdoch)
- N. FL/S. GA Veterans Health System (Peruvemba Sriram)
- Northport VA Medical Center (Shing Shing Yeh)
- Overton Brooks VA Medical Center (Ronald Washburn)
- Philadelphia VA Medical Center (Darshana Jhala)
- Phoenix VA Health Care System (Samuel Aguayo)
- Portland VA Medical Center (David Cohen)
- Providence VA Medical Center (Satish Sharma)

- Richard Roudebush VA Medical Center (John Callaghan)
- Salem VA Medical Center (Kris Ann Oursler)
- San Francisco VA Health Care System (Mary Whooley)
- South Texas Veterans Health Care System (Sunil Ahuja)
- Southeast Louisiana Veterans Health Care System (Amparo Gutierrez)
- Southern Arizona VA Health Care System (Ronald Schifman)
- Sioux Falls VA Health Care System (Jennifer Greco)
- St. Louis VA Health Care System (Michael Rauchman)
- Syracuse VA Medical Center (Richard Servatius)
- VA Eastern Kansas Health Care System (Mary Oehlert)
- VA Greater Los Angeles Health Care System (Agnes Wallbom)
- VA Loma Linda Healthcare System (Ronald Fernando)
- VA Long Beach Healthcare System (Timothy Morgan)
- VA Maine Healthcare System (Todd Stapley)
- VA New York Harbor Healthcare System (Scott Sherman)
- VA Pacific Islands Health Care System (Gwenevere Anderson)
- VA Palo Alto Health Care System (Philip Tsao)
- VA Pittsburgh Health Care System (Elif Sonel)
- VA Puget Sound Health Care System (Edward Boyko)
- VA Salt Lake City Health Care System (Laurence Meyer)
- VA San Diego Healthcare System (Samir Gupta)
- VA Southern Nevada Healthcare System (Joseph Fayad)
- VA Tennessee Valley Healthcare System (Adriana Hung)
- Washington DC VA Medical Center (Jack Lichy)
- W.G. (Bill) Hefner VA Medical Center (Robin Hurley)
- White River Junction VA Medical Center (Brooks Robey)
- William S. Middleton Memorial Veterans Hospital (Robert Striker)

### International Age-related Macular Degeneration Genomics Consortium (IAMDGC) members

The list of consortium members reflects the author list of the previous publication by Fritsche et al.: A large genome-wide association study of age-related macular degeneration highlights contributions of rare and common variants. Nature Genetics 2016;48:134. PMID: 26691988

Lars G Fritsche[1,100], Wilmar Igl[2,100], Jessica N Cooke Bailey[3,100], Felix Grassmann[4,100], Sebanti Sengupta[1,100], Jennifer L Bragg-Gresham[1,5], Kathryn P Burdon[6], Scott J Hebbring[7], Cindy Wen[8], Mathias Gorski[2], Ivana K Kim[9], David Cho[10], Donald Zack[11–15], Eric Souied[16], Hendrik P N Scholl[11,17], Elisa Bala[18], Kristine E Lee[19], David J Hunter[20,21], Rebecca J Sardell[22], Paul Mitchell[23], Joanna E Merriam[24], Valentina Cipriani[25,26], Joshua DHoffman[27], Tina Schick[28], Yara T E Lechanteur[29], Robyn H Guymer[30], Matthew P Johnson[31], Yingda Jiang[32], Chloe M Stanton[33], Gabriëlle H S Buitendijk[34,35], Xiaowei Zhan[1,36,37], Alan M Kwong[1], Alexis Boleda[38], Matthew Brooks[38], Linn Gieser[38], Rinki Ratnapriya[38], Kari E Branham[39], Johanna R Foerster[1], John R Heckenlively[39], Mohammad I Othman[39], Brendan J Vote[6], Helena Hai Liang[30], Emmanuelle Souzeau[40], Ian L McAllister[41], Timothy Isaacs[41], Janette Hall[40], Stewart Lake[40], David A Mackey[6,30,41], Ian J Constable[41], Jamie E Craig[40], Terrie E Kitchner[7], Zhenglin Yang[42,43], Zhiguang Su[44], Hongrong Luo[8], Daniel Chen[8], Hong Ouyang[8], Ken Flagg[8], Danni Lin[8], Guanping Mao[8], Henry Ferreyra[8], Klaus Stark[2], Claudia Nvon Strachwitz[45], Armin Wolf[46], Caroline Brandl[2,4,47], Guenther Rudolph[46], Matthias Olden[2], Margaux A Morrison[48], Denise J Morgan[48], Matthew Schu[49–53], Jeeyun Ahn[54], Giuliana Silvestri[55], Evangelia E Tsironi[56], Kyu Hyung Park[57], Lindsay A Farrer[49–53], Anton Orlin[58], Alexander Brucker[59], Mingyao Li[60], Christine A Curcio[61], Saddek Mohand-Saïd[62–65], José-Alain Sahel[25,62–67], Isabelle Audo[62–64,68], Mustapha Benchaboune[65], Angela J Cree[69], Christina A Rennie[70], Srinivas V Goverdhan[69], Michelle Grunin[71], Shira Hagbi-Levi[71], Peter Campochiaro[11,13], Nicholas Katsanis[72–74], Frank GHolz[17], Frédéric Blond[62–64], Hélène Blanché[75], Jean-François Deleuze[75,76], Robert P Igo Jr[3], Barbara Truitt[3], Neal S Peachey[18,77], Stacy M Meuer[19], Chelsea E Myers[19], Emily L Moore[19], Ronald Klein[19], Michael A Hauser[78–80], Eric A Postel[78], Monique D Courtenay[22], Stephen G Schwartz[81], Jaclyn L Kovach[81], William K Scott[22], Gerald Liew[23], Ava G Tan[23], Bamini Gopinath[23], John C Merriam[24], R Theodore Smith[24,82], Jane C Khan[41,83,84], Humma Shahid[84,85], Anthony T Moore[25,26,86], J Allie McGrath[27], Reneé Laux[3], Milam A Brantley Jr[87], Anita Agarwal[87], Lebriz Ersoy[28], Albert Caramoy[28], Thomas Langmann[28], Nicole T M Saksens[29], Eiko K deJong[29], Carel B Hoyng[29], Melinda S Cain[30], Andrea J Richardson[30], Tammy M Martin[88], John Blangero[31], Daniel E Weeks[32,89], Bal Dhillon[90], Cornelia Mvan Duijn[35], Kimberly F Doheny[91], Jane Romm[91], Caroline C W Klaver[34,35], Caroline Hayward[33], Michael B Gorin[92,93], Michael L Klein[88], Paul N Baird[30], Anneke I den Hollander[29,94], Sascha Fauser[28], John R W Yates[25,26,84], Rando Allikmets[24,95], Jie Jin Wang[23], Debra A Schaumberg[20,96,97], Barbara EKKlein[19], Stephanie A Hagstrom[77], Itay Chowers[71], Andrew J Lotery[69], Thierry Léveillard[62–64], Kang Zhang[8,44], Murray H Brilliant[7], Alex WHewitt[6,30,41], Anand Swaroop[38], Emily Y Chew[98], Margaret A Pericak-Vance[22,101], Margaret DeAngelis[48,101], Dwight Stambolian[10,101], Jonathan L Haines[3,99,101], Sudha K Iyengar[3,101], Bernhard H F Weber[4,101], Gonçalo R Abecasis[1,101] & Iris MHeid[2,101]

[1]Center for Statistical Genetics, Department of Biostatistics, University of Michigan, Ann Arbor, Michigan, USA. [2]Department of Genetic Epidemiology, University of Regensburg, Regensburg, Germany. [3]Department of Epidemiology and Biostatistics, Case Western Reserve University School of Medicine, Cleveland, Ohio, USA. [4]Institute of Human Genetics, University of Regensburg, Regensburg, Germany. [5]Kidney Epidemiology and Cost Center, Department of Internal Medicine–Nephrology, University of Michigan, Ann Arbor, Michigan, USA. [6]School of Medicine, Menzies Research Institute Tasmania, University of Tasmania, Hobart, Tasmania, Australia. [7]Center for Human Genetics, Marshfield Clinic

Research Foundation, Marshfield, Wisconsin, USA. [8]Department of Ophthalmology, University of California, San Diego and Veterans Affairs San Diego Health System, La Jolla, California, USA. [9]Retina Service, Massachusetts Eye and Ear, Department of Ophthalmology, Harvard Medical School, Boston, Massachusetts, USA. [10]Department of Ophthalmology, Perelman School of Medicine, University of Pennsylvania, Philadelphia, Pennsylvania, USA. [11]Department of Ophthalmology, Wilmer Eye Institute, Johns Hopkins University School of Medicine, Baltimore, Maryland, USA. [12]Department of Molecular Biology and Genetics, Johns Hopkins University School of Medicine, Baltimore, Maryland, USA. [13]Department of Neuroscience, Johns Hopkins University School of Medicine, Baltimore, Maryland, USA. [14]Institute of Genetic Medicine, Johns Hopkins University School of Medicine, Baltimore, Maryland, USA. [15]Institue de la Vision, Université Pierre et Marie Curie, Paris, France. [16]Hôpital Intercommunal de Créteil, Hôpital Henri Mondor, Université Paris Est Créteil, Créteil, France. [17]Department of Ophthalmology, University of Bonn, Bonn, Germany. [18]Louis Stokes Cleveland Veterans Affairs Medical Center, Cleveland, Ohio, USA. [19]Department of Ophthalmology and Visual Sciences, University of Wisconsin, Madison, Wisconsin, USA. [20]Department of Epidemiology, Harvard School of Public Health, Boston, Massachusetts, USA. [21]Department of Nutrition, Harvard School of Public Health, Boston, Massachusetts, USA. [22]John P. Hussman Institute for Human Genomics, Miller School of Medicine, University of Miami, Miami, Florida, USA. [23]Centre for Vision Research, Department of Ophthalmology and Westmead Millennium Institute for Medical Research, University of Sydney, Sydney, New South Wales, Australia. [24]Department of Ophthalmology, Columbia University, New York, New York, USA. [25]University College London Institute of Ophthalmology, University College London, London, UK. [26]Moorfields Eye Hospital, London, UK. [27]Center for Human Genetics Research, Vanderbilt University Medical Center, Nashville, Tennessee, USA. [28]Department of Ophthalmology, University Hospital of Cologne, Cologne, Germany. [29]Department of Ophthalmology, Radboud University Medical Centre, Nijmegen, the Netherlands. [30]Centre for Eye Research Australia, University of Melbourne, Royal Victorian Eye and Ear Hospital, East Melbourne, Victoria, Australia. [31]South Texas Diabetes and Obesity Institute, School of Medicine, University of Texas Rio Grande Valley, Brownsville, Texas, USA. [32]Department of Biostatistics, Graduate School of Public Health, University of Pittsburgh, Pittsburgh, Pennsylvania, USA. [33]Medical Research Council (MRC) Human Genetics Unit, Institute of Genetics and Molecular Medicine, University of Edinburgh, Edinburgh, UK. [34]Department of Ophthalmology, Erasmus Medical Center, Rotterdam, the Netherlands. [35]Department of Epidemiology, Erasmus Medical Center, Rotterdam, the Netherlands. [36]Quantitative Biomedical Research Center, Department of Clinical Science, University of Texas Southwestern Medical Center, Dallas, Texas, USA. [37]Center for the Genetics of Host Defense, University of Texas Southwestern Medical Center, Dallas, Texas, USA. [38]Neurobiology, Neurodegeneration and Repair Laboratory (N-NRL), National Eye Institute, US National Institutes of Health, Bethesda, Maryland, USA. [39]Department of Ophthalmology and Visual Sciences, University of Michigan, Kellogg Eye Center, Ann Arbor, Michigan, USA. [40]Department of Ophthalmology, Flinders Medical Centre, Flinders University, Adelaide, South Australia, Australia. [41]Centre for Ophthalmology and Visual Science, Lions Eye Institute, University of Western Australia, Perth, Western Australia, Australia. [42]Sichuan Provincial Key Laboratory for Human Disease Gene Study, Hospital of the University of Electronic Science and Technology of China and Sichuan Provincial People's Hospital, Chengdu, China. [43]Sichuan Translational Medicine Hospital, Chinese Academy of Sciences, Chengdu, China. [44]Molecular Medicine Research Center, State Key Laboratory of Biotherapy, West China Hospital, Sichuan University, Chengdu, China. [45]EyeCentre Southwest, Stuttgart, Germany. [46]University Eye Clinic, Ludwig Maximilians University, Munich, Germany. [47]Department of Ophthalmology, University Hospital Regensburg, Regensburg, Germany. [48]Department of Ophthalmology and Visual Sciences, University of Utah, Salt Lake City, Utah, USA. [49]Department of Medicine (Biomedical Genetics), Boston University Schools of Medicine and Public Health, Boston, Massachusetts, USA. [50]Department of Ophthalmology, Boston University Schools of Medicine and Public Health, Boston, Massachusetts, USA. [51]Department of Neurology, Boston University Schools of Medicine and Public Health, Boston, Massachusetts, USA. [52]Department of Epidemiology, Boston University Schools of Medicine and Public Health, Boston, Massachusetts, USA. [53]Department of Biostatistics, Boston University Schools of Medicine and Public Health, Boston, Massachusetts, USA. [54]Department of Ophthalmology, Seoul Metropolitan Government Seoul National University Boramae Medical Center, Seoul, Republic of Korea. [55]Centre for Experimental Medicine, Queen's University, Belfast, UK. [56]Department of Ophthalmology, University of Thessaly, School of Medicine, Larissa, Greece. [57]Department of Ophthalmology, Seoul National University Bundang Hospital, Seongnam, Republic of Korea. [58]Department of Ophthalmology, Weill Cornell Medical College, New York, New York, USA. [59]Scheie Eye Institute, Department of Ophthalmology, University of Pennsylvania Perelman School of Medicine, Philadelphia, Pennsylvania, USA. [60]Department of Biostatistics and Epidemiology, University of Pennsylvania Perelman School of Medicine, Philadelphia, Pennsylvania, USA. [61]Department of Ophthalmology, University of Alabama at Birmingham, Birmingham, Alabama, USA. [62]INSERM, Paris, France. [63]Institut de la Vision, Department of Genetics, Paris, France. [64]Centre National de la Recherche Scientifique (CNRS), Paris, France. [65]Centre Hospitalier National d'Ophtalmologie des Quinze-Vingts, Paris, France. [66]Fondation Ophtalmologique Adolphe de Rothschild, Paris, France. [67]Académie des Sciences–Institut de France, Paris, France. [68]Department of Molecular Genetics, Institute of Ophthalmology, London, UK. [69]Clinical and Experimental Sciences, Faculty of Medicine, University of Southampton, Southampton, UK. [70]University Hospital Southampton, Southampton, UK. [71]Department of Ophthalmology, Hadassah Hebrew University Medical Center, Jerusalem, Israel. [72]Center for Human Disease Modeling, Duke University, Durham, North Carolina, USA. [73]Department of Cell Biology, Duke University, Durham, North Carolina, USA. [74]Department of Pediatrics, Duke University, Durham, North Carolina, USA. [75]Centre d'Etude du Polymorphisme Humain (CEPH) Fondation Jean Dausset, Paris, France. [76]Commissariat à l'Energie Atomique et aux Energies Alternatives (CEA), Institut de Génomique, Centre National de Génotypage, Evry, France. [77]Cole Eye Institute, Cleveland Clinic, Cleveland, Ohio, USA. [78]Department of Ophthalmology, Duke University Medical Center, Durham, North Carolina, USA. [79]Department of Medicine, Duke University Medical Center, Durham, North Carolina, USA. [80]Duke Molecular Physiology Institute, Duke University Medical Center, Durham, North Carolina, USA. [81]Bascom Palmer Eye Institute, University of Miami Miller School of Medicine, Naples, Florida, USA. [82]Department of Ophthalmology, New York University School of Medicine, New York, New York, USA. [83]Department of Ophthalmology, Royal Perth Hospital, Perth, Western Australia, Australia. [84]Department of Medical Genetics, Cambridge Institute for Medical Research, University of Cambridge, Cambridge, UK. [85]Department of Ophthalmology, Cambridge University Hospitals National Health Service (NHS) Foundation Trust, Cambridge, UK. [86]Department of Ophthalmology, University of California San Francisco Medical School, San Francisco, California, USA. [87]Department of Ophthalmology and Visual Sciences, Vanderbilt University, Nashville, Tennessee, USA. [88]Casey Eye Institute, Oregon Health and Science University, Portland, Oregon, USA. [89]Department of Human Genetics, Graduate School of Public Health, University of Pittsburgh, Pittsburgh, Pennsylvania, USA. [90]School of Clinical Sciences, University of Edinburgh, Edinburgh, UK. [91]Center for Inherited Disease Research (CIDR) Institute of Genetic Medicine, Johns Hopkins University School of Medicine, Baltimore, Maryland, USA. [92]Department of Ophthalmology, David Geffen School of Medicine, Stein Eye Institute, University of California, Los Angeles, Los Angeles, California, USA. [93]Department of Human Genetics, David Geffen School of Medicine, University of California, Los Angeles, Los Angeles, California, USA. [94]Department of Human Genetics, Radboud University Medical Centre, Nijmegen, the Netherlands. [95]Department of Pathology and Cell Biology, Columbia University, New York, New York, USA. [96]Center for Translational Medicine, Moran Eye Center, University of Utah School of Medicine, Salt Lake City, Utah, USA. [97]Division of Preventive Medicine, Brigham and Women's Hospital, Harvard Medical School, Boston, Massachusetts, USA. [98]Division of Epidemiology and Clinical Applications, Clinical Trials Branch, National Eye Institute, US National Institutes of Health, Bethesda, Maryland, USA. [99]Institute for Computational Biology, Case

Western Reserve University School of Medicine, Cleveland, Ohio, USA. [100]These authors contributed equally to this work. [101]These authors jointly supervised this work.
